# Real-time forecasting the trajectory of monkeypox outbreaks at the national and global levels, July – October 2022

**DOI:** 10.1101/2022.11.02.22281844

**Authors:** Amanda Bleichrodt, Sushma Dahal, Kevin Maloney, Lisa Casanova, Ruiyan Luo, Gerardo Chowell

## Abstract

**Background:** Beginning May 7, 2022, multiple nations reported an unprecedented surge in monkeypox cases. Unlike past outbreaks, differences in affected populations, transmission mode, and clinical characteristics have been noted. With the existing uncertainties of the outbreak, real-time short-term forecasting can guide and evaluate the effectiveness of public health measures.

**Methods:** We obtained publicly available data on confirmed weekly cases of monkeypox at the global level and for seven countries (with the highest burden of disease at the time this study was initiated) from the Our World in Data (OWID) GitHub repository and CDC website. We generated short-term forecasts of new cases of monkeypox across the study areas using an ensemble n-sub-epidemic modeling framework based on weekly cases using 10-week calibration periods. We report and assess the weekly forecasts with quantified uncertainty from the top-ranked, second-ranked, and ensemble sub-epidemic models. Overall, we conducted 324 weekly sequential 4-week ahead forecasts across the models from the week of July 28th, 2022, to the week of October 13th, 2022.

**Results:** The last 10 of 12 forecasting periods (starting the week of August 11^th^, 2022), show either a plateauing or declining trend of monkeypox cases for all models and areas of study. According to our latest 4-week ahead forecast from the top-ranked model, a total of 6232 (95% PI: 487.8, 12468.0) cases could be added globally from the week of 10/20/2022 to the week of 11/10/2022. At the country level, the top-ranked model predicts that the United States will report the highest cumulative number of new cases for the 4-week forecasts (median based on OWID data: 1806 (95% PI: 0.0, 5544.5)). The top-ranked and weighted ensemble models outperformed all other models in short-term forecasts.

**Conclusions:** Our top-ranked model consistently predicted a decreasing trend in monkeypox cases on the global and country-specific scale during the last ten sequential forecasting periods. Our findings reflect the potential impact of increased immunity, and behavioral modification among high-risk populations.

## Background

On May 7th, 2022, a case of monkeypox (mpox) with recent travel to Nigeria was reported in England [1, 2]. Shortly after, the Centers for Disease Control and Prevention (CDC) identified a case of monkeypox in Massachusetts on May 15th, 2022 [3]. Since then, multiple nations, including the U.S., have reported a surge in monkeypox cases, mostly among males within the communities of gay, bisexual, and other men who have sex with men (MSM) [4–6]. The World Health Organization (WHO) declared monkeypox a global health emergency on July 23rd, 2022 [7]. As of November 23rd, 2022, over 79,900 monkeypox cases have been reported in non-endemic countries, especially in the US, Spain, and Brazil, during the ongoing outbreak [8]. Given that this is an emerging infection in non-endemic countries, with little historical information about how outbreaks might unfold, mathematical models can help generate real-time forecasts of the trajectory of the epidemics and guide public health measures appropriate for a given geographic setting.

Monkeypox is an endemic zoonotic virus in Africa, most similar in clinical presentation to the Variola (Smallpox) virus [9]. Both are part of the *Orthopoxvirus* genus, which includes other viruses such as cowpox and Vaccinia virus - used in smallpox vaccines [9, 10]. Monkeypox symptoms include, but are not limited to, flu-like symptoms followed by a raised rash on the face and extremities. The incubation period is usually 6-13 days, with a symptomatic period ranging from 2 to 4 weeks [9]. Fortunately, monkeypox is not an airborne pathogen. Instead, transmission is mainly driven by prolonged close contact with infected individuals or direct contact with skin lesions, respiratory secretions, or recently contaminated objects - a feature that may facilitate control through basic public health measures [9].

The ability to forecast country-specific epidemic trajectories is particularly useful in an outbreak like this, which paints a unique epidemiological picture compared to past outbreaks within both endemic and non-endemic countries [4, 6, 11, 12]. For instance, sexual and intimate contact, specifically between men, has driven the great majority of infections [5, 13, 14]. Likewise, over 98% of cases in the United States and Spain are male, and most have identified as MSM [13, 15]. Cases of the ongoing outbreak are less likely to report prodromal symptoms, and rash occurs most frequently in the genital region [4, 13]. The scale of community spread is unprecedented [16].

While much has been learned about the epidemiology of this emerging outbreak during the last few months, substantial uncertainties remain about the effect of several variables on the epidemic trajectory, including the frequency and role of asymptomatic individuals, the role of pre-existing immunity from previous smallpox immunization campaigns, and the efficacy of available vaccines [17]. In this context, semi-mechanistic growth models are especially suitable for conducting short-term forecasts to guide response efforts and evaluate the impact of control measures, including behavior changes that mitigate transmission rates, contact tracing, and vaccination, on growth trends [18]. Previous real-time forecasting studies have employed a variety of mathematical models in the context of influenza, SARS, Ebola, and COVID-19 [19–23]. Here, we employ an ensemble sub-epidemic modeling framework to characterize epidemic trajectories that result from sub-epidemics aggregation through an optimization process [19]. This framework has yielded competitive performance in short-term forecasts of various infectious disease outbreaks [19, 23]. In this study, we generate 4-week ahead forecasts of laboratory-confirmed cases of monkeypox in near real-time at the global level and for nations that have reported the great majority of the cases: Brazil, Canada, England, France, Germany, Spain, and the United States. We also evaluate the model fit and performance of the forecasts based on the mean absolute error (MAE), mean square error (MSE), 95% prediction interval coverage (PI), and weighted interval score (WIS).

## Methods

### Data

We obtained weekly updates of the daily confirmed monkeypox cases by reporting date from publicly available sources by the CDC and the Our World in Data (OWID) GitHub repository [24, 25]. At the global level and for countries that have reported the great majority of the cases, including Brazil, Canada, England, France, Germany, Spain, and the United States, we retrieved daily case series from the GitHub Our World in Data (OWID) repository [8, 25]. We reported forecasts based on CDC and OWID team data for the United States. The CDC and OWID data sources define a confirmed case as a person with a laboratory-confirmed case of monkeypox [26, 27]. Data were downloaded every Wednesday evening from the CDC and every Friday afternoon from the GitHub Our World in Data (OWID) repository from the week of July 28th, 2022, through the week of October 13th, 2022. For the week of July 28th, 2022, data posted by the OWID team on August 9th, 2022, was used to produce the forecast as it was the earliest data available.

### The *n*-sub-epidemic modeling framework

A detailed description of our modeling framework is given in ref. [19]. In this n-sub-epidemic modeling framework, epidemic trajectories are modeled as the aggregation of overlapping and asynchronous sub-epidemics. A sub-epidemic follows the 3-parameter generalized-logistic growth model (GLM), which has displayed competitive performance [28­30]. This model is given by the following differential equation:

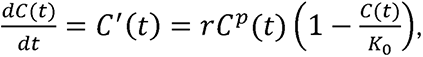

where *C*(*t*) denotes the cumulative number of cases at time t and 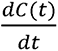 describes the curve of daily cases over time *t*. The parameter *r* is positive, denoting the growth rate per unit of time, *K*_0_ is the final outbreak size, and *p* ∈ [0, 1] is the “scaling of growth” parameter which allows the model to capture early sub-exponential and exponential growth patterns. If *p* = 0, this equation describes a constant number of new cases over time, while *p* = 1 indicates that the early growth phase is exponential. Intermediate values of *p* (0 < *p* < 1) describe early sub-exponential (e.g., polynomial) growth dynamics.

An *n*-sub-epidemic trajectory comprises *n* overlapping sub-epidemics and is given by the following system of coupled differential equations:

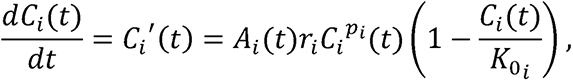

Where *C_i_*(*t*) tracks the cumulative number of cases for sub-epidemic *i*, and the parameters that characterize the shape of the *i_th_* sub-epidemic are given by (*r_i_*, *p_i_*, *K*_0*i*_), for *i* = 1*,…, n.* Thus, the 1-sub-epidemic model is equivalent to the generalized growth model described above. When *n> 1*, we model the onset timing of the (*i* + 1)*_th_* sub-epidemic, where (*i* + 1) ≤ *n*, by employing an indicator variable given by *A_i_*(*t*) so that the (*i* + 1)*_th_* sub-epidemic is triggered when the cumulative curve of the *i_th_* sub-epidemic exceeds *C_thr_*.

The (*i* + 1)*_th_* sub-epidemic is only triggered when *C_thr_ < K*_0_*i*__. Then, we have:

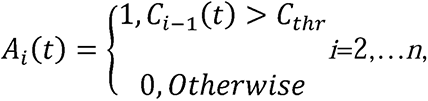

where *A*_1_(*t*) =1 for the first sub-epidemic. Hence, the total number of parameters that are needed to model an *n*-sub-epidemic trajectory is given by 3*n* + 1. The initial number of cases is given by *C*_1_(0) = *I*_0_, where *I*_0_ is the initial number of cases in the observed data. The cumulative curve of the *n*-sub-epidemic trajectory is given by:

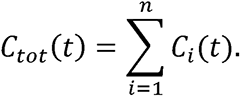

Hence, this modeling framework is suitable for diverse epidemic patterns including those characterized by multiple peaks.

### Parameter estimation for the n-sub-epidemic model

The time series of new weekly monkeypox cases are denoted by:

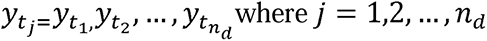

Here, *t_j_* are the time points for the time series data, *n_d_* is the number of observations. Using these case series, we estimate a total of *3n* +1 model parameters, namely *Θ* = (*C_thr_, r*_1_, *r*_1_, *K*_0_1__, …,*r_n_,p_n_*, *K*_0_*n*__). Let *f*(*t, Θ*) denote the expected curve of new monkeypox cases of the epidemic’s trajectory. We can estimate model parameters by fitting the model solution to the observed data via nonlinear least squares [31] or via maximum likelihood estimation assuming a specific error structure [32]. For nonlinear least squares, this is achieved by searching for the set of parameters *Θ* that minimizes the sum of squared differences between the observed data *y_t_j__*=*y*_*t*_1__,*y*_*t*_2__ …. *y_t_n_d___* and the model mean ƒ(*t*,*Θ*). That is, *Θ = (C_thr_, r*_1_, *p*_1_, *K*_0_1__,…,*r_n_,p_n_*,*K*_0_*n*__) is estimated by 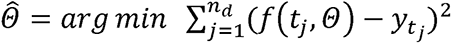.

We quantify parameter uncertainty using a bootstrapping approach described in [33], which allows the computation of standard errors and related statistics in the absence of closed-form solutions. To that end, we use the best-fit model *f*(*t, Θ*) to generate *B*-times replicated simulated datasets of size *n_d_*, where the observation at time *t_j_* is sampled from a normal distribution with mean 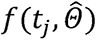 and variance 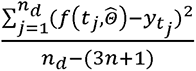. Then, we refit the model to each *B* simulated dataset to re-estimate each parameter. The new parameter estimates for each realization are denoted by *Θ̂*_*b*_ where *b* = 1,2,…,*B*. Using the sets of re-estimated parameters (*Θ̂*_*b*_), the empirical distribution of each estimate can be characterized, and the resulting uncertainty around the model fit can similarly be obtained from *f*(*t*, *Θ̂*_1_), *f*(*t*, *Θ̂*_2_), …, *f*(*t*, *Θ̂*_*B*_). We run the calibrated model forward in time to generate short-term forecasts with quantified uncertainty.

### Selecting the top-ranked sub-epidemic models

We used the *AIC_c_* values of the set of best fit models based on one and two subepidemics to select the top-ranked sub-epidemic models. We ranked the models from best to worst according to their *AIC_c_* values, which is given by [34, 35]:

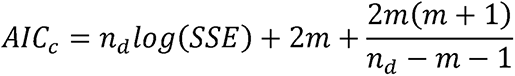

where 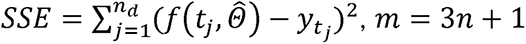 is the number of model parameters, and *n_d_* is the number of data points. Parameters from the above formula for *AIC_c_* are estimated from the nonlinear least-squares fit, which implicitly assumes normal distribution for error.

### Constructing ensemble n-sub-epidemic models

We generate ensemble models from the weighted combination of the highest-ranking sub-epidemic models as deemed by the *AIC_ci_.* for the *i*-th ranked model where *AlC_C_*_1_ *≤* … ≤ *AIC_cI_* and *i* = 1, …, *I.* An ensemble derived from the top-ranking *I* models is denoted by Ensemble(*I*). Thus, Ensemble(2) refers to the ensemble model generated from the weighted combination of the top-ranking 2 sub-epidemic models. We compute the weight *w_t_* for the *i*-th model, *i* = 1, …, *I*, where *Σ_wı_* = 1 as follows:

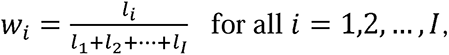

where *l_i_*is the relative likelihood of model *i*, which is given by 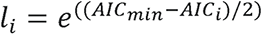 [36], and hence *w_I_ ≤···≤ W*_1_. The prediction intervals based on the ensemble model can be obtained using a bootstrap approach similar as before. We employed the first-ranked and the second-ranked models to derive the ensemble forecasts. *AIC_c_* values of the top models for the most recent forecast can be found in figure 1s (Additional file 1) [24, 25].

### Forecasting strategy

Using a 10-week calibration period for each model, we have conducted 324 real-time weekly sequential 4-week ahead forecasts across studied areas and models (week of July 28^th^ - week of October 13^th^, 2022) thus far. At the national and global levels, we also report forecasting performance metrics for 8 sequential forecasting periods covering the weeks of July 28th, 2022, through September 15, 2022, for which data was available to assess the 4-weeks ahead forecasts. We also compare the predicted cumulative cases for the 4-week forecasts across models for a given setting. Cumulative cases for a given model were calculated as the sum of median number of new cases predicted during the 4-week forecast. Forecasts were evaluated using data reported during the week of October 13^th^, 2022.

### Performance metrics

Across geographic areas, we assessed the quality of our model fit and performance of the short-term forecasts for each model by using four standard performance metrics: the mean squared error (MSE) [37], the mean absolute error (MAE) [38], the coverage of the 95% prediction interval (PI) [37], and the weighted interval score (WIS) [39, 20]. While MSE and

## Results

### Performance of forecasts

Table 1 summarizes the mean forecasting performance metrics of the models by geographic area. Figures 2s-10s (Additional file 1), and tables 1s-4s (Additional file 2) show the forecasting performance metrics for each of the 8 sequential forecasts. Regarding the average MAE, MSE, and WIS across forecasting periods, the first-ranked and weighted ensemble models outperformed the other models for most studied areas except for France, the US (OWID), and the global level (Table 1; Additional file 2: Table 5s). The individual MAE, MSE, and WIS values for each of the eight sequential forecasts evaluated are shown in tables 1s, 2s, and 4s, respectively (Additional file 2).

**Table 1.**
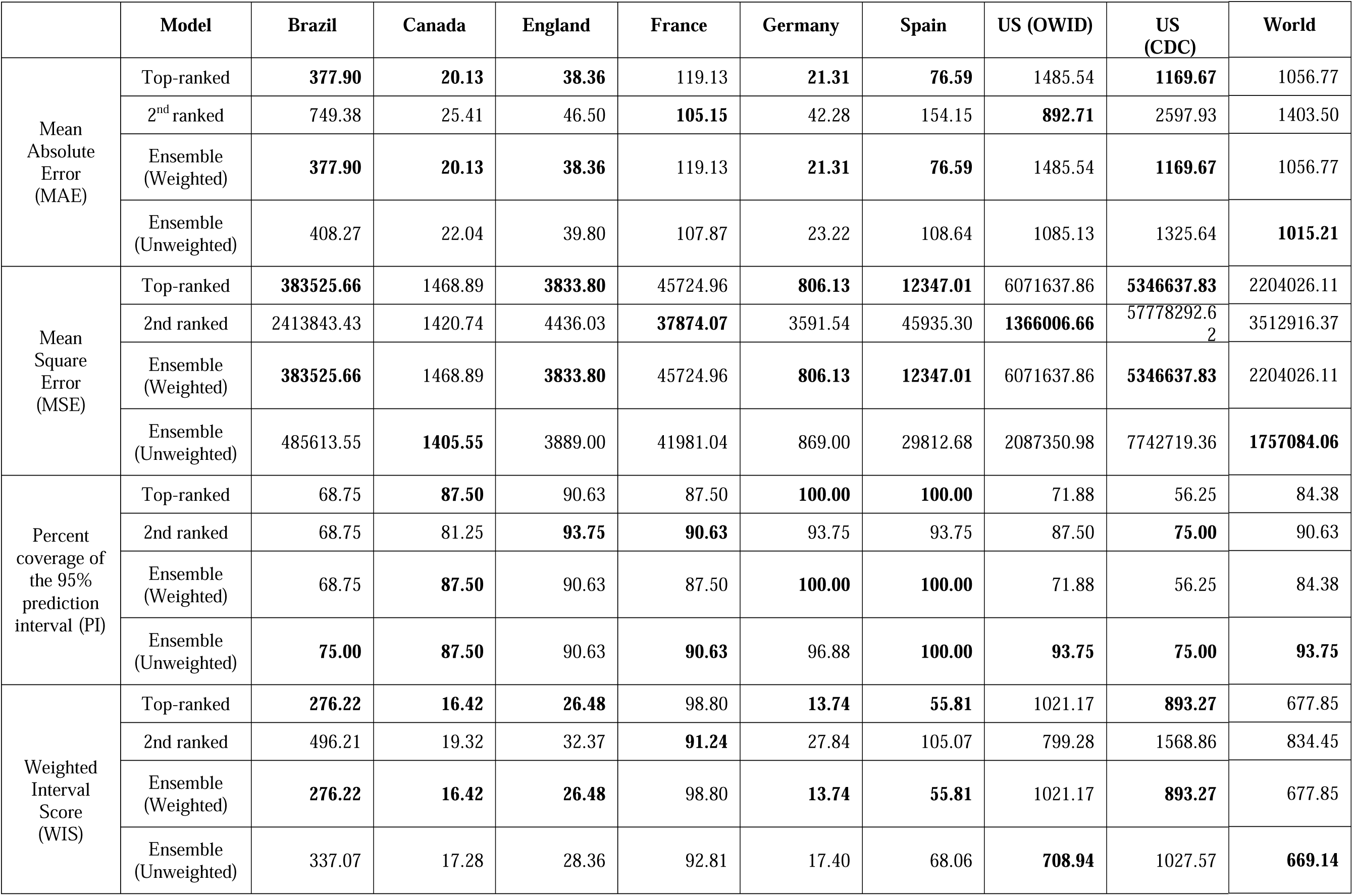
Mean performance metrics of the forecasts generated by the sub-epidemic models. The mean performance metrics of the forecasts generated by the sub-epidemic models’ performance across 8 sequential forecasting periods (Week of July 28^th^ through the week of September 15^th^, 2022) for each geographical area. Only weeks in which observed case data was available are included. Values highlighted in bold correspond to best performing model for a given geographical region and metric.

Regarding the 95% PI coverage, most models had similar coverage value across the 8 sequential forecasts. For example, in Canada, France, England, and Germany, all four models had the same 95% PI coverage in 7 of 8 forecasts, with lower coverage value during early forecasting periods (25% to 75%) and higher coverage value during later forecasting periods (100%) (Additional file 2: Table 3s). Across 72 forecasts involving all study areas (2 datasets for the U.S.), 54.17% (39/72) of the 95% PI coverage was 100% for all four models. In general, the 95% PI coverage improved during the declining phase compared to the earlier periods (Additional file 1: Figs. 2s-10s). Figures 1-9 show the forecasts for the weighted ensemble model for each study area while the forecasts from the top-ranked and second-ranked models are given in supplementary figures (Additional file 1: Figs. 11s-28s).

**Figure 1.**
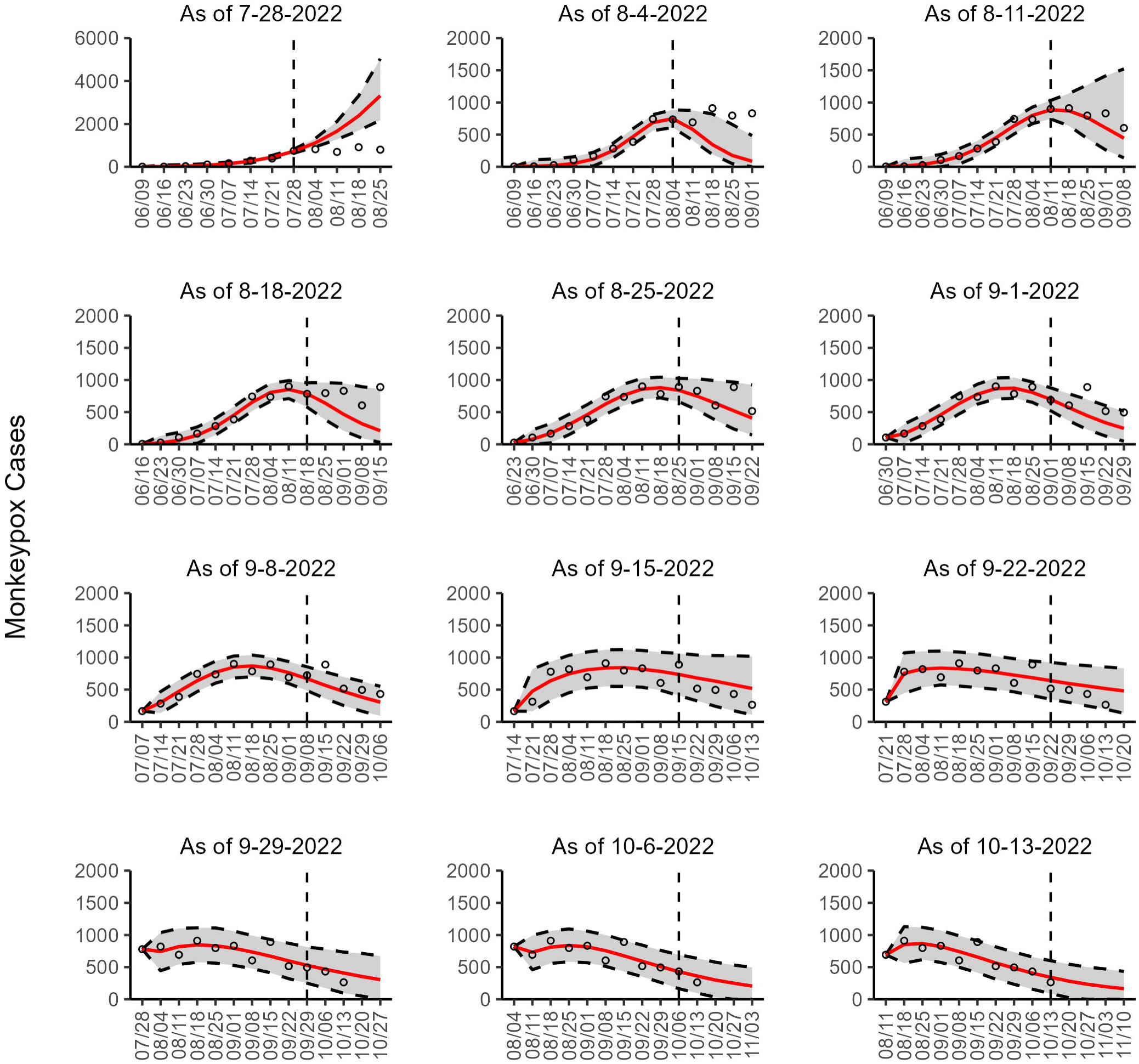
The overlayed forecasted and reported monkeypox cases for the weeks of 7/28/2022 through the week of 10/13/2022 for Brazil. The forecasts are derived from the weighted ensemble sub-epidemic model using 10-week calibration data, and the reported cases are obtained from the OWID GitHub [25]. The black circles to the left of the vertical line represent the reported cases as of the Friday of the forecast period; the solid red line corresponds to the best fit model; the dashed black lines correspond to the 95% prediction intervals. The black circles to the right of the vertical line represent the reported case counts (as of 10/21/2022) for the corresponding date. The vertical dashed black line indicates the start of the forecast period. For the week of 7/28/2022, data posted by the OWID team on 8/9/2022 was used to produce the forecast as it was the earliest version of data available.

**Figure 2.**
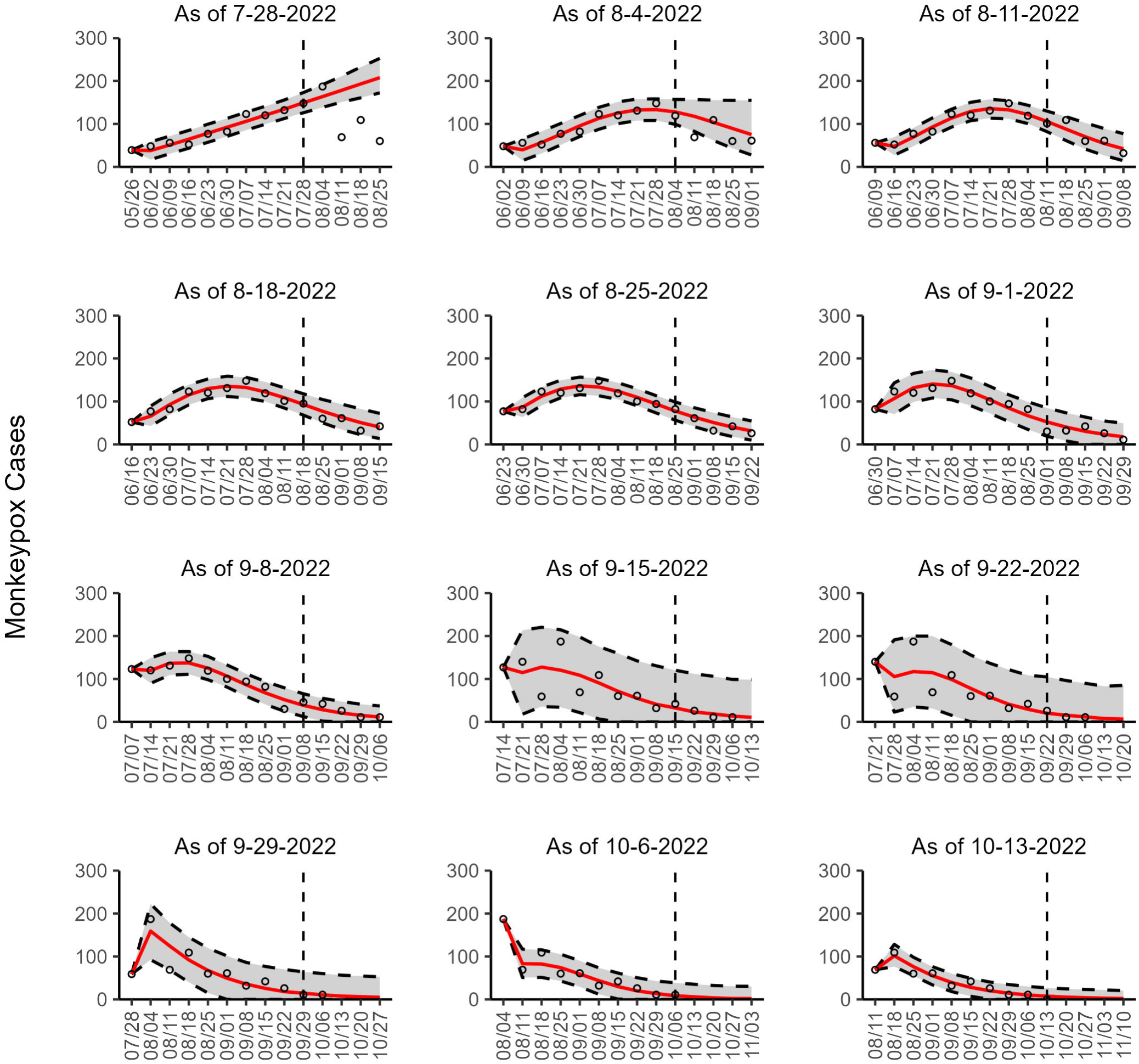
The overlayed forecasted and reported monkeypox cases for the weeks of 7/28/2022 through the week of 10/13/2022 for Canada. The forecasts are derived from the weighted ensemble sub-epidemic model using 10-week calibration data, and the reported cases are obtained from the OWID GitHub [25]. The black circles to the left of the vertical line represent the reported cases as of the Friday of the forecast period; the solid red line corresponds to the best fit model; the dashed black lines correspond to the 95% prediction intervals. The black circles to the right of the vertical line represent the reported case counts (as of 10/21/2022) for the corresponding date. The vertical dashed black line indicates the start of the forecast period. For the week of 7/28/2022, data posted by the OWID team on 8/9/2022 was used to produce the forecast as it was the earliest version of data available.

**Figure 3.**
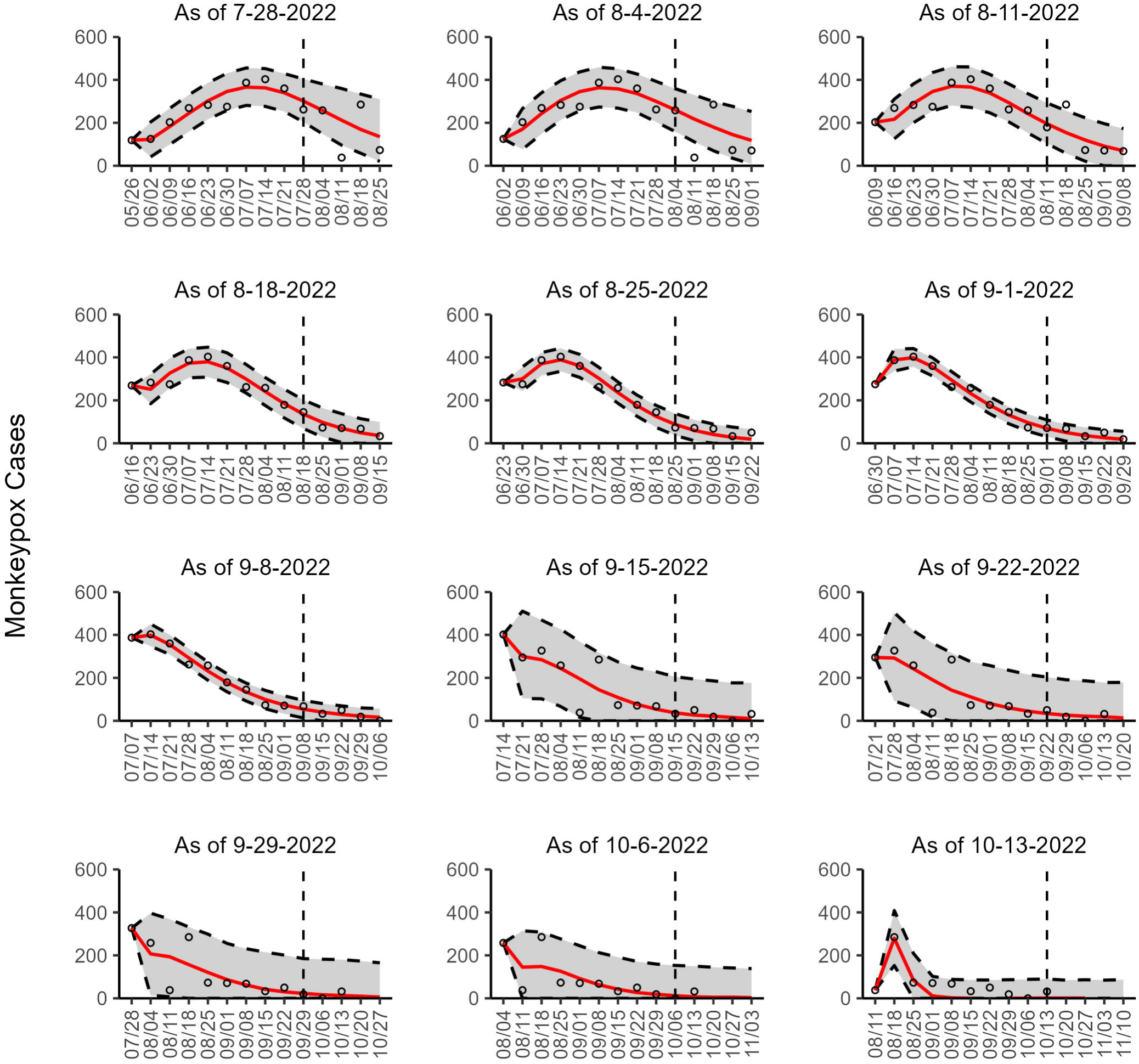
The overlayed forecasted and reported monkeypox cases for the weeks of 7/28/2022 through the week of 10/13/2022 for England. The forecasts are derived from the weighted ensemble sub-epidemic model using 10-week calibration data, and the reported cases are obtained from the OWID GitHub [25]. The black circles to the left of the vertical line represent the reported cases as of the Friday of the forecast period; the solid red line corresponds to the best fit model; the dashed black lines correspond to the 95% prediction intervals. The black circles to the right of the vertical line represent the reported case counts (as of 10/21/2022) for the corresponding date. The vertical dashed black line indicates the start of the forecast period. For the week of 7/28/2022, data posted by the OWID team on 8/9/2022 was used to produce the forecast as it was the earliest version of data available.

**Figure 4.**
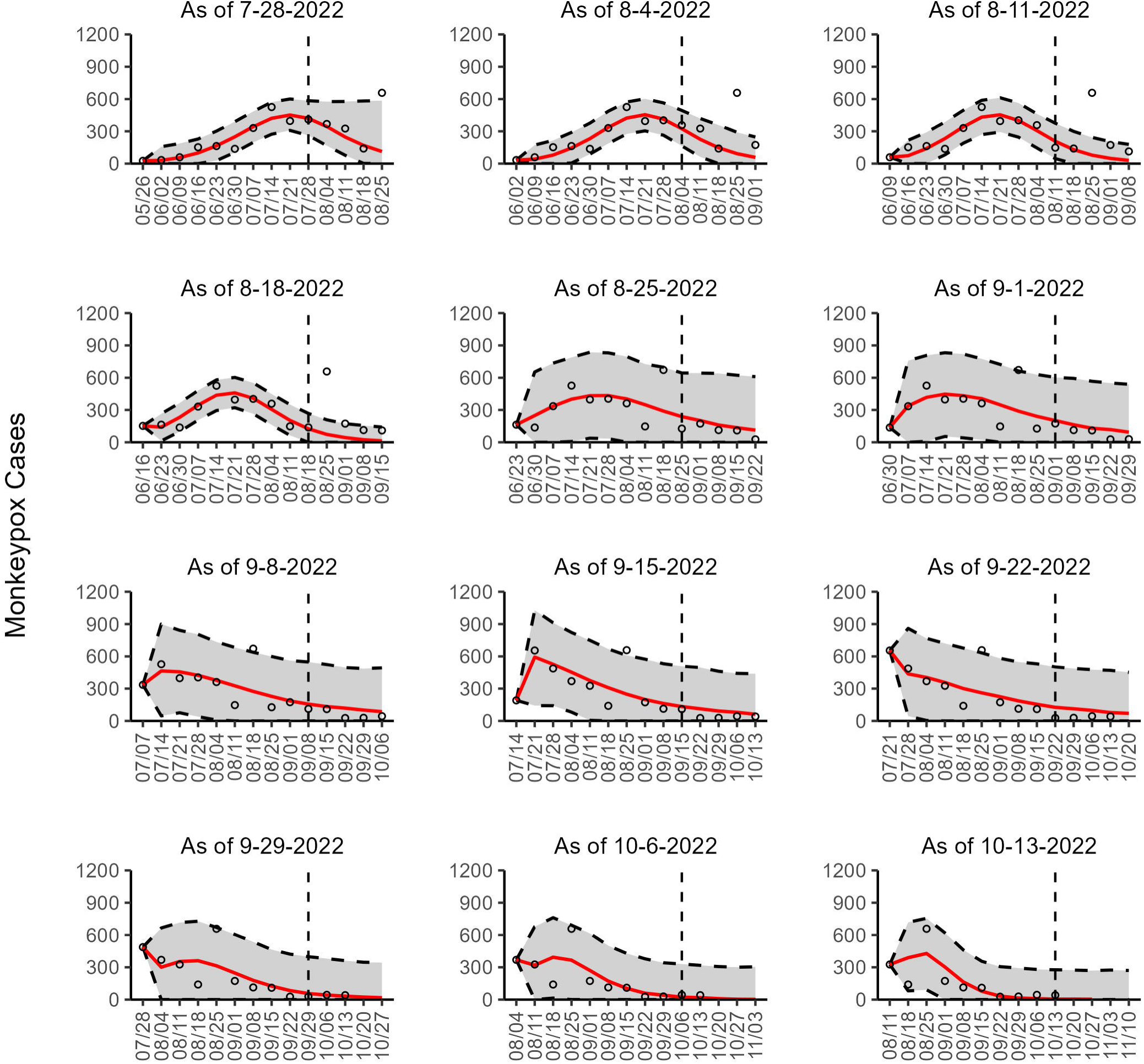
The overlayed forecasted and reported monkeypox cases for the weeks of 7/28/2022 through the week of 10/13/2022 for France. The forecasts are derived from the weighted ensemble sub-epidemic model using 10-week calibration data, and the reported cases are obtained from the OWID GitHub [25]. The black circles to the left of the vertical line represent the reported cases as of the Friday of the forecast period; the solid red line corresponds to the best fit model; the dashed black lines correspond to the 95% prediction intervals. The black circles to the right of the vertical line represent the reported case counts (as of 10/21/2022) for the corresponding date. The vertical dashed black line indicates the start of the forecast period. For the week of 7/28/2022, data posted by the OWID team on 8/9/2022 was used to produce the forecast as it was the earliest version of data available.

**Figure 5.**
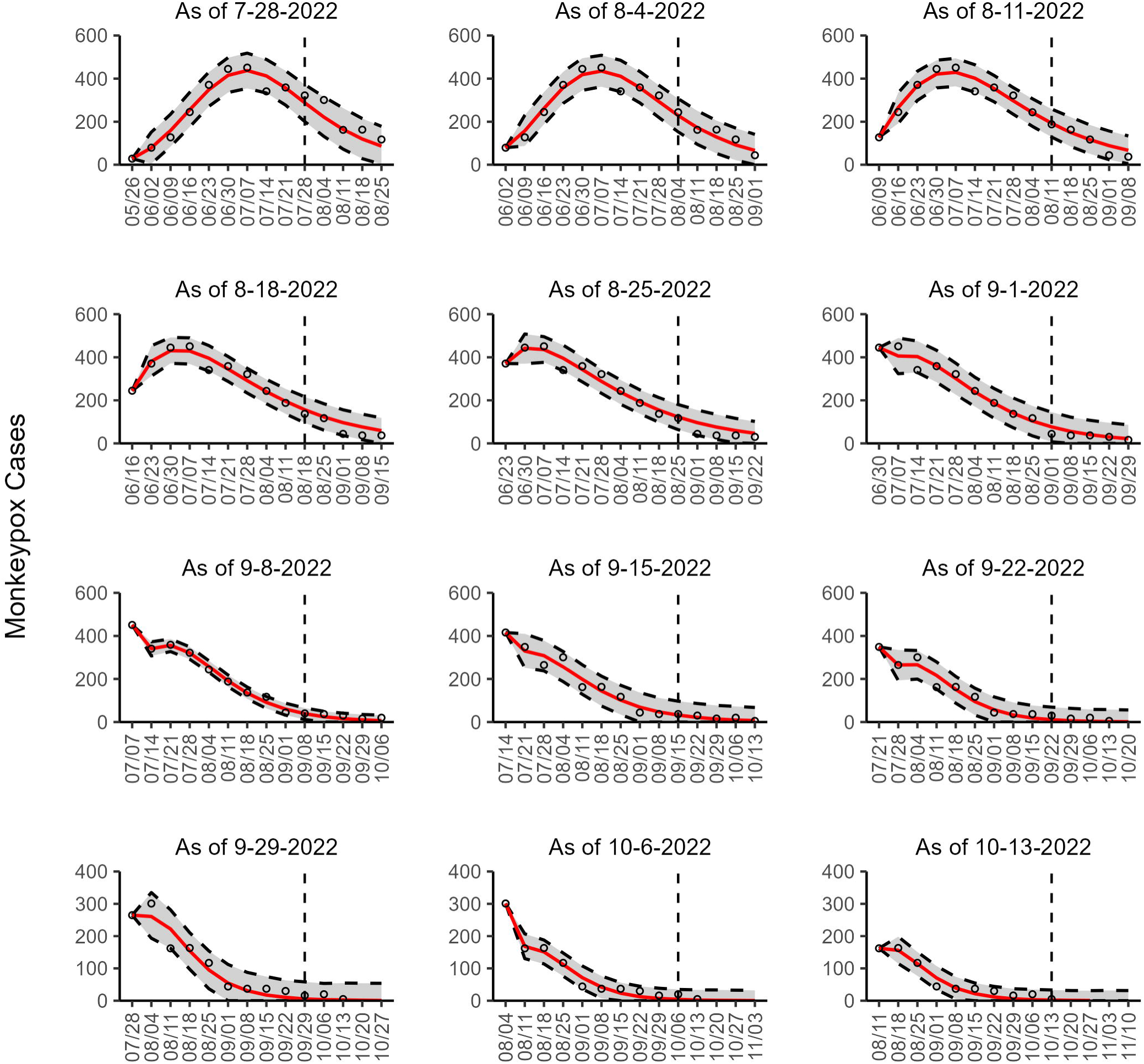
The overlayed forecasted and reported monkeypox cases for the weeks of 7/28/2022 through the week of 10/13/2022 for Germany. The forecasts are derived from the weighted ensemble sub-epidemic model using 10-week calibration data, and the reported cases are obtained from the OWID GitHub [25]. The black circles to the left of the vertical line represent the reported cases as of the Friday of the forecast period; the solid red line corresponds to the best fit model; the dashed black lines correspond to the 95% prediction intervals. The black circles to the right of the vertical line represent the reported case counts (as of 10/21/2022) for the corresponding date. The vertical dashed black line indicates the start of the forecast period. For the week of 7/28/2022, data posted by the OWID team on 8/9/2022 was used to produce the forecast as it was the earliest version of data available.

**Figure 6.**
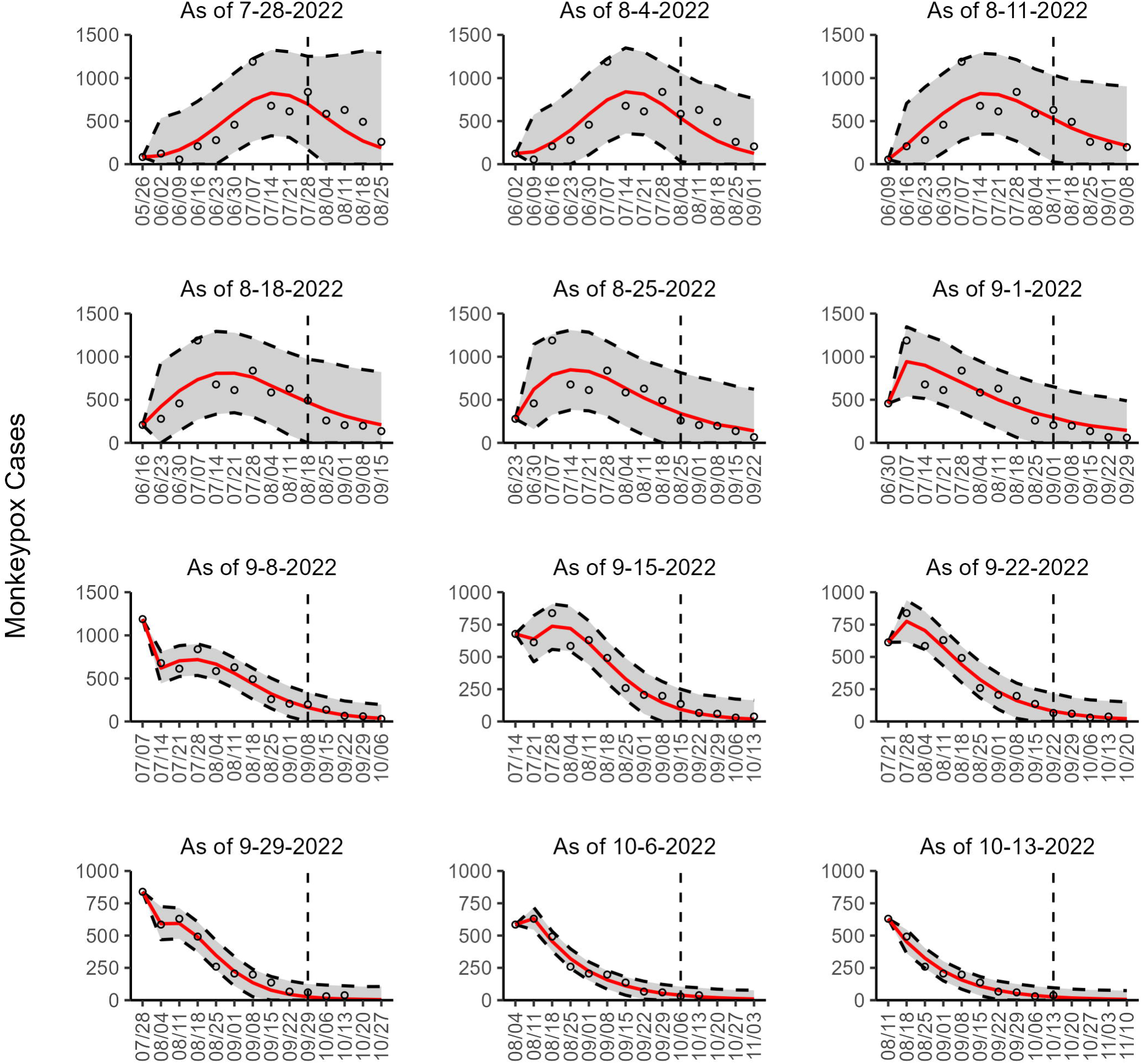
The overlayed forecasted and reported monkeypox cases for the weeks of 7/28/2022 through the week of 10/13/2022 for Spain. The forecasts are derived from the weighted ensemble sub-epidemic model using 10-week calibration data, and the reported cases are obtained from the OWID GitHub [25]. The black circles to the left of the vertical line represent the reported cases as of the Friday of the forecast period; the solid red line corresponds to the best fit model; the dashed black lines correspond to the 95% prediction intervals. The black circles to the right of the vertical line represent the reported case counts (as of 10/21/2022) for the corresponding date. The vertical dashed black line indicates the start of the forecast period. For the week of 7/28/2022, data posted by the OWID team on 8/9/2022 was used to produce the forecast as it was the earliest version of data available.

**Figure 7.**
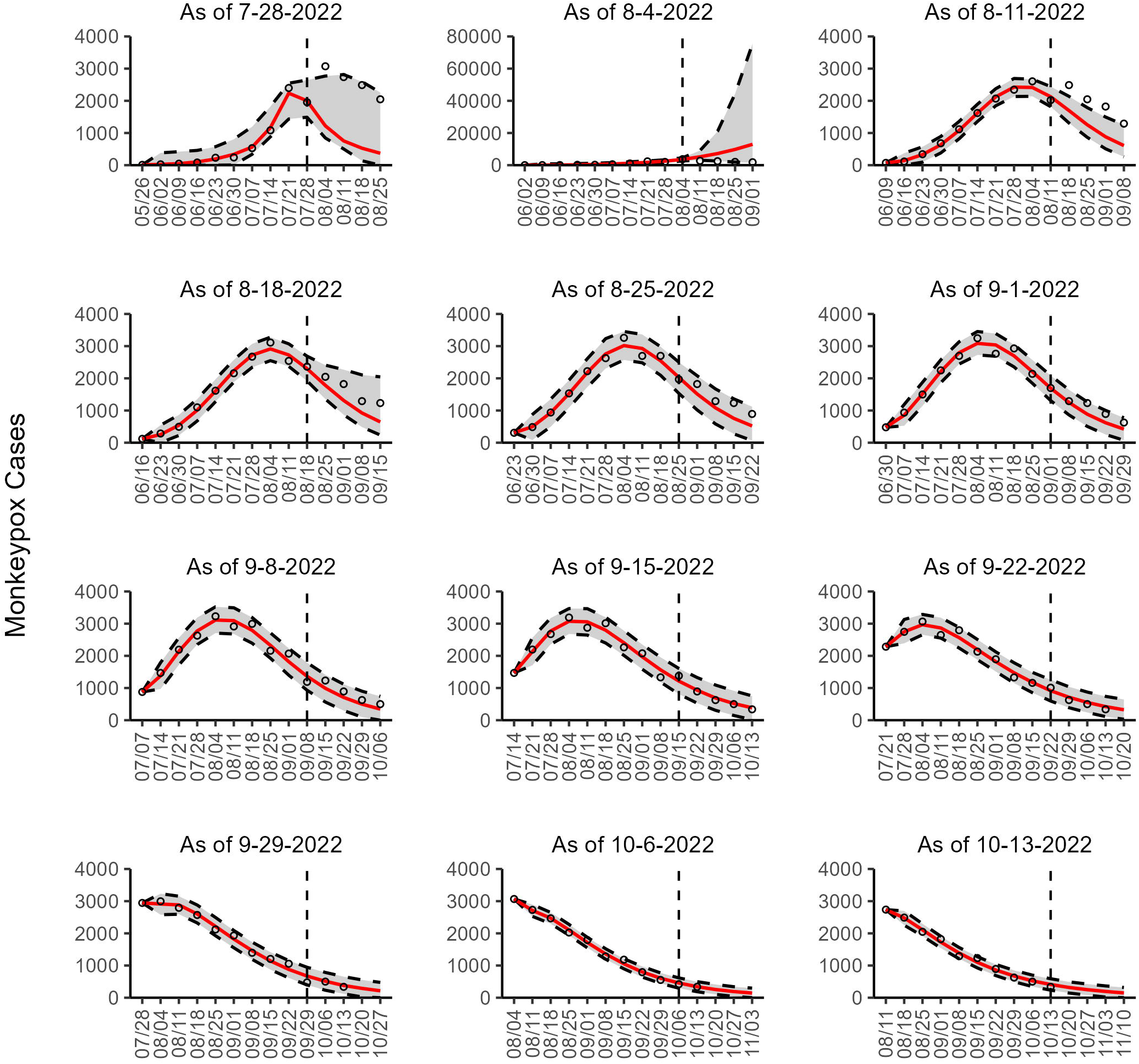
The overlayed forecasted and reported monkeypox cases for the weeks of 7/28/2022 through the week of 10/13/2022 for the United States. The forecasts are derived from the weighted ensemble sub-epidemic model using 10-week calibration data, and the reported cases are obtained from the CDC [24]. The black circles to the left of the vertical line represent the reported cases as of the Wednesday of the forecast period; the solid red line corresponds to the best fit model; the dashed black lines correspond to the 95% prediction intervals. The black circles to the right of the vertical line represent the reported case counts (as of 10/21/2022) for the corresponding date.

**Figure 8.**
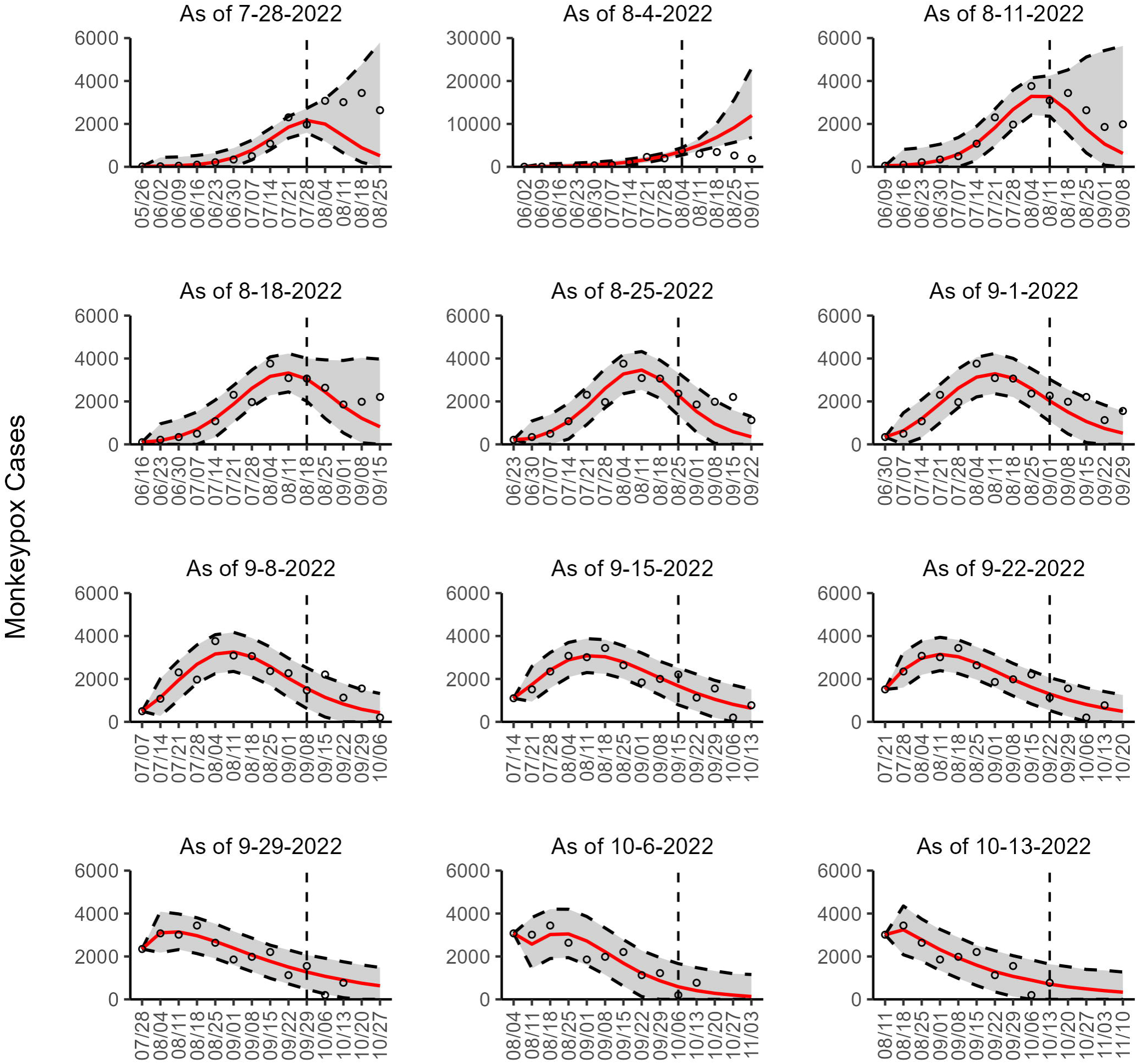
The overlayed forecasted and reported monkeypox cases for the weeks of 7/28/2022 through the week of 10/13/2022 for the United States. The forecasts are derived from the weighted ensemble sub-epidemic model using 10-week calibration data, and the reported cases are obtained from the OWID GitHub [24]. The black circles to the left of the vertical line represent the reported cases as of the Friday of the forecast period; the solid red line corresponds to the best fit model; the dashed black lines correspond to the 95% prediction intervals. The black circles to the right of the vertical line represent the reported case counts (as of 10/21/2022) for the corresponding date. The vertical dashed black line indicates the start of the forecast period. For the week of 7/28/2022, data posted by the OWID team on 8/9/2022 was used to produce the forecast as it was the earliest version of data available.

**Figure 9.**
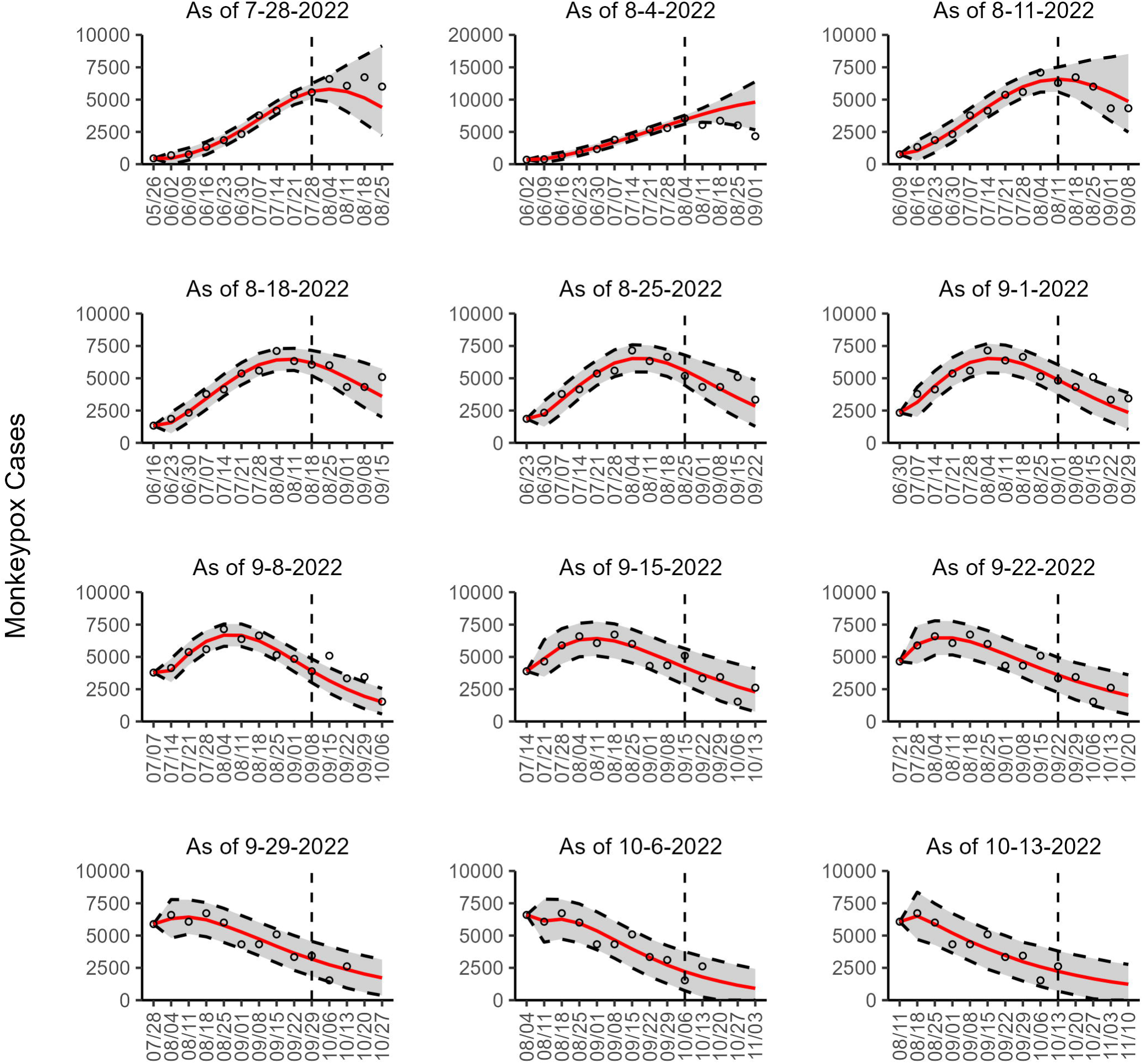
The overlayed forecasted and reported monkeypox cases for the weeks of 7/28/2022 through the week of 10/13/2022 for the World. The forecasts are derived from the weighted ensemble sub-epidemic model using 10-week calibration data, and the reported cases are obtained from the OWID GitHub [25]. The black circles to the left of the vertical line represent the reported cases as of the Friday of the forecast period; the solid red line corresponds to the best fit model; the dashed black lines correspond to the 95% prediction intervals. The black circles to the right of the vertical line represent the reported case counts (as of 10/21/2022) for the corresponding date. The vertical dashed black line indicates the start of the forecast period. For the week of 7/28/2022, data posted by the OWID team on 8/9/2022 was used to produce the forecast as it was the earliest version of data available.

### Performance of the model fit

The mean calibration performance metrics are shown in table 6s (Additional file 2). Overall, the sub-epidemic models yielded good fits for the 10-week calibration periods. In terms of the mean MAE, the second-ranked model performed better than the other models in all the study areas. Likewise, the second-ranked model outperformed other models in terms of mean MSE in all the study areas except Canada. In terms of mean WIS, the unweighted ensemble model performed better than the other 55.5% of the time. The coverage of the 95% PI coverage was consistently 100% across models and study areas except for France, where for the recent two forecast periods, the top-ranked and the weighted ensemble models reached 90% coverage. The performance metrics for each sequential calibration period are shown in tables 7s-10s (Additional file 2) and figures 29s-37s (Additional file 1).

### Global forecasts

Based on the global trend, our latest 4-week ahead forecasts from the three models continue to support a significant slowdown in the growth rate of monkeypox (Figs. 10, 11, 12). Our most recent forecasts for the week of October 13th, 2022, predict a short-term decline in the number of new cases reported worldwide. According to our latest forecasts from the top-ranked model and weighted ensemble model, a total of 6232 (95% PI: 487.8, 12468.0 and 95% PI: 492.8, 12463.1) cases could be added globally during the 4 weeks from October 20th to the week of November 10, 2022 (Table 2; Additional file 2: Table 12s).

**Figure 10.**
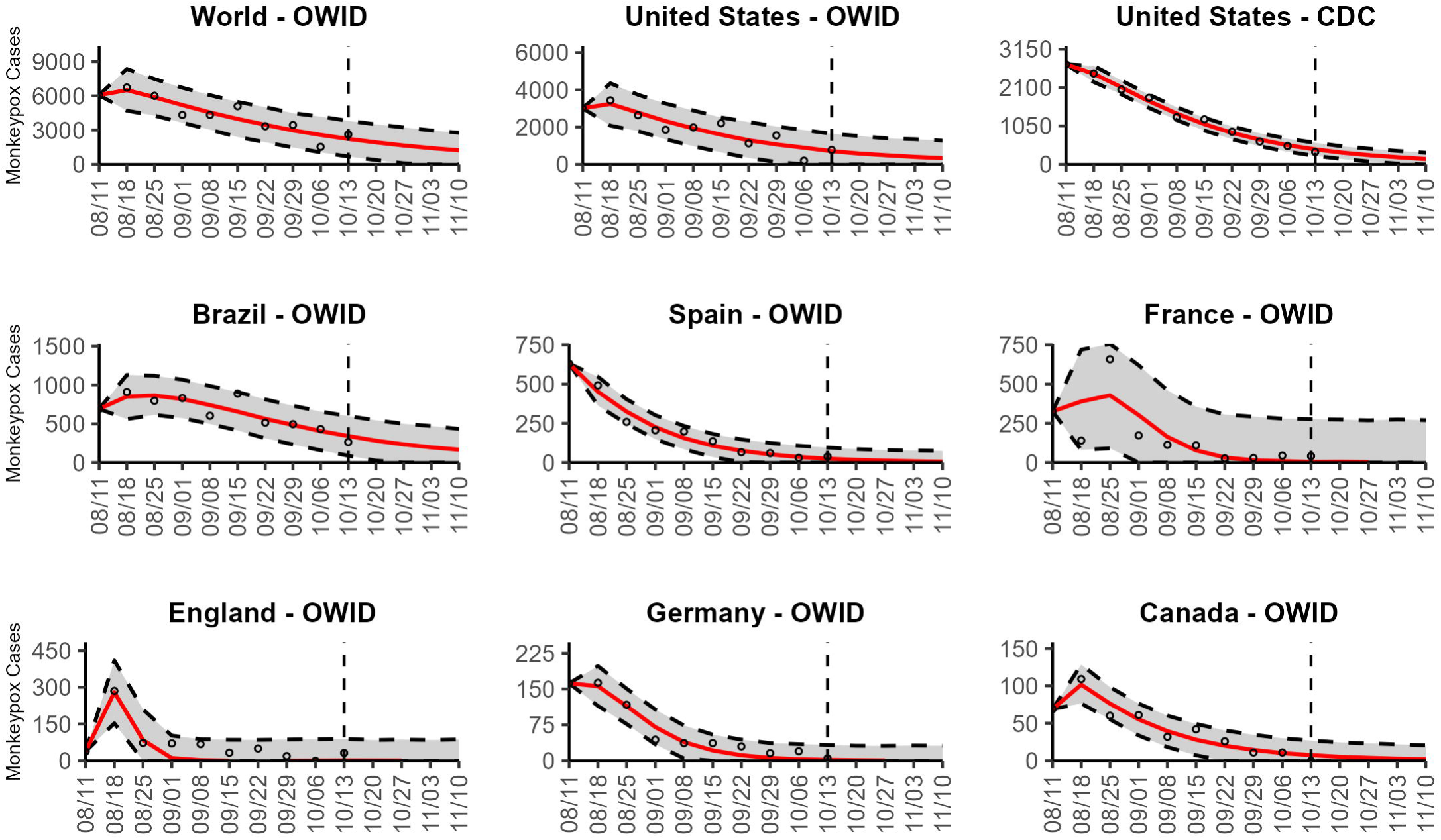
The latest 4-week ahead forecasts of reported monkeypox cases for the week of 10/13/2022 derived from the top-ranked sub-epidemic model based on weekly cases using 10-week calibration data. The black circles to the left of the vertical line represent the reported cases as of the Wednesday (CDC forecast) or Friday (OWID forecast) of the forecast period; the solid red line corresponds to the best fit model; the dashed black lines correspond to the 95% prediction intervals. The vertical dashed black line indicates the start of the forecast period.

**Figure 11.**
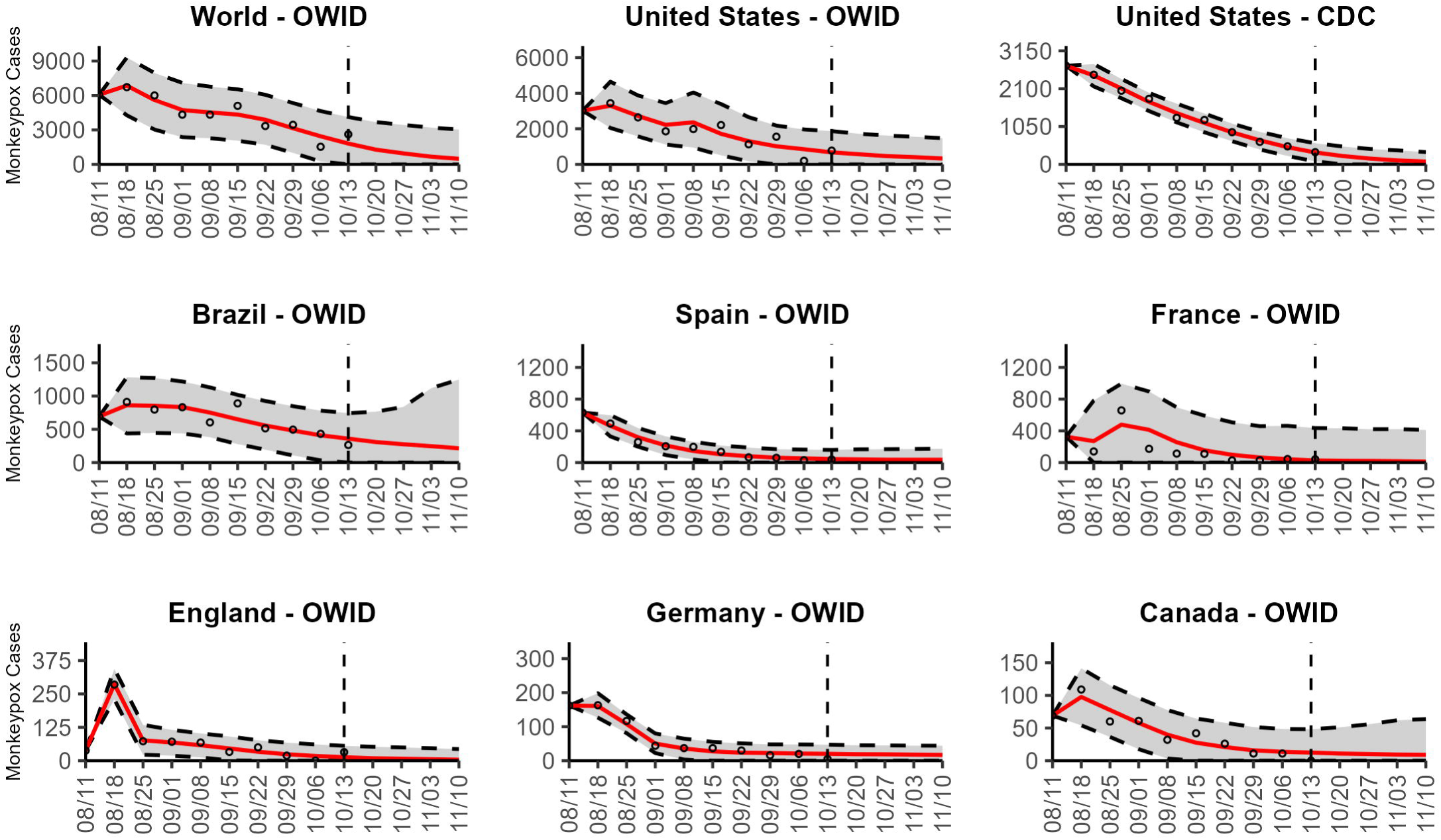
The latest 4-week ahead forecasts of reported monkeypox cases for the week of 10/13/2022 derived from the second-ranked sub-epidemic model based on weekly cases using 10-week calibration data. The black circles to the left of the vertical line represent the reported cases as of the Wednesday (CDC forecast) or Friday (OWID forecast) of the forecast period; the solid red line corresponds to the best fit model; the dashed black lines correspond to the 95% prediction intervals. The vertical dashed black line indicates the start of the forecast period.

**Figure 12.**
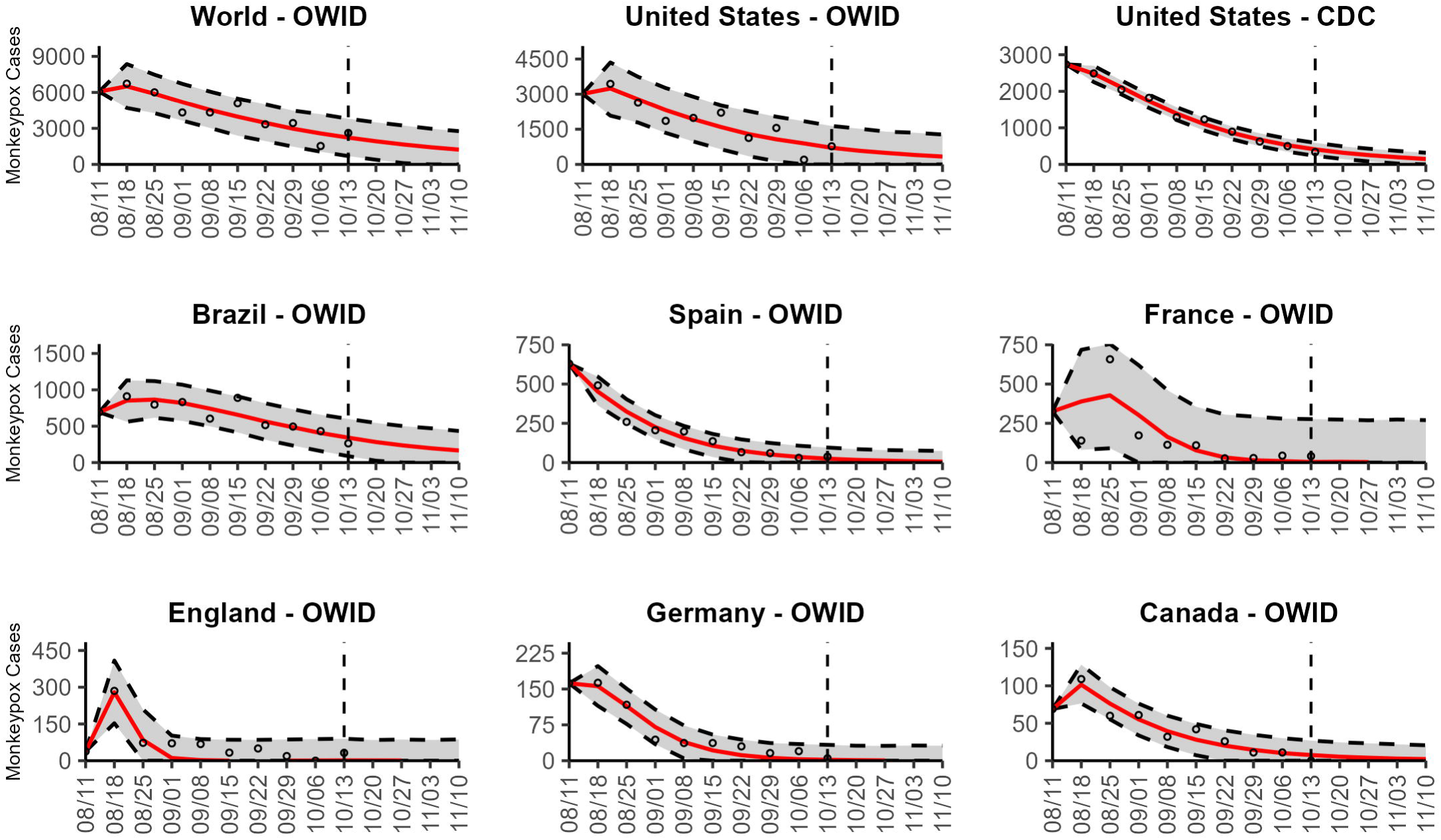
The latest 4-week ahead forecasts of reported monkeypox cases for the week of 10/13/2022 derived from the weighted ensemble sub-epidemic model based on weekly cases using 10-week calibration data. The black circles to the left of the vertical line represent the reported cases as of the Wednesday (CDC forecast) or Friday (OWID forecast) of the forecast period; the solid red line corresponds to the best fit model; the dashed black lines correspond to the 95% prediction intervals. The vertical dashed black line indicates the start of the forecast period.

**Table 2.** Predicted cumulative number of newly forecasted monkeypox cases and 95% PIs for the top-ranked model. Predicted cumulative number of newly reported monkeypox cases and 95% PIs for each 4-week ahead forecasts for the weeks of 7-28-2022 through 10-13-2022 based on a weekly cases using a 10-week calibration period for the top-ranked sub-epidemic model.

| Week |  | Brazil | Canada | England | France | Germany | Spain | US<br>(OWID) | U.S.<br>(CDC) | World |
| --- | --- | --- | --- | --- | --- | --- | --- | --- | --- | --- |
| 07-28-2022 | Median | 8463.7 | 742.3 | 773.9 | 872.4 | 591.2 | 1391.2 | 4838.8 | 3972.2 | 20957.3 |
|  | LB | 6067.5 | 622.3 | 322.9 | 245.6 | 234.1 | 0.0 | 2038.2 | 1482.7 | 14263.4 |
|  | UB | 11783.6 | 887.6 | 1384.3 | 2322.0 | 972.4 | 5137.4 | 17645.5 | 14737.1 | 32121.5 |
| 08-04-2022 | Median | 1187.2 | 385.9 | 660.3 | 522.6 | 458.7 | 966.3 | 33042.3 | 31740.8 | 34936.8 |
|  | LB | 651.6 | 215.0 | 224.7 | 62.0 | 167.6 | 0.0 | 21005.3 | 20265.5 | 24148.0 |
|  | UB | 2841.9 | 621.8 | 1157.7 | 1316.6 | 767.2 | 3435.6 | 55553.7 | 52503.6 | 42706.4 |
| 08-11-2022 | Median | 2685.3 | 254.5 | 434.9 | 286.1 | 420.3 | 1237.7 | 6058.4 | 4301.5 | 22930.0 |
|  | LB | 1477.3 | 143.3 | 79.0 | 0.0 | 162.3 | 0.0 | 2242.7 | 3118.5 | 15153.2 |
|  | UB | 5348.1 | 385.7 | 848.3 | 924.7 | 696.3 | 3754.3 | 20689.6 | 5592.6 | 32717.9 |
| 08-18-2022 | Median | 1631.6 | 231.2 | 251.2 | 152.8 | 354.1 | 1158.6 | 6247.6 | 4539.4 | 18643.0 |
|  | LB | 711.3 | 120.6 | 32.6 | 0.0 | 114.4 | 0.0 | 1816.2 | 3226.2 | 12818.4 |
|  | UB | 3662.6 | 352.7 | 515.4 | 677.8 | 593.4 | 3503.7 | 15853.4 | 6067.3 | 25447.7 |
| 08-25-2022 | Median | 2312.1 | 185.7 | 147.6 | 609.1 | 277.7 | 806.8 | 3420.0 | 3756.9 | 15412.5 |
|  | LB | 1288.4 | 97.3 | 11.7 | 0.0 | 57.8 | 0.0 | 610.6 | 2218.4 | 9456.3 |
|  | UB | 3903.2 | 277.4 | 340.1 | 2517.0 | 506.4 | 2753.8 | 7594.1 | 5342.4 | 22458.5 |
| 09-01-2022 | Median | 1593.3 | 111.7 | 130.1 | 503.9 | 147.1 | 755.3 | 3824.4 | 3175.9 | 12899.5 |
|  | LB | 768.7 | 8.0 | 9.9 | 0.0 | 0.0 | 0.0 | 571.0 | 1697.9 | 7794.2 |
|  | UB | 2599.4 | 240.7 | 281.3 | 2252.0 | 410.0 | 2166.7 | 8101.4 | 4689.0 | 18540.1 |
| 09-08-2022 | Median | 1723.7 | 75.6 | 108.4 | 438.6 | 56.7 | 271.0 | 2978.7 | 2542.4 | 9097.0 |
|  | LB | 866.6 | 1.9 | 0.0 | 0.0 | 0.0 | 0.0 | 223.6 | 959.6 | 5279.6 |
|  | UB | 2648.9 | 180.9 | 267.2 | 1998.6 | 158.0 | 933.0 | 6706.9 | 4176.0 | 13323.9 |
| 09-15-2022 | Median | 2401.9 | 66.0 | 74.0 | 346.6 | 51.6 | 143.9 | 3826.9 | 2526.1 | 11764.0 |
|  | LB | 844.3 | 0.0 | 0.0 | 0.0 | 0.0 | 0.0 | 690.5 | 999.5 | 5704.9 |
|  | UB | 4116.9 | 415.6 | 733.7 | 1836.3 | 303.1 | 739.6 | 7379.2 | 4086.6 | 18545.3 |
| 09-22-2022 | Median | 2144.9 | 41.7 | 79.9 | 357.9 | 12.8 | 143.3 | 2958.6 | 2011.8 | 10205.4 |
|  | LB | 875.0 | 0.0 | 0.0 | 0.0 | 0.0 | 0.0 | 265.3 | 768.4 | 4305.2 |
|  | UB | 3452.4 | 354.2 | 734.6 | 1882.9 | 236.1 | 663.5 | 6078.7 | 3299.8 | 16332.0 |
| 09-29-2022 | Median | 1534.7 | 29.5 | 45.0 | 115.6 | 7.7 | 32.3 | 3365.1 | 1395.1 | 8853.4 |
|  | LB | 320.1 | 0.0 | 0.0 | 0.0 | 0.0 | 0.0 | 326.2 | 377.7 | 3312.8 |
|  | UB | 2898.0 | 223.8 | 701.1 | 1432.3 | 215.9 | 437.5 | 6758.5 | 2469.3 | 14514.1 |
| 10-06-2022 | Median | 1117.8 | 13.6 | 20.0 | 32.7 | 4.3 | 66.1 | 1017.4 | 931.4 | 5297.9 |
|  | LB | 129.8 | 0.0 | 0.0 | 0.0 | 0.0 | 0.0 | 0.0 | 330.9 | 244.2 |
|  | UB | 2240.8 | 129.0 | 574.3 | 1229.4 | 131.4 | 335.6 | 5154.0 | 1551.8 | 11508.3 |
| 10-13-2022 | Median | 878.9 | 14.2 | 2.7 | 10.3 | 1.5 | 43.0 | 1806.3 | 910.1 | 6231.5 |
|  | LB | 25.0 | 0.0 | 0.0 | 0.0 | 0.0 | 0.0 | 0.0 | 245.3 | 487.8 |
|  | UB | 1951.5 | 90.4 | 342.8 | 1088.0 | 125.7 | 315.6 | 5544.5 | 1603.5 | 12468.0 |

The uncertainty of the cumulative predicted median cases was higher for the second-ranked model compared to the top-ranked and ensemble models for most forecasts. For example, for Brazil, the 95% prediction interval for the cumulative median forecasts generated during the week of August 25, 2022, was 1288 to 3903 for the top-ranked model and ensemble model compared to 1038 to 27926 for the second-ranked model (Additional file 1: Figs. 41s, Table 2, Additional file 2: Table 11s). The cumulative predicted median number of cases for each of the models across 12 sequential forecasting weeks for all study areas are shown in table 2 (top-ranked model), table 11s (Additional file 2) (second-ranked model), table 12s (Additional file 2) (weighted ensemble model), table 13s (Additional file 2) (unweighted ensemble model) and figures 38s-46s (Additional file 1).

### Country-level forecasts

Our most recent forecasts from the top-ranked, second-ranked, and the corresponding ensemble sub-epidemic model support a continued declining trend for each of the seven countries (Figs. 10, 12). However, the upper 95% PI of the forecast for Brazil derived from the second-ranked sub-epidemic model, albeit with substantially reduced statistical support, indicates the possibility of a further increasing trend in the number of new cases starting late October 2022 (Fig. 11). Out of a total of 324 forecasts generated from the week of July 28^th^ through the week of October 13^th^ 2022, for all study areas across the three models, 306 (94.4%) forecasts showed a declining trend of monkeypox cases (Figs. 1-9; Additional file 1: Figs.11s-28s).

The United States exhibited a slow increase in cases from late May 2022 until late June 2022, followed by a rapid increase, peaking during early to mid-August 2022. According to our latest forecasts from the weighted ensemble model, a maximum of 1604 and 554 cases could be added from the week of October 20th to the week of November 10, 2022, in the U.S. based on the CDC data and the OWID data, respectively (Additional file 2: Table 12s; Additional file 1: Figs. 39s, 40s). The uncertainty of the predictions based on OWID data is higher than those based on the CDC data (Figs. 10, 11, 12; Additional file 1: Figs. 30s, 40s).

The epidemic curve peaked earliest in Germany in late June 2022. Two weeks later, in mid-July, the epidemic peaked in England. However, the epidemics in the U.S. and Brazil seem to have peaked later, during early August and mid-August, respectively. For Brazil, the top-ranked model predicted the possibility of adding a median of 879 cases (95% PI: 25, 1952) during the 4 weeks until November 10, 2022 (Table 2; Additional file 1: Fig. 41s). Similarly, based on the top-ranked model, Germany and England could each add a median 1.5 (95% PI: 0, 126) and 2.7 (95% PI: 0, 343) cases, respectively, between the week of October 20 to November 10, 2022 (Table 2; Additional file 1: Figs. 42s, 43s). Likewise, for France, Spain, and Canada, the number of cases peaked during the week of July 14-July 21, 2022, followed by a decline in the number of reported new cases. During the most recent 4-week ahead forecast, the top-ranked model predicted very few cases (in terms of median value) of monkeypox in France (median:10, 95% PI: 0, 1088), Spain (median: 43, 95% PI:0, 316), and Canada (median:14, 95% PI: 0, 90) (Table 2; Additional file 1: Figs. 44s, 45s, 46s).

It is reassuring that the cumulative number of new cases predicted for the weeks of September 29th, October 6th, and October 13th, 2022, show a declining pattern for Germany, France, and England, indicating a significant reduction in the growth rate (Table 2; Additional file 1: Figs. 42s-44s). For Brazil, the pattern in the number of cumulative cases in 4-week ahead predictions has consistently declined from the week of September 15 to the week of October 13 (from 2402 to 879, 63.4% decline) (Table 2; Additional file 1: Fig. 41s). For the forecasts generated during the last three weeks (September 29th, October 6th, and October 13th), the predicted median cumulative cases remain below 50 for Canada and England, below 10 for Germany, and below 70 for Spain, suggesting that the trend is stabilizing at low case numbers (Table 2; Additional file 1: Figs. 42s, 43s, 45s, 46s). Table 2, tables 11s-13s (Additional file 2), and figures 38s-46s (Additional file 1) summarize the forecasting results for each model. Our forecast for the United States can be found in our GitHub repository [40] and on our webpage [41].

## Discussion

We report results from short-term (4 weeks ahead) forecasts of monkeypox cases using a sub-epidemic modeling framework for the world and seven countries that, at the time this study began, had reported the great majority of cases. Our forecasts continue to support an overall declining trend in the number of new cases of monkeypox at the global and country-specific levels. Based on the top-ranked model and weighted ensemble model, we predict that during the next four weeks (the week of October 20th, 2022, through the week of November 10, 2022), a total 6232 (95% PI: 487.8, 12468.0, and 95% PI: 492.8, 12463.1) cases of monkeypox could be added globally. At the country level, our top-ranked model indicates that the highest number of new cases will be reported in the United States (OWID data) (median: 1806, 95% PI: 0, 5545) followed by Brazil (median: 879, 95% PI: 25, 1952) and Spain (median: 43, 95% PI: 0, 316).

Overall, our models have performed reasonably well across study areas. The top-ranked and weighted ensemble model outperformed other models on average in forecasting performance. Our results offer valuable information to policymakers to guide the continued allocation of resources and inform mitigation efforts. More broadly, findings suggest that the epidemic could be brought under near-complete control in some regions should public health measures continue to be sustained, especially among the high-risk groups [42–44]. Indeed, a core group of higher-risk people is thought to disproportionately contribute to transmission and thereby sustain sexually transmitted infection (STI) epidemics. Monkeypox is inherently different from other STIs like HIV, which has a lifelong duration, or bacterial STIs, which can be acquired repeatedly. Cases may decline rapidly as immunity increases among core group members, either due to infection or vaccination. Without a core group driving the epidemic, monkeypox may become endemic with low transmission levels [45–47]. The current monkeypox outbreak is unprecedented in size and geographic scope. As of November 23rd, 2022, a total of 110 countries globally have reported monkeypox cases at 80,899 [8]. Only seven countries have historically reported monkeypox cases indicating that more than 93% of the countries reporting cases are non-endemic to monkeypox [8]. Our latest short-term forecasts from top-ranked models and weighted ensemble models conducted in near real-time indicate a clear, continued slow-down in the number of new cases globally and in each country included in the study. This mirrors the recent continental declines in cases reported for Europe and the Americas [43, 48, 49].

Findings support the significant impact of current measures to contain the outbreak in different areas. For example, in the United States, the primary strategy has been a combination of increasing education around monkeypox (e.g., symptoms, transmission), encouraging practices that reduce potential close contacts and increasing access to vaccination and testing for high-risk groups [50–52]. Although supply-chain shortages impacted early vaccine access, as of August 26th, 2022, the availability of monkeypox vaccines had increased to sufficient levels to combat the outbreak. The racial disparities in access to vaccines that arose in late August are continuing to persist [53–55] though some progress has been made in improving the monkeypox vaccination among racial and ethnic minority groups in the US. For example, according to a recent morbidity and mortality weekly report (MMWR), between May 22–June 25 and July 31–October 10, 2022, the proportion of monkeypox vaccine recipients increased from 15% to 23% among Hispanic and from 6% to 13% among Black population [56]. In addition, although vaccines are more recently available, behavioral modification within high-risk groups appears to be driving declines in cases. Continuing these behaviors (e.g., limiting one-time sexual encounters) is crucial in slowing the transmission of monkeypox [5, 42, 57]. Based on the early evidence from Europe, the World Health Organization (WHO) is quite optimistic that the current outbreak of monkeypox can be contained with the improvement in vaccine supply chains in addition to early detection of the cases and educational interventions which lead to behavioral modifications in the high-risk groups [58]. Moreover, recent research indicates that the majority of monkeypox cases resulting in severe disease or death have been among MSM with compromised immune systems (e.g., due to untreated HIV infection). Specifically, increased burden has been noted among Black populations, and those experiencing mental health challenges or housing insecurities, reflecting the existing inequities in access to resources diagnosis, treatment, and prevention of monkeypox [59].

Our study is not exempt from limitations. First, our analysis relied on weekly time series data of lab-confirmed monkeypox cases from two sources, which display irregular daily reporting patterns [24, 25]. These sources use different approaches in compiling data and addressing data issues, which affect the characterization of the epidemic curve. For example, the CDC data uses cases with reporting data that includes either the positive laboratory test report date, CDC call center reporting date, or case data entry date into CDC’s emergency response common operating platform [24]. The OWID team uses laboratory-confirmed case data reported to the World Health Organization via WHO Member States [25, 60]. In addition, the data used for forecasting could also be underestimated due to delays between the date of testing and the date of reporting. Also, the weekly data used in our real-time forecasts has exhibited revisions that retrospectively adjusted the time series. Hence, significant increases or decreases may be observed in reported cases for the same date between forecasting periods. Indeed, the CDC acknowledged the presence of data adjustments within their posted data [61]. Similar issues have been noted in COVID-19 forecasting studies since ground truth data adjustments occurred during the pandemic [20]. Nevertheless, in the case of our study and the COVID-19 forecasting study, forecasts are being conducted in real-time using ground truth data. Therefore, each weekly forecast is generated using the latest time series available on each prediction date whereas forecasts were scored using the most up-to-date data at the time of the study (week of October 13^th^, 2022). Because we are dealing with limited epidemic data in this study, we often examined forecasts derived from the second-ranked sub-epidemic model even when it yielded substantially diminished statistical support relative to the top-ranked model. The models employed in this study are semi-mechanistic in that they give insight into the nature of the process that generated the epidemic trends in terms of the aggregation of sub-epidemic trajectories. However, the models are not intended to quantify the effects of different factors, such as behavior change and vaccination, on the declining trend. Finally, it should be noted that our short-term forecasts are based on the inherent assumption that current behavior practices will not change substantially, at least over short time horizons. Further, our models are not sensitive to long-term forecasting episodic risk behaviors that are seasonal or event specific (e.g., LGBTQ Pride festivals). For example, a previous study has reported episodic risk behaviors among MSM, such as condomless anal sex with new male sex partners while vacationing [62].

In future work, we plan to systematically assess the forecasting performance of the models against other competing models, such as the Autoregressive Integrated Moving Average (ARIMA), which has been broadly applied to forecast time series of epidemics and various other phenomena such as the weather and the stock market [19, 63, 64]. During the COVID-19, this sub-epidemic modeling framework demonstrated reliable forecasting performance in 10 to 30-day ahead forecasts of daily deaths, outperforming ARIMA models in weekly short-term forecasts covering the U.S. trajectory of the COVID-19 pandemic from the early phase of spring 2020 to the Omicron-dominated wave [19].

## Conclusions

In summary, real-time forecasting during epidemic emergencies offers actionable information that governments can use to anticipate healthcare needs and strategize the intensity and configuration of public health interventions. Our study provides and assesses near-real time short-term forecasts of monkeypox cases globally and for seven countries that have reported a higher number of cases for a large part of the epidemic. Our models continue to predict a slowdown in the incidence of monkeypox both at the global and country levels. Overall, our models have demonstrated utility in providing short-term forecasts, capturing the growth rate slowdown and peak timing with reasonable accuracy. Our findings likely reflect the impact of increased immunity and behavioral modification among high-risk populations.

## Data Availability

The datasets analyzed during the current study include the U.S. Monkeypox Case Trends Reported to CDC data [https://www.cdc.gov/poxvirus/monkeypox/response/2022/mpx-trends.html] and data available from the Our World in Health Monkeypox GitHub repository [https://github.com/owid/monkeypox]. The forecast generated during the current study are available in the Monkeypox short term Forecasts, USA GitHub repository [https://github.com/gchowell/monkeypox-usa] and the Monkeypox Forecasting Center website [https://publichealth.gsu.edu/research/monkeypox-forecasting-center/].

https://github.com/owid/monkeypox

https://github.com/gchowell/monkeypox-usa

https://publichealth.gsu.edu/research/monkeypox-forecasting-center/

## List of Abbreviations

ARIMA: Autoregressive Integrated Moving Average model
CDC: The Centers for Disease Control and Prevention
OWID: Our World in Data
LGBTQ: Lesbian, Gay, Bisexual, Transgender, and Queer/Questioning
MAE: Mean Absolute Error
mpox: Monkeypox
MSE: Mean Square Error
MSM: Men who have sex with men
PI: Prediction interval
STI: Sexual transmitted illness
WHO: World Health Organization
WIS: Weighted Interval Score

## Declarations

### Ethics approval and consent to participate

Not applicable.

### Consent for publication

Not applicable.

### Competing interest

G.C. is a Board Member for the journal. The other authors declare that they have no competing interests.

### Funding

A.B. and S.D. are supported by a 2CI fellowship from Georgia State University. G.C. is partially supported from NSF grants 1610429 and 1633381 and R01 GM 130900.

### Authors’ contributions

A.B., S.D. and G.C. analyzed the data. A.B. and G.C. retrieved and managed data; A.B., S.D. R.L., K.M., and L.C., and G.C. wrote the first draft of the manuscript. All authors contributed to writing and revising subsequent versions of the manuscript. All authors read and approved the final manuscript.

## Acknowledgements

We would like to acknowledge the GSU Advanced Research Computing Technology & Innovation Core (ARCTIC) team for their help with the development of the monkeypox forecast website.

## Additional Files

Additional File 1

.pdf

Supplementary Figures

Additional figures related to the dissemination and evaluation of models used to produce the forecasts included within the study.

Additional File 2

.pdf

Supplementary Tables

Additional tables related to the dissemination and evaluation of models used to produce the forecasts included within the study.

## Notes

### Competing Interest Statement

The authors have declared no competing interest.

### Summary of Updates

New citations have been added, data quality aspect have been mentioned in limitation and each additonal file in which referenced supplementary figures and tables are located are explicitly cited.

## References

1. Monkeypox cases confirmed in England – latest updates. In: Infectious diseases GOV.UK. 2022. https://www.gov.uk/government/news/monkeypox-cases-confirmed-in-england-latest-updates. Accessed 2 Sep 2022.

2. WHO. Monkeypox - United Kingdom of Great Britain and Northern Ireland. In: Disease Outbreak News. World Health Organization 2022. https://www.who.int/emergencies/disease-outbreak-news/item/2022-DON383. Accessed 2 Sep 2022.

3. Scales A. Massachusetts public health officials confirm case of monkeypox. In: News. Massachusetts Department of Public Health. 2022. https://www.mass.gov/news/massachusetts-public-health-officials-confirm-case-of-monkeypox. Accessed 2 Sep 2022.

4. Philpott D. Epidemiologic and clinical characteristics of monkeypox cases—United States, May 17–July 22, 2022. MMWR Morbidity and Mortality Weekly Report. 2022; 71.

5. Spicknall IH, Pollock ED, Clay PA, Oster AM, Charniga K et al. Modeling the impact of sexual networks in the transmission of Monkeypox virus among gay, bisexual, and other men who have sex with men—United States, 2022. MMWR Morbidity and Mortality Weekly Report. 2022; 71(35):1131–1135.

6. Monkeypox outbreak 2022 – Global. https://www.who.int/emergencies/situations/monkeypox-oubreak-2022. Accessed 29 Oct 2022.

7. Monkeypox declared a global health emergency by the World Health Organization. In: Health U.N. News 2022. https://news.un.org/en/story/2022/07/1123152. Accessed 3 Sep 2022.

8. Global Map & Case Count. https://www.cdc.gov/poxvirus/monkeypox/response/2022/world-map.html. Accessed 28 Nov 2022.

9. WHO. Key facts. In: Monkeypox. World Health Organization. 2022. https://www.who.int/news-room/fact-sheets/detail/monkeypox. Accessed 2 Sep 2022.

10. Poxvirus Diseases. https://www.cdc.gov/poxvirus/diseases.html. Accessed 2 Sep 2022.

11. Past U.S. Cases & Outbreaks. https://www.cdc.gov/poxvirus/monkeypox/outbreak/us-outbreaks.html. Accessed 2 Sep 2022.

12. Mandavilli A. Three Pressing Questions About Monkeypox: Spread, Vaccination, Treatment. In: Monkeypox. The New York Times. 2022. https://www.nytimes.com/2022/07/29/health/monkeypox-spread-vaccine-treatment.html. Accessed 6 Sep 2022.

13. Martínez JI, Montalbán EG, Bueno SJ, Martínez FM, Juliá AN et al. Monkeypox outbreak predominantly affecting men who have sex with men, Madrid, Spain, 26 April to 16 June 2022. Eurosurveillance. 2022; 27(27):2200471.

14. Selb R, Werber D, Falkenhorst G, Steffen G, Lachmann R et al. A shift from travel-associated cases to autochthonous transmission with Berlin as epicentre of the monkeypox outbreak in Germany, May to June 2022. Eurosurveillance. 2022; 27(27):2200499.

15. WHO. Multi-country outbreak of monkeypox, External situation report #4 - 24 August 2022. 4 edn. In: Emergency Situational Updates. World Health Organization. 2022. Accessed 29 Oct 2022.

16. Schmerling RH. What to Know About Monkeypox. In: Education. Harvard Medical School. 2022. https://hms.harvard.edu/news/what-know-about-monkeypox. Accessed 2 Sep 2022.

17. Molteni M, Branswell H, Joseph A, Mast J. 10 key questions about monkeypox the world needs to answer. In: Health. STAT. 2022. https://www.statnews.com/2022/08/30/10-key-questions-about-monkeypox-the-world-needs-to-answer/. Accessed 2 Sep 2022.

18. WHO. Vaccines and immunization for monkeypox: interim guidance, 24 August 2022. World Health Organization. 2022. https://www.who.int/publications/i/item/WHO-MPX-Immunization-2022.2-eng. Accessed 29 Oct 2022.

19. Chowell G, Dahal S, Tariq A, Roosa K, Hyman JM et al. An ensemble n-sub-epidemic modeling framework for short-term forecasting epidemic trajectories: Application to the COVID-19 pandemic in the USA. PLOS Computational Biology. 2022; 18(10):e1010602.

20. Cramer EY, Ray EL, Lopez VK, Bracher J, Brennen A et al. Evaluation of individual and ensemble probabilistic forecasts of COVID-19 mortality in the United States. Proceedings of the National Academy of Sciences. 2022; 119(15):e2113561119.

21. Viboud C, Sun K, Gaffey R, Ajelli M, Fumanelli L et al. The RAPIDD ebola forecasting challenge: Synthesis and lessons learnt. Epidemics. 2018; 22:13–21.

22. Biggerstaff M, Johansson M, Alper D, Brooks LC, Chakraborty P et al. Results from the second year of a collaborative effort to forecast influenza seasons in the United States. Epidemics. 2018; 24:26–33.

23. Chowell G, Tariq A, Hyman JM. A novel sub-epidemic modeling framework for short-term forecasting epidemic waves. BMC medicine. 2019; 17(1):1–18.

24. U.S. Monkeypox Case Trends Reported to CDC. Centers for Disease Control and Prevention. https://www.cdc.gov/poxvirus/monkeypox/response/2022/mpx-trends.html. Accessed 19 Oct 2022.

25. Mathieu E, Peters F, Peters C. Monkeypox Data. https://github.com/owid/monkeypox. Accessed 21 Oct 2022.

26. Case Definitions for Use in the 2022 Monkeypox Response. https://www.cdc.gov/poxvirus/monkeypox/clinicians/case-definition.html. Accessed 29 Oct 2022.

27. WHO. Multi-country monkeypox outbreak in non-endemic countries. In: Disease Outbreak News. World Health Organization. 2022. https://www.who.int/emergencies/disease-outbreak-news/item/2022-DON385. Accessed 2 Sep 2022.

28. Chowell G, Hincapie-Palacio D, Ospina J, Pell B, Tariq A et al. Using phenomenological models to characterize transmissibility and forecast patterns and final burden of Zika epidemics. PLoS currents. 2016; 8.

29. Pell B, Kuang Y, Viboud C, Chowell G. Using phenomenological models for forecasting the 2015 Ebola challenge. Epidemics. 2018; 22:62–70.

30. Shanafelt DW, Jones G, Lima M, Perrings C, Chowell G. Forecasting the 2001 foot-and-mouth disease epidemic in the UK. EcoHealth. 2018; 15(2):338–347.

31. Banks HT, Hu S, Thompson WC. Modeling and inverse problems in the presence of uncertainty. CRC Press; 2014.

32. Roosa K, Luo R, Chowell G. Comparative assessment of parameter estimation methods in the presence of overdispersion: a simulation study. Math Biosci Eng. 2019; 16:4299–4313.

33. Hastie T, Tibshirani R, Friedman JH, Friedman JH. The elements of statistical learning: data mining, inference, and prediction, vol 2. Springer; 2009.

34. Hurvich CM, Tsai C-L. Regression and time series model selection in small samples. Biometrika. 1989; 76(2):297–307.

35. Sugiura N. Further analysts of the data by akaike’s information criterion and the finite corrections: Further analysts of the data by akaike’s. Communications in Statistics-theory and Methods. 1978; 7(1):13–26.

36. Burnham KP, Anderson DR. Model selection and multimodel inference: a practical information-theoretic approach. 2nd ed. New York, NY: Springer-Verlag 2002: p. 488.

37. Gneiting T, Raftery AE. Strictly proper scoring rules, prediction, and estimation. Journal of the American statistical Association. 2007; 102(477):359–378.

38. Kuhn M, Johnson K. Applied predictive modeling, vol 26. Springer; 2013.

39. Bracher J, Ray EL, Gneiting T, Reich NG. Evaluating epidemic forecasts in an interval format. PLoS computational biology. 2021; 17(2):e1008618.

40. Chowell G, Bleichrodt A, Dahal S. Monkeypox short term Forecasts, USA. https://github.com/gchowell/monkeypox-usa/wiki. Accessed 23 Oct 2022.

41. Chowell G, Bleichrodt A, Dahal S. Forecasts of national monkeypox incidence in the United States. https://publichealth.gsu.edu/research/monkeypox-forecasting-center/. Accessed 19 Oct 2022.

42. Delaney KP. Strategies adopted by gay, bisexual, and other men who have sex with men to prevent monkeypox virus transmission—United States, August 2022. MMWR Morbidity and mortality weekly report. 2022; 71.

43. Doucleff M, Huang P. Monkeypox cases in the U.S. are way down - can the virus be eliminated? In: Public Health. NPR. 2022. https://www.npr.org/sections/health-shots/2022/10/17/1129234501/monkeypox-cases-in-the-u-s-are-way-down-can-the-virus-be-eliminated. Accessed 28 Oct 2022.

44. Reardon S. What does the future look like for monkeypox? In: News Feature. nature. 2022. 250–252. https://www.nature.com/articles/d41586-022-03204-7. Accessed 28 Oct 2022.

45. Doherty IA, Padian NS, Marlow C, Aral SO. Determinants and consequences of sexual networks as they affect the spread of sexually transmitted infections. The Journal of infectious diseases. 2005; 191 Suppl 1:S42–S54.

46. Liljeros F, Edling CR, Amaral LAN. Sexual networks: implications for the transmission of sexually transmitted infections. Microbes and infection. 2003; 5(2):189–196.

47. Thomas JC, Tucker MJ. The development and use of the concept of a sexually transmitted disease core. Journal of Infectious Diseases. 1996; 174 Suppl 2:S134–S143.

48. Kupferschmidt K. Monkeypox cases are plummeting. Scientists are debating why. In: All News. Science. 2022. https://www.science.org/content/article/monkeypox-cases-are-plummeting-scientists-are-debating-why. Accessed 29 Oct 2022.

49. Soucheray S. Global monkeypox cases drop 22%. In: Featured News Topics. CIDRAP. 2022. https://www.cidrap.umn.edu/news-perspective/2022/09/global-monkeypox-cases-drop-22. Accessed 28 Oct 2022.

50. FACT SHEET: Biden-Harris Administration’s Monkeypox Outbreak Response. In: Statements and Releases. The White House. 2022. https://www.whitehouse.gov/briefing-room/statements-releases/2022/06/28/fact-sheet-biden-harris-administrations-monkeypox-outbreak-response/. Accessed 28 Aug 2022.

51. Prevention. https://www.cdc.gov/poxvirus/monkeypox/prevention/index.html. Accessed 6 Sep 2022.

52. Huang P. Monkeypox cases are rising in the U.S. What’s being done to stop the outbreak? Edited by Khalid A. In: Health. NPR. 2022. https://www.npr.org/2022/07/29/1114417062/monkeypox-cases-are-rising-in-the-u-s-whats-being-done-to-stop-the-outbreak. Accessed 29 Aug 2022.

53. Kates J, Artiga S, Dawson L. National Data Show Continuing Disparities in Monkeypox (MPX) Cases and Vaccinations Among Black and Hispanic People. In: Racial Equity and Health Policy. KFF. 2022. https://www.kff.org/racial-equity-and-health-policy/issue-brief/national-data-show-continuing-disparities-in-mpx-monkeypox-cases-and-vaccinations-among-black-and-hispanic-people/#:~:text=MPX%20case%20rates%20among%20Black,and%208.3%20per%20100%2C000%2C%20respectively. Accessed 29 Oct 2022.

54. Stobbe M, Johnson CK, Miller Z. US Data Reveals Racial Gaps in Monkeypox Vaccinations. In: Health News. U.S. News 2022. https://www.usnews.com/news/health-news/articles/2022-08-26/monkeypox-vaccine-supply-now-sufficient-biden-officials-say. Accessed 24 Oct 2022.

55. Wortman ZE, Kansagra SM, Tilson EC, Moore Z, Farrington DC, et al. To Drive Equity In Monkeypox Response, States Should Learn From COVID-19. In: Health Affairs Forefront. HealthAffairs. 2022. https://www.healthaffairs.org/content/forefront/drive-equity-monkeypox-response-states-should-draw-lessons-covid-19#:~:text=Health%20Affairs%20Forefront-,To%20Drive%20Equity%20In%20Monkeypox%20Response,Should%20Learn%20From%20COVID%2D19&text=Since%20the%20US%20Department%20of,and%20who%20is%20getting%20vaccinated. Accessed 29 Oct 2022.

56. Kriss JL. Receipt of First and Second Doses of JYNNEOS Vaccine for Prevention of Monkeypox—United States, May 22–October 10, 2022. MMWR Morbidity and Mortality Weekly Report. 2022; 71.

57. Stone W, Huang P. The monkeypox outbreak may be slowing in the U.S., but health officials urge caution. In: Public Health. NPR. 2022. https://www.npr.org/sections/health-shots/2022/08/26/1119659681/early-signs-suggest-monkeypox-may-be-slowing-in-the-us. Accessed 6 Sep 2022.

58. Grover N. Monkeypox outbreak can be eliminated in Europe, WHO says. Edited by Birsel R, Baum B. In: Europe. Reuters. 2022. https://www.reuters.com/world/europe/monkeypox-outbreak-can-be-eliminated-europe-who-officials-2022-08-30/. Accessed 6 Sep 2022.

59. Miller MJ. Severe Monkeypox in Hospitalized Patients—United States, August 10– October 10, 2022. MMWR Morbidity and Mortality Weekly Report. 2022; 71.

60. 2022 Monkeypox Outbreak: Global Trends. https://worldhealthorg.shinyapps.io/mpx_global/#section-fns3. Accessed 29 Oct 2022.

61. Technical report: Multi-national monkeypox outbreak, United States, 2022. In: Monkeypox Technical Reports. vol. 4. 2022 Outbreak Cases & Data: Centers for Disease Control and Prevention; 2022.

62. Elsesser SA, Oldenburg CE, Biello KB, Mimiaga MJ, Safren SA et al. Seasons of risk: anticipated behavior on vacation and interest in episodic antiretroviral pre-exposure prophylaxis (PrEP) among a large national sample of US men who have sex with men (MSM). AIDS and Behavior. 2016; 20(7):1400–1407.

63. Mondal P, Shit L, Goswami S. Study of effectiveness of time series modeling (ARIMA) in forecasting stock prices. International Journal of Computer Science, Engineering and Applications. 2014; 4(2):13.

64. Tektaş M. Weather forecasting using ANFIS and ARIMA models. Environmental Research, Engineering and Management. 2010; 51(1):5–10.

