## Additional file 1 for "Real-time forecasting the trajectory of monkeypox outbreaks at the national and global levels, July – October 2022"

**Additional file 1: Supplementary figures**

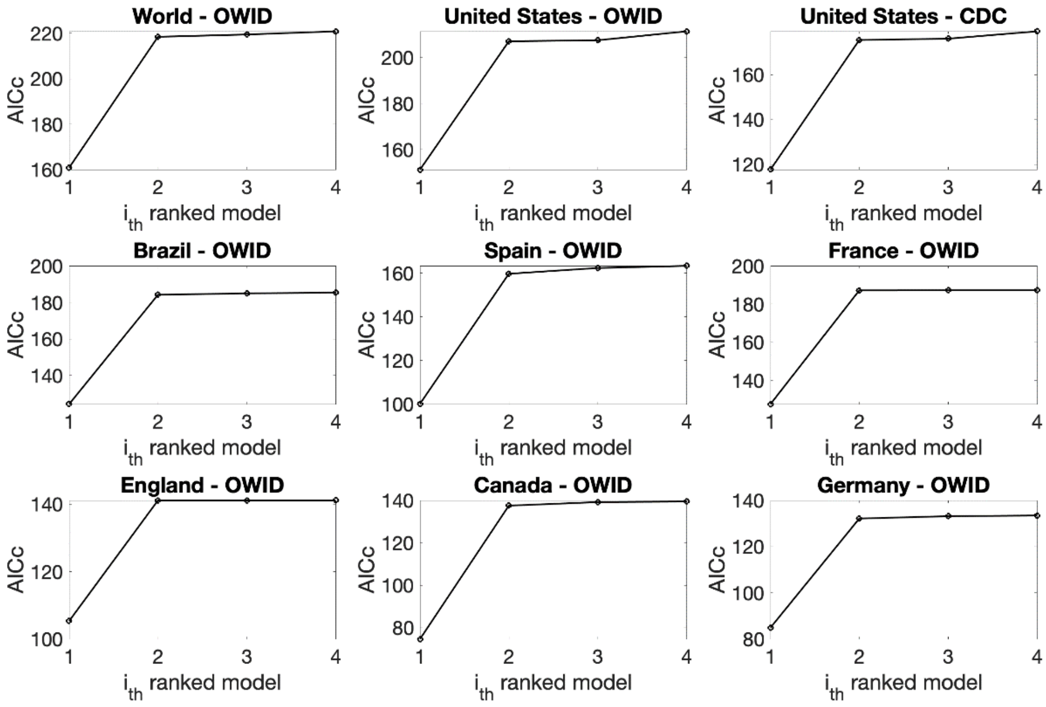


**Figure 1s.** AICc values of the top sub-epidemic models for the latest forecasts produced during the week of October 13th, 2022.

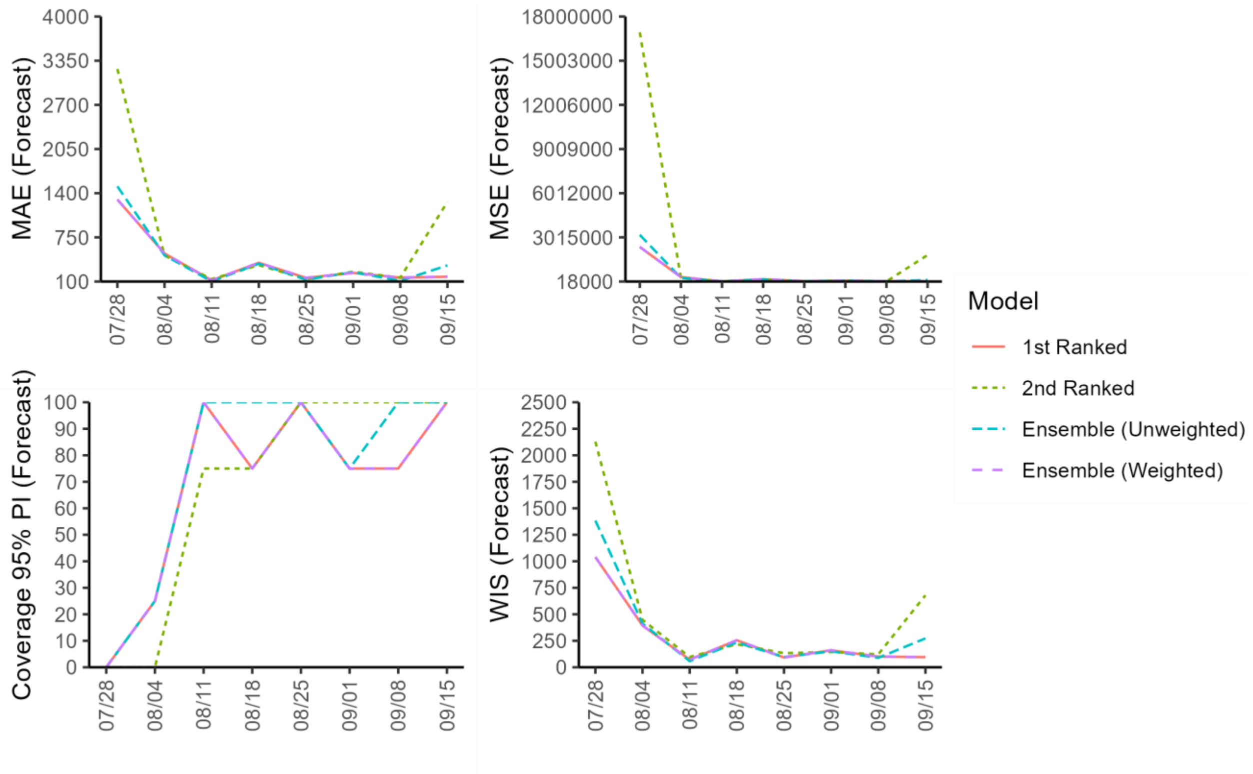


**Figure 2s.** Performance metrics of the forecasts generated by the sub-epidemic models across 8 sequential forecasting periods (Week of July 28$^{th}$ through the week of September 15$^{th}$, 2022) for Brazil.

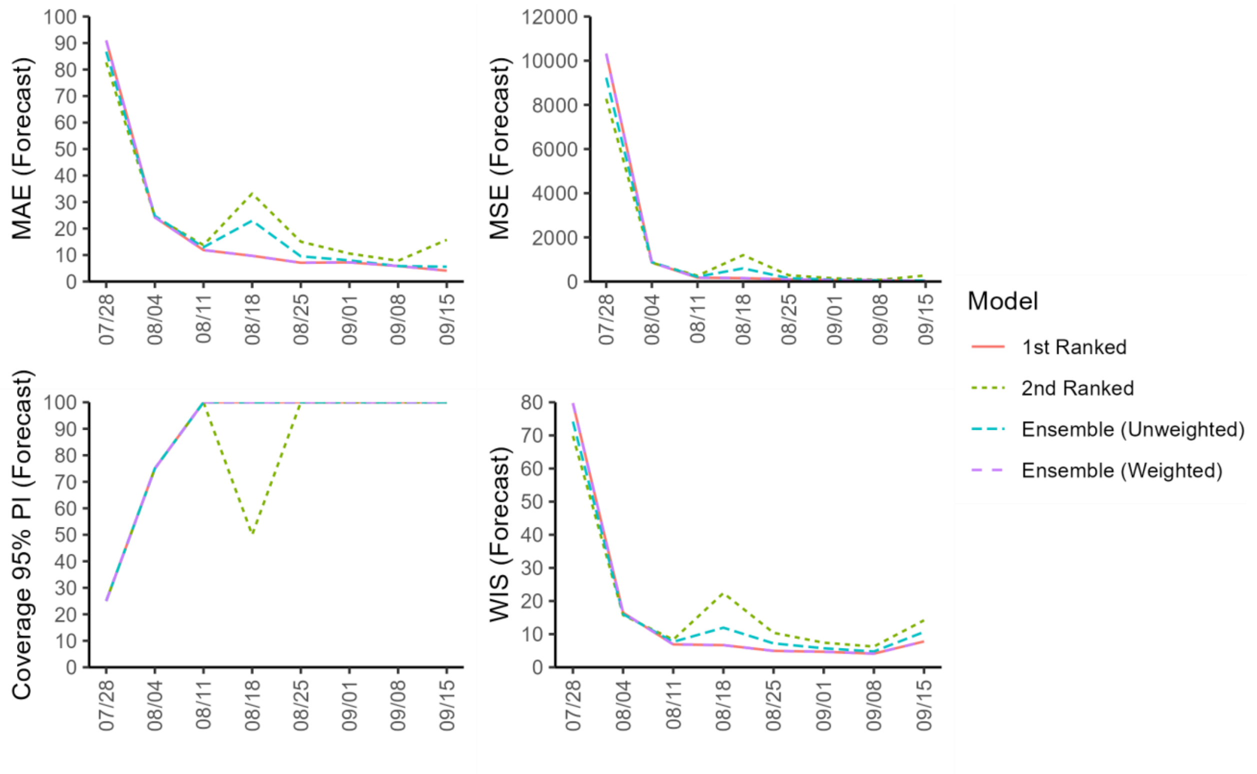


**Figure 3s.** Performance metrics of the forecasts generated by the sub-epidemic models across 8 sequential forecasting periods (Week of July 28th through the week of September 15th, 2022) for Canada.

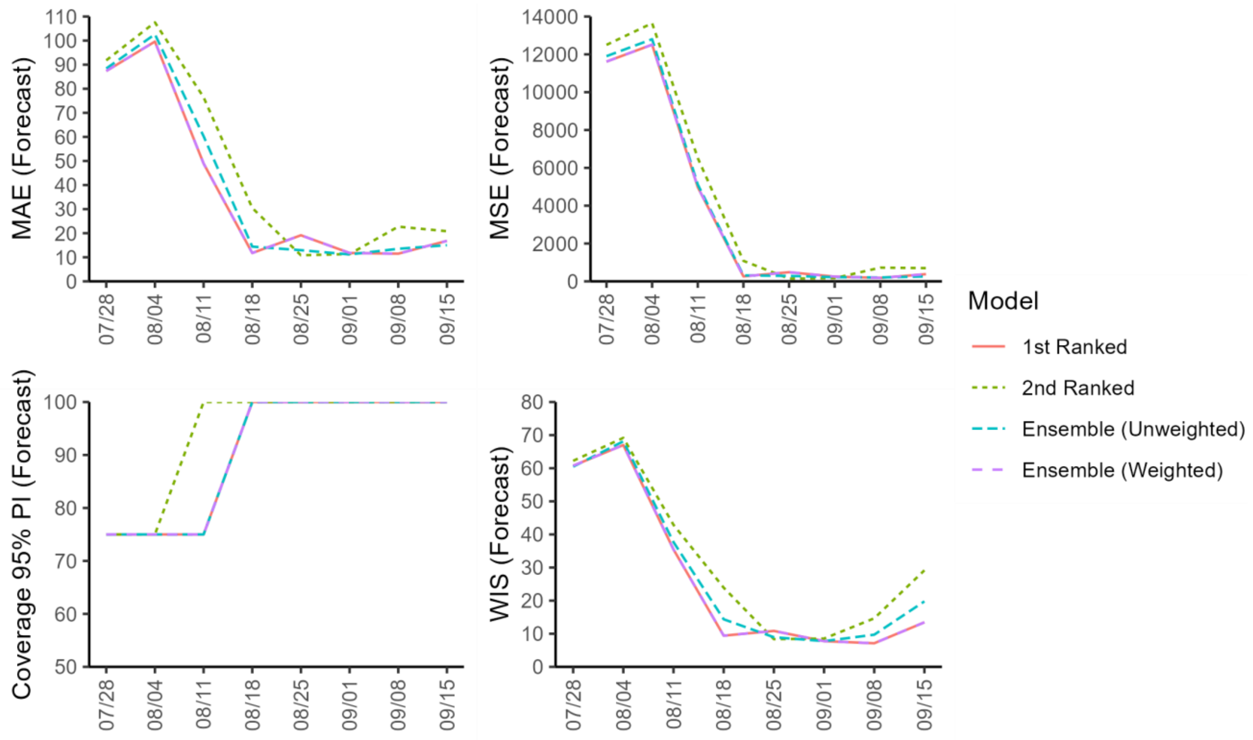


**Figure 4s.** Performance metrics of the forecasts generated by the sub-epidemic models across 8 sequential forecasting periods (Week of July 28th through the week of September 15th, 2022) for England.

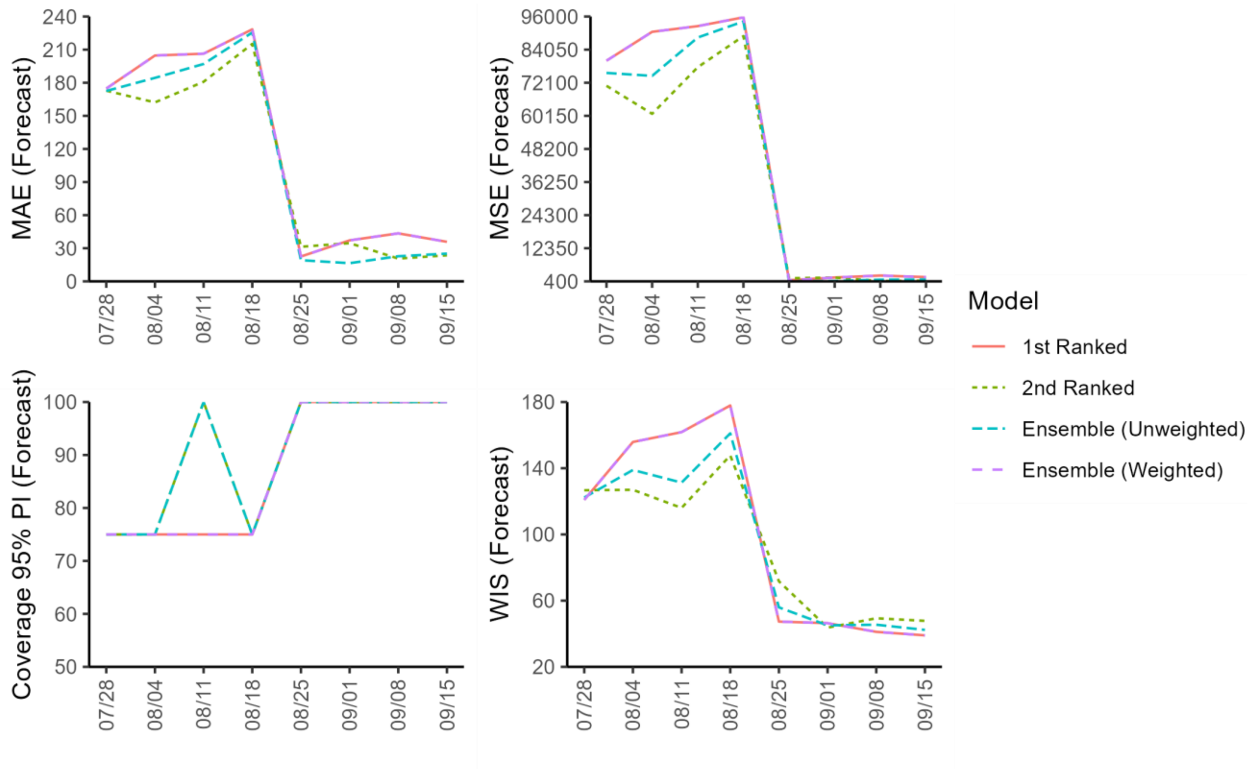


**Figure 5s.** Performance metrics of the forecasts generated by the sub-epidemic models across 8 sequential forecasting periods (Week of July 28th through the week of September 15th, 2022) for France.

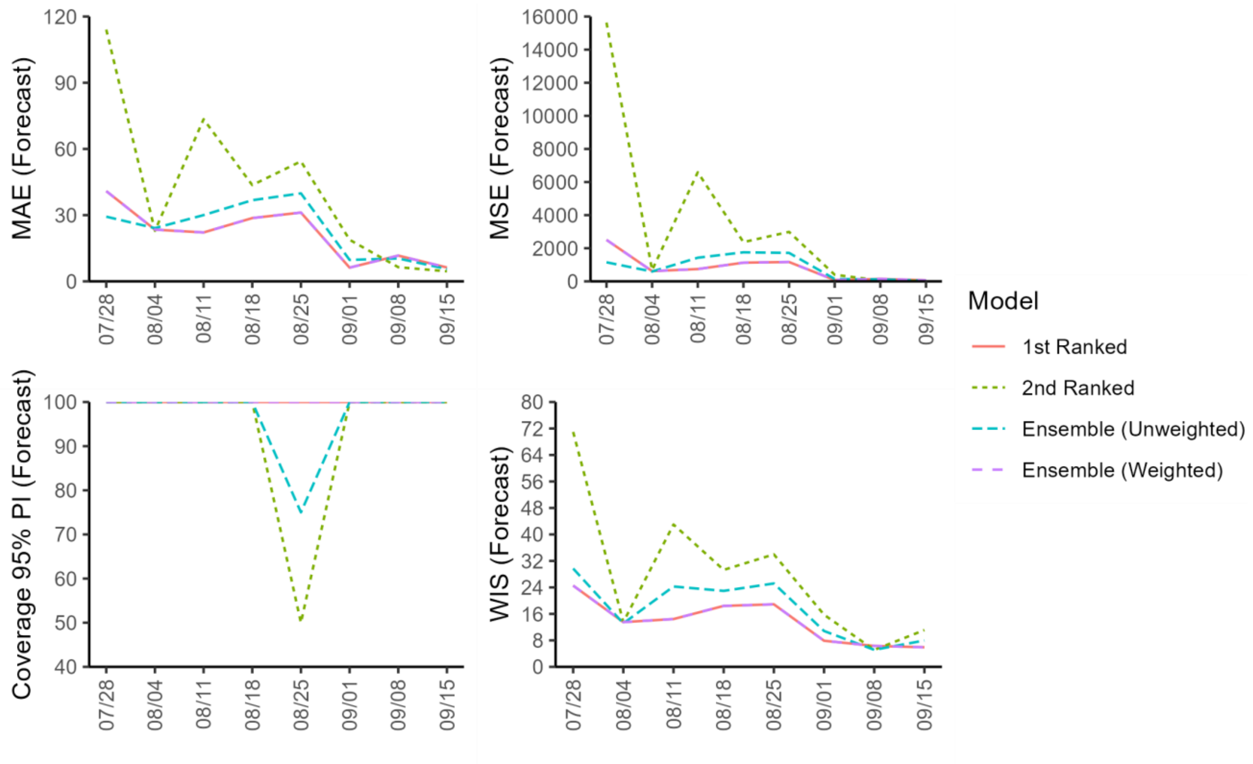


**Figure 6s.** Performance metrics of the forecasts generated by the sub-epidemic models across 8 sequential forecasting periods (Week of July 28th through the week of September 15th, 2022) for Germany.

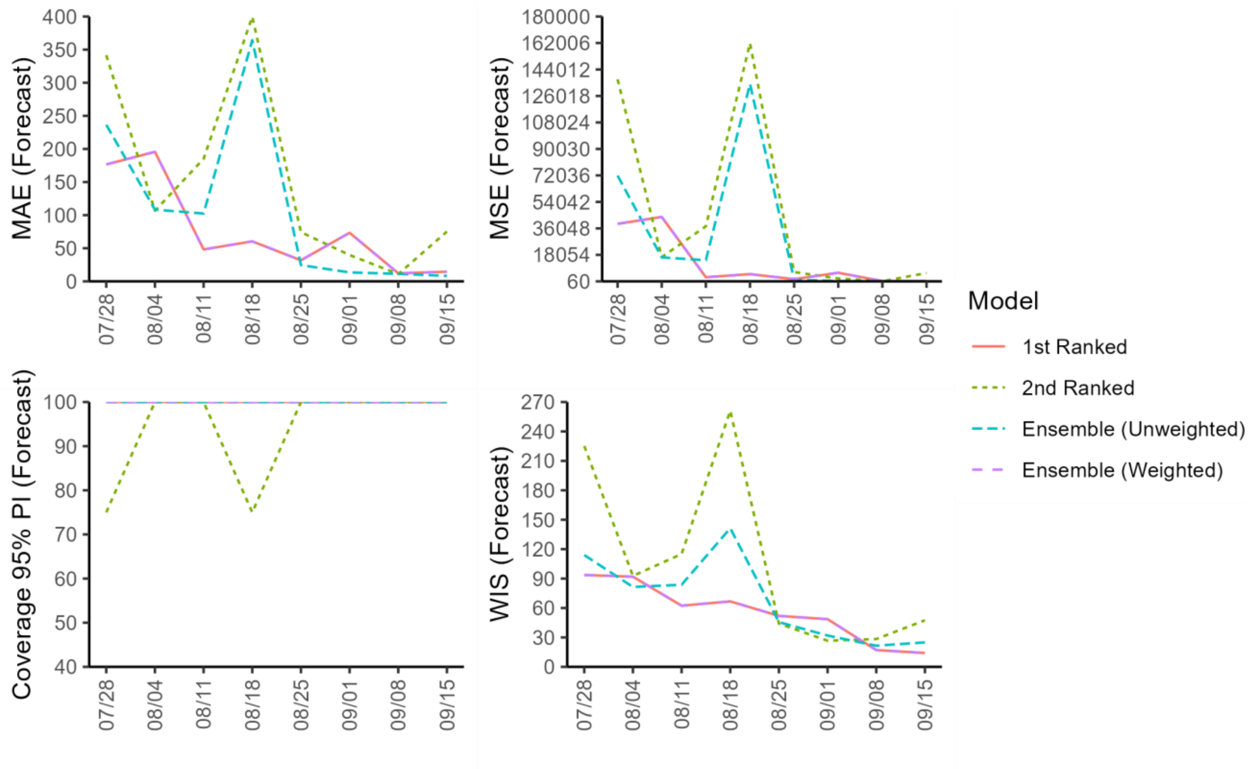


**Figure 7s.** Performance metrics of the forecasts generated by the sub-epidemic models across 8 sequential forecasting periods (Week of July 28th through the week of September 15th, 2022) for Spain.

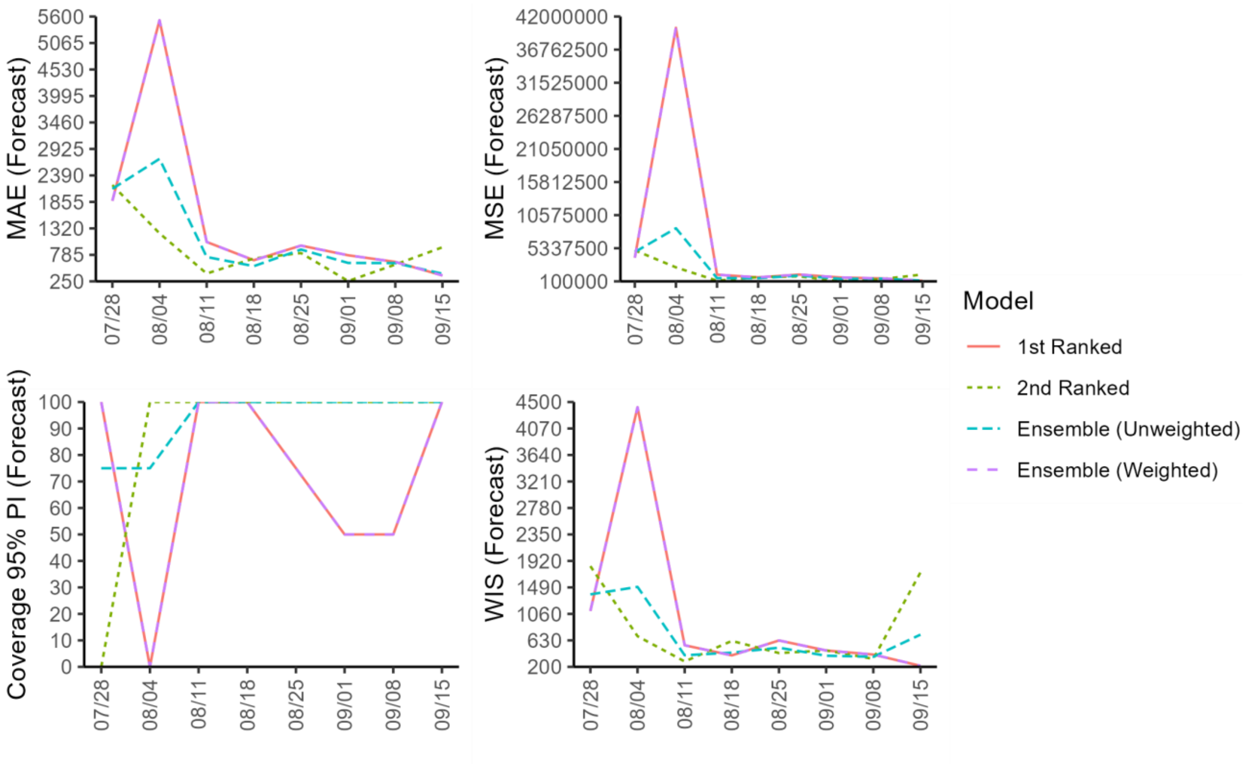


**Figure 8s.** Performance metrics of the forecasts generated by the sub-epidemic models across 8 sequential forecasting periods (Week of July 28th through the week of September 15th, 2022) for the United States (OWID).

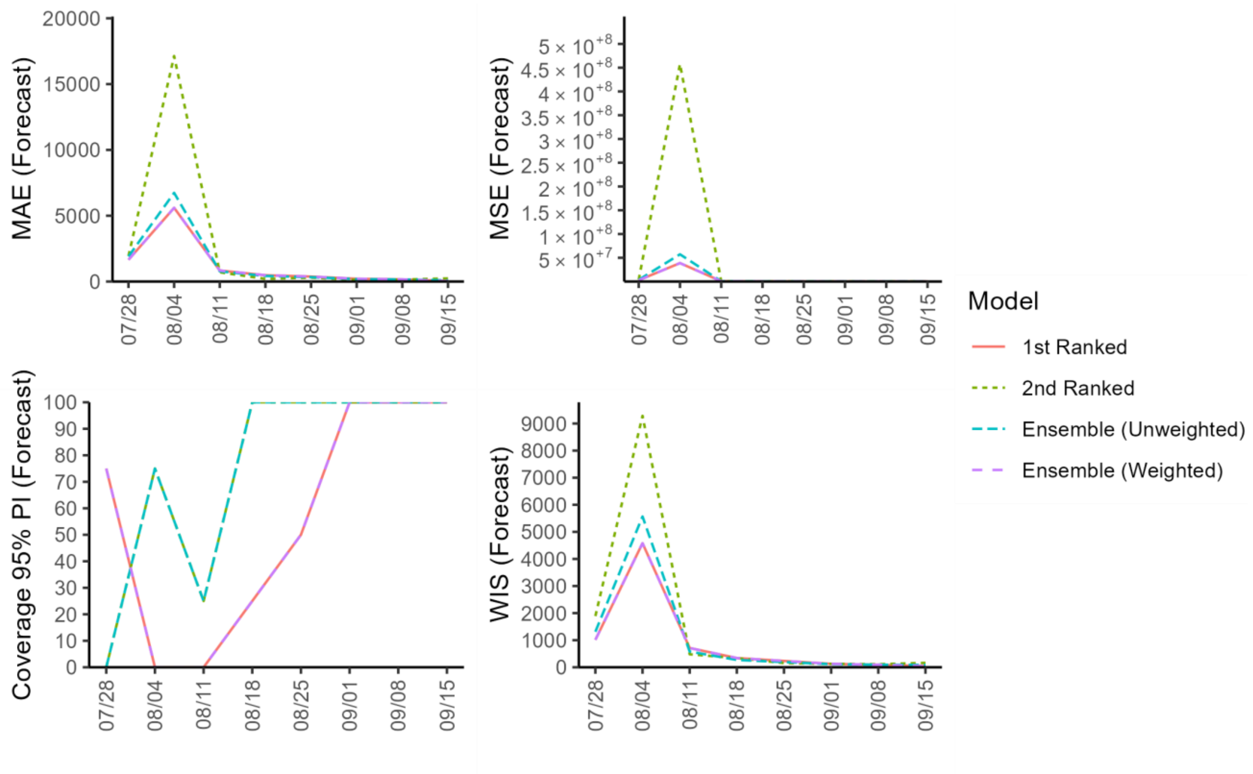


**Figure 9s.** Performance metrics of the forecasts generated by the sub-epidemic models across 8 sequential forecasting periods (Week of July 28th through the week of September 15th, 2022) for the United States (CDC).

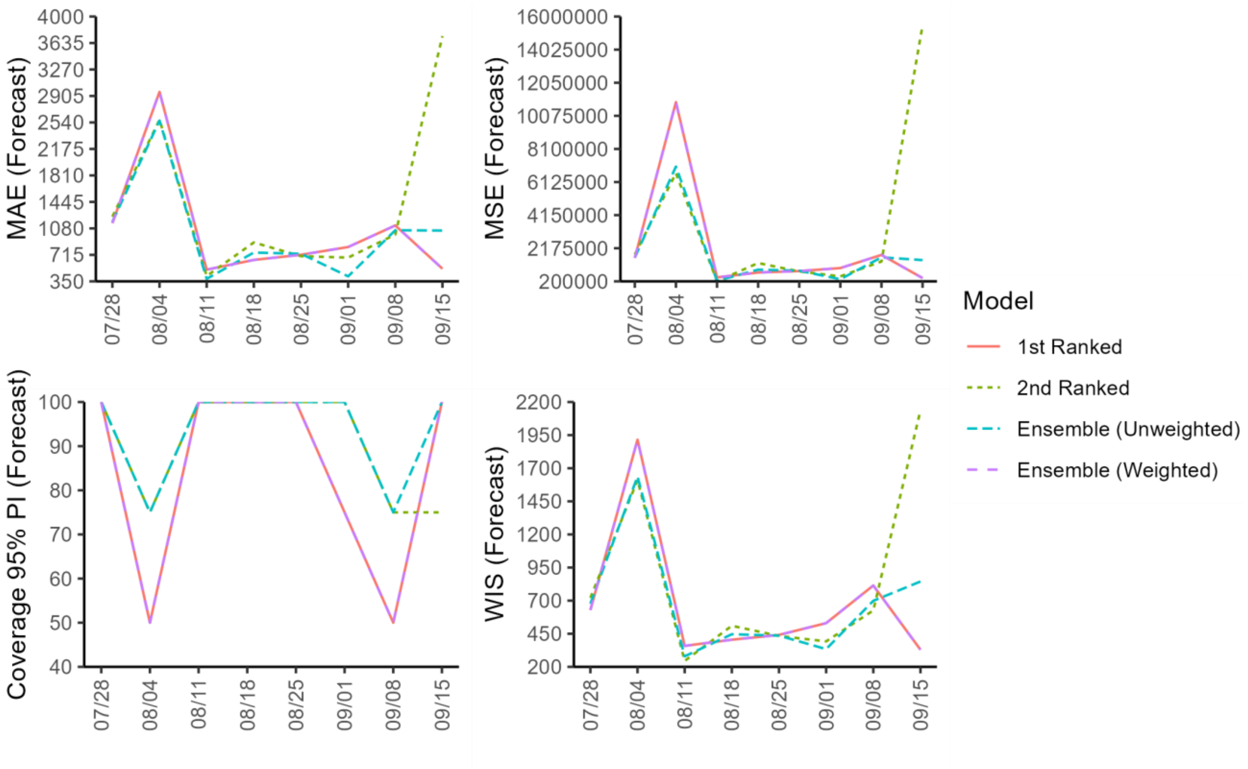


**Figure 10s.** Performance metrics of the forecasts generated by the sub-epidemic models across 8 sequential forecasting periods (Week of July 28th through the week of September 15th, 2022) for the World.

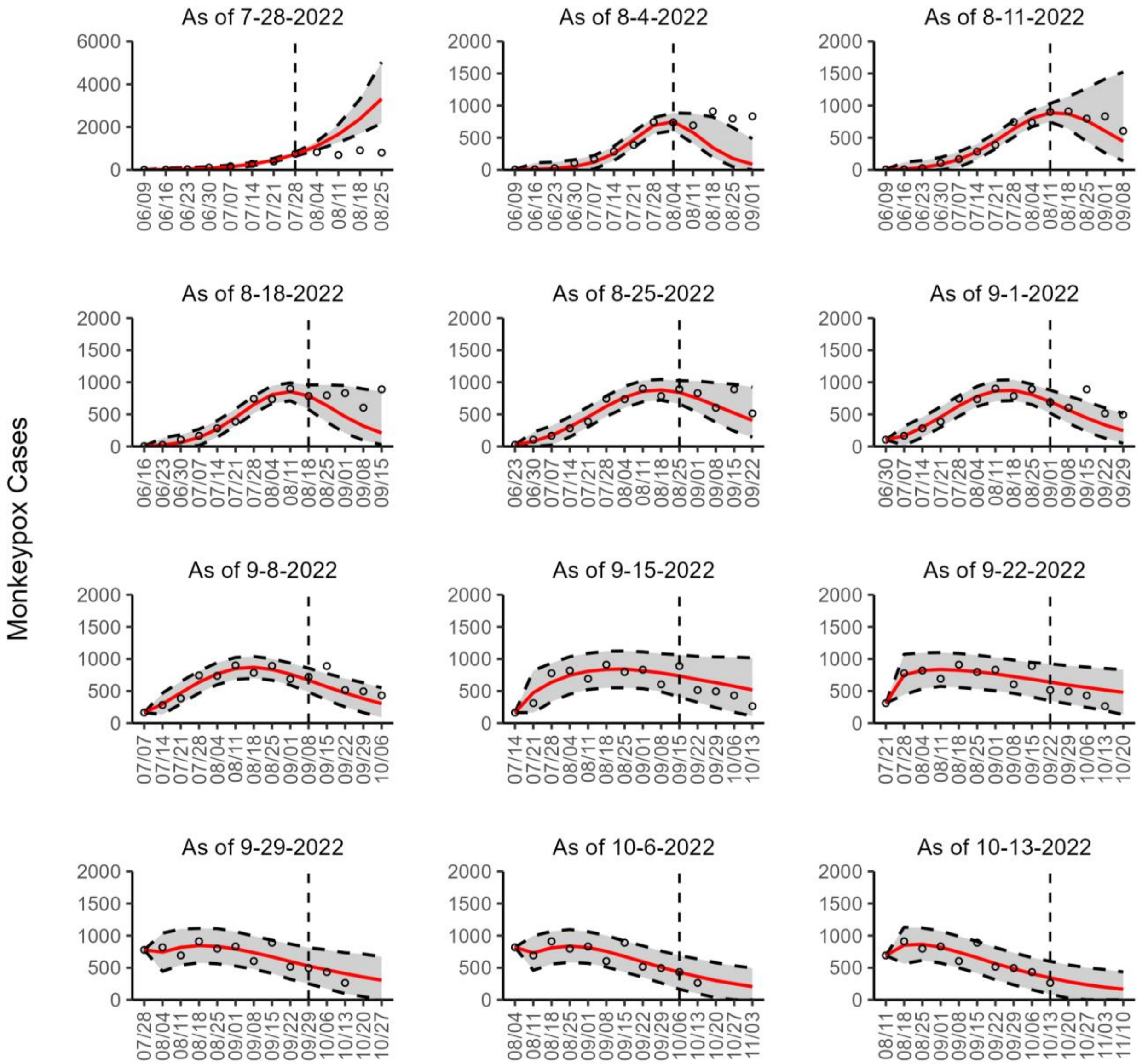


**Figure 11s.** The overlayed forecasted and reported monkeypox cases for the weeks of 7/28/2022 through the week of 10/13/2022 for Brazil. The forecasts are derived from the top-ranked sub-epidemic model using 10-week calibration data, and the reported cases are obtained from the OWID GitHub [25]. The black circles to the left of the vertical line represent the reported cases as of the Friday of the forecast period; the solid red line corresponds to the best fit model; the dashed black lines correspond to the 95% prediction intervals. The black circles to the right of the vertical line represent the reported case counts (as of 10/21/2022) for the corresponding date. The vertical dashed black line indicates the start of the forecast period. For the week of 7/28/2022, data posted by the OWID team on 8/9/2022 was used to produce the forecast as it was the earliest version of data available.

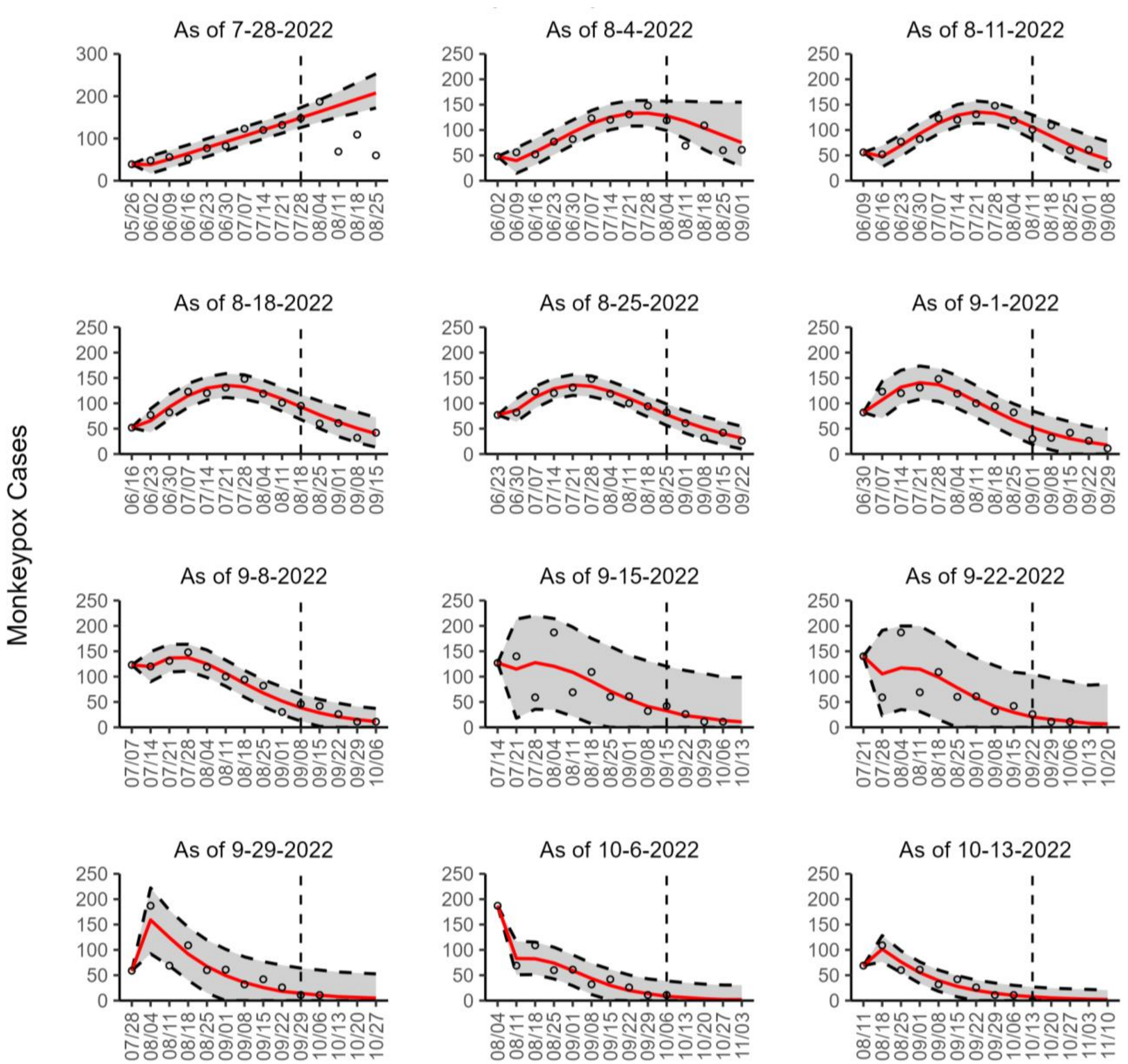


**Figure 12s.** The overlayed forecasted and reported monkeypox cases for the weeks of 7/28/2022 through the week of 10/13/2022 for Canada. The forecasts are derived from the top-ranked sub-epidemic model using 10-week calibration data, and the reported cases are obtained from the OWID GitHub [25]. The black circles to the left of the vertical line represent the reported cases as of the Friday of the forecast period; the solid red line corresponds to the best fit model; the dashed black lines correspond to the 95% prediction intervals. The black circles to the right of the vertical line represent the reported case counts (as of 10/21/2022) for the corresponding date. The vertical dashed black line indicates the start of the forecast period. For the week of 7/28/2022, data posted by the OWID team on 8/9/2022 was used to produce the forecast as it was the earliest version of data available.

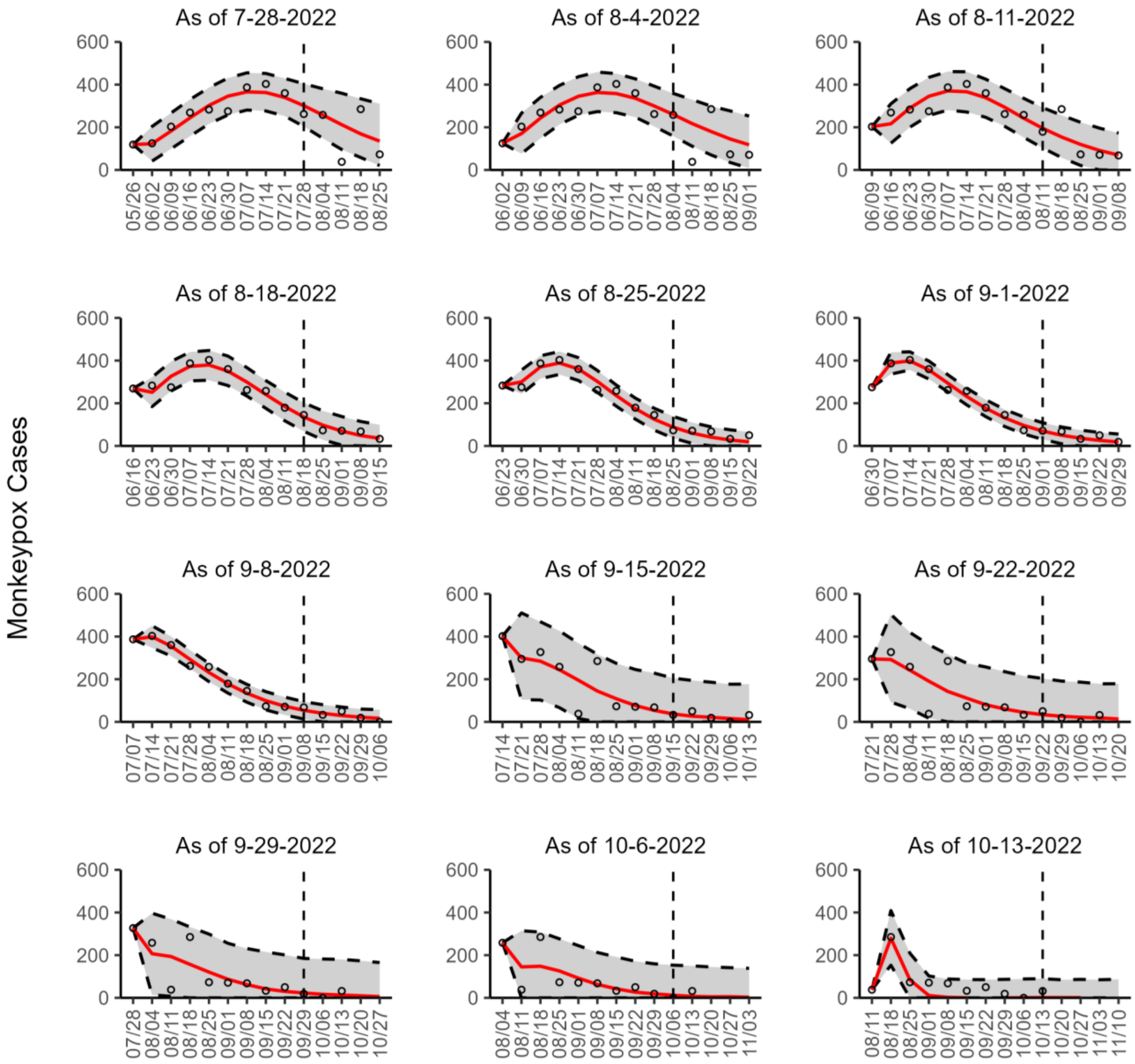


**Figure 13s.** The overlayed forecasted and reported monkeypox cases for the weeks of 7/28/2022 through the week of 10/13/2022 for England. The forecasts are derived from the top-ranked sub-epidemic model using 10-week calibration data, and the reported cases are obtained from the OWID GitHub [25]. The black circles to the left of the vertical line represent the reported cases as of the Friday of the forecast period; the solid red line corresponds to the best fit model; the dashed black lines correspond to the 95% prediction intervals. The black circles to the right of the vertical line represent the reported case counts (as of 10/21/2022) for the corresponding date. The vertical dashed black line indicates the start of the forecast period. For the week of 7/28/2022, data posted by the OWID team on 8/9/2022 was used to produce the forecast as it was the earliest version of data available.

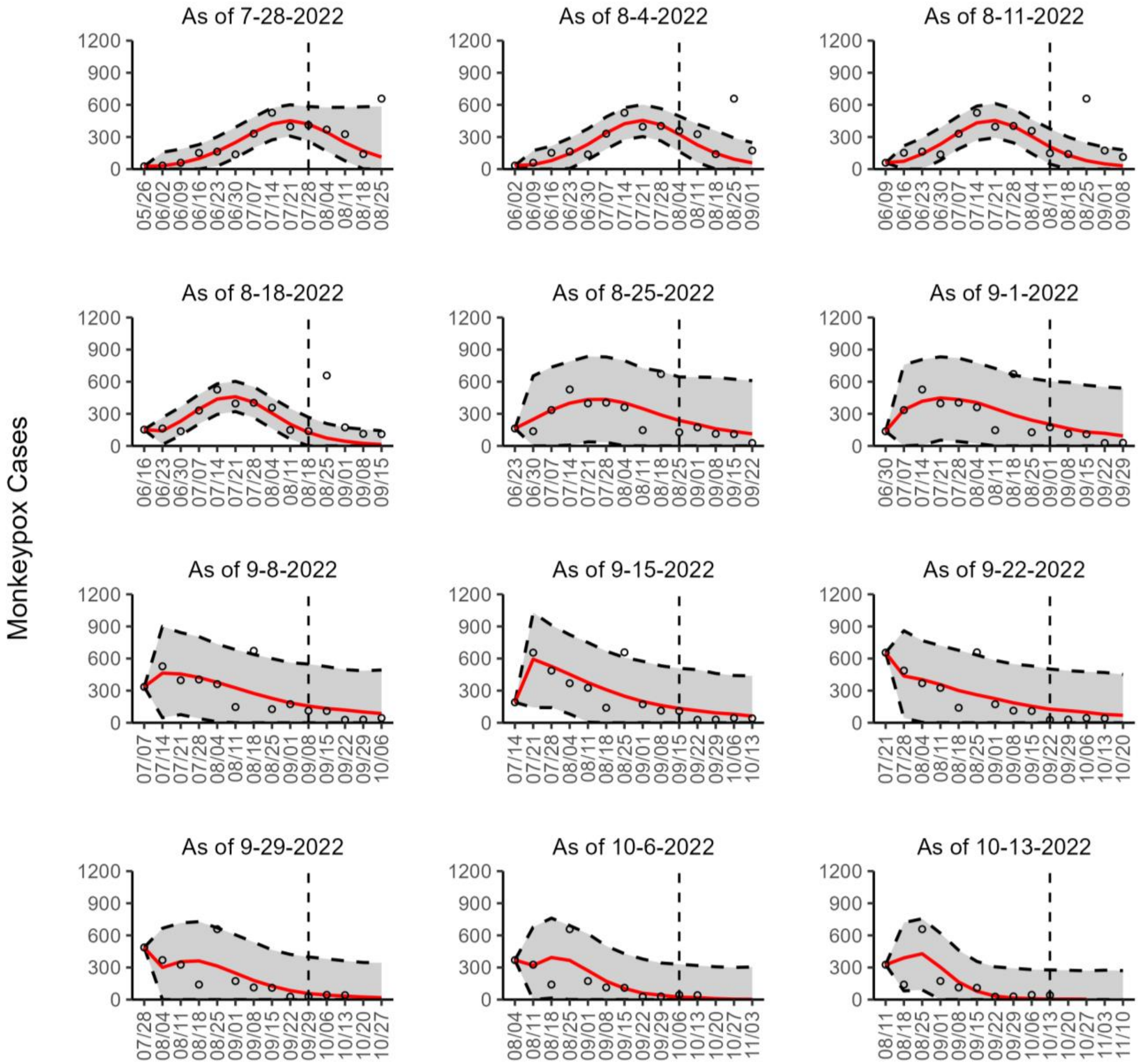


**Figure 14s.** The overlayed forecasted and reported monkeypox cases for the weeks of 7/28/2022 through the week of 10/13/2022 for France. The forecasts are derived from the top-ranked sub-epidemic model using 10-week calibration data, and the reported cases are obtained from the OWID GitHub [25]. The black circles to the left of the vertical line represent the reported cases as of the Friday of the forecast period; the solid red line corresponds to the best fit model; the dashed black lines correspond to the 95% prediction intervals. The black circles to the right of the vertical line represent the reported case counts (as of 10/21/2022) for the corresponding date. The vertical dashed black line indicates the start of the forecast period. For the week of 7/28/2022, data posted by the OWID team on 8/9/2022 was used to produce the forecast as it was the earliest version of data available.

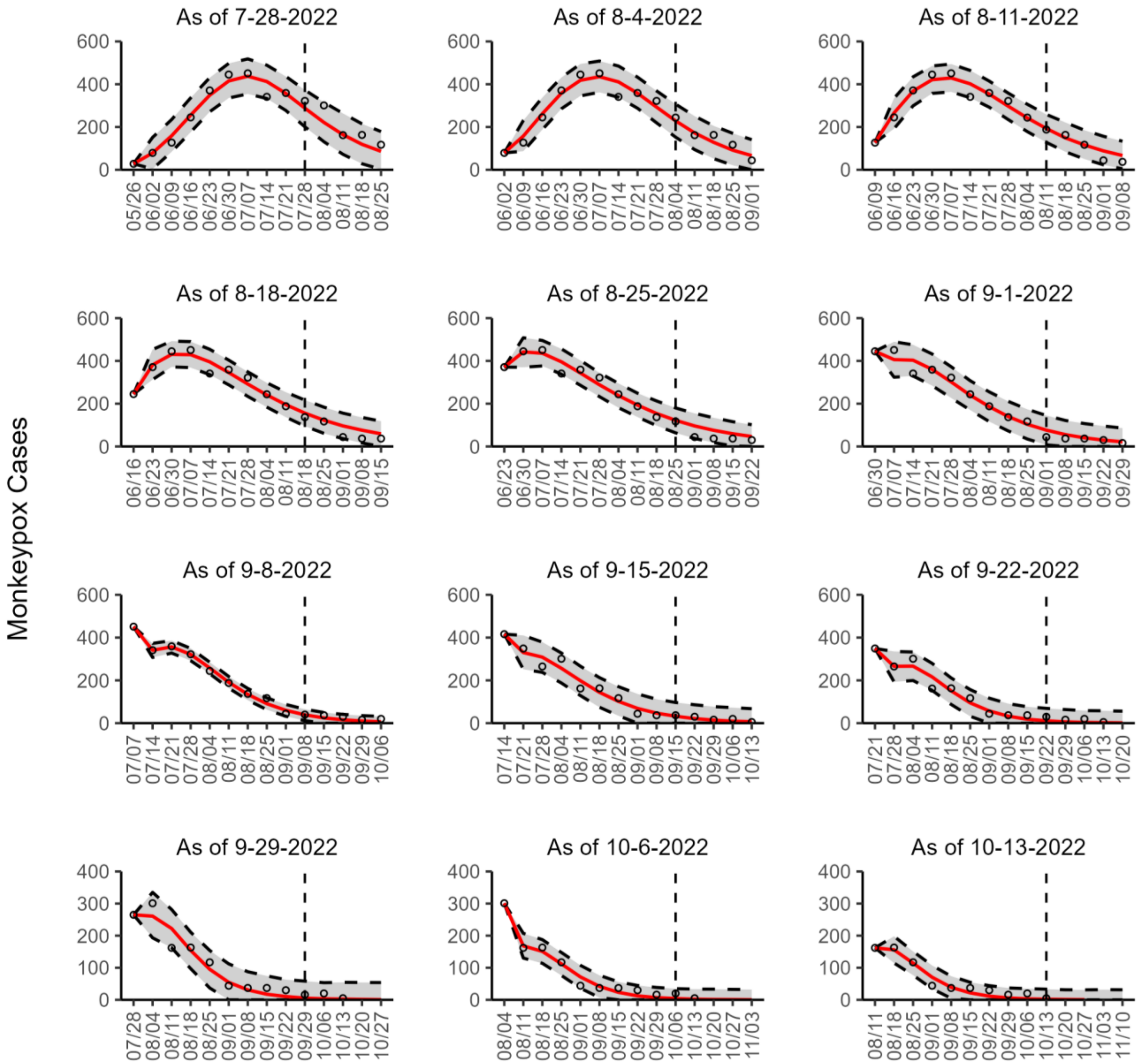


**Figure 15s.** The overlayed forecasted and reported monkeypox cases for the weeks of 7/28/2022 through the week of 10/13/2022 for Germany. The forecasts are derived from the top-ranked sub-epidemic model using 10-week calibration data, and the reported cases are obtained from the OWID GitHub [25]. The black circles to the left of the vertical line represent the reported cases as of the Friday of the forecast period; the solid red line corresponds to the best fit model; the dashed black lines correspond to the 95% prediction intervals. The black circles to the right of the vertical line represent the reported case counts (as of 10/21/2022) for the corresponding date. The vertical dashed black line indicates the start of the forecast period. For the week of 7/28/2022, data posted by the OWID team on 8/9/2022 was used to produce the forecast as it was the earliest version of data available.

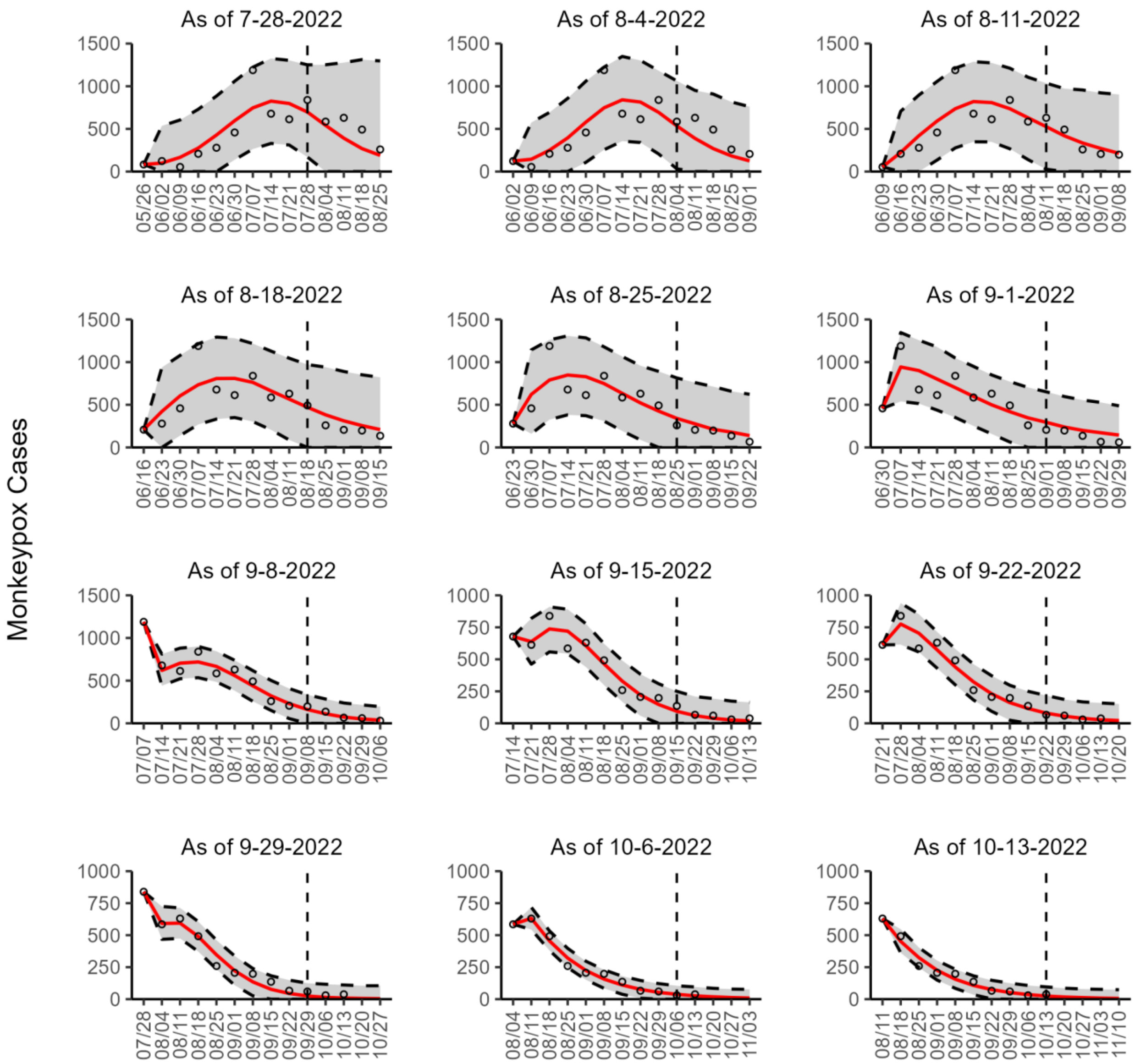


**Figure 16s.** The overlayed forecasted and reported monkeypox cases for the weeks of 7/28/2022 through the week of 10/13/2022 for Spain. The forecasts are derived from the top-ranked sub-epidemic model using 10-week calibration data, and the reported cases are obtained from the OWID GitHub [25]. The black circles to the left of the vertical line represent the reported cases as of the Friday of the forecast period; the solid red line corresponds to the best fit model; the dashed black lines correspond to the 95% prediction intervals. The black circles to the right of the vertical line represent the reported case counts (as of 10/21/2022) for the corresponding date. The vertical dashed black line indicates the start of the forecast period. For the week of 7/28/2022, data posted by the OWID team on 8/9/2022 was used to produce the forecast as it was the earliest version of data available.

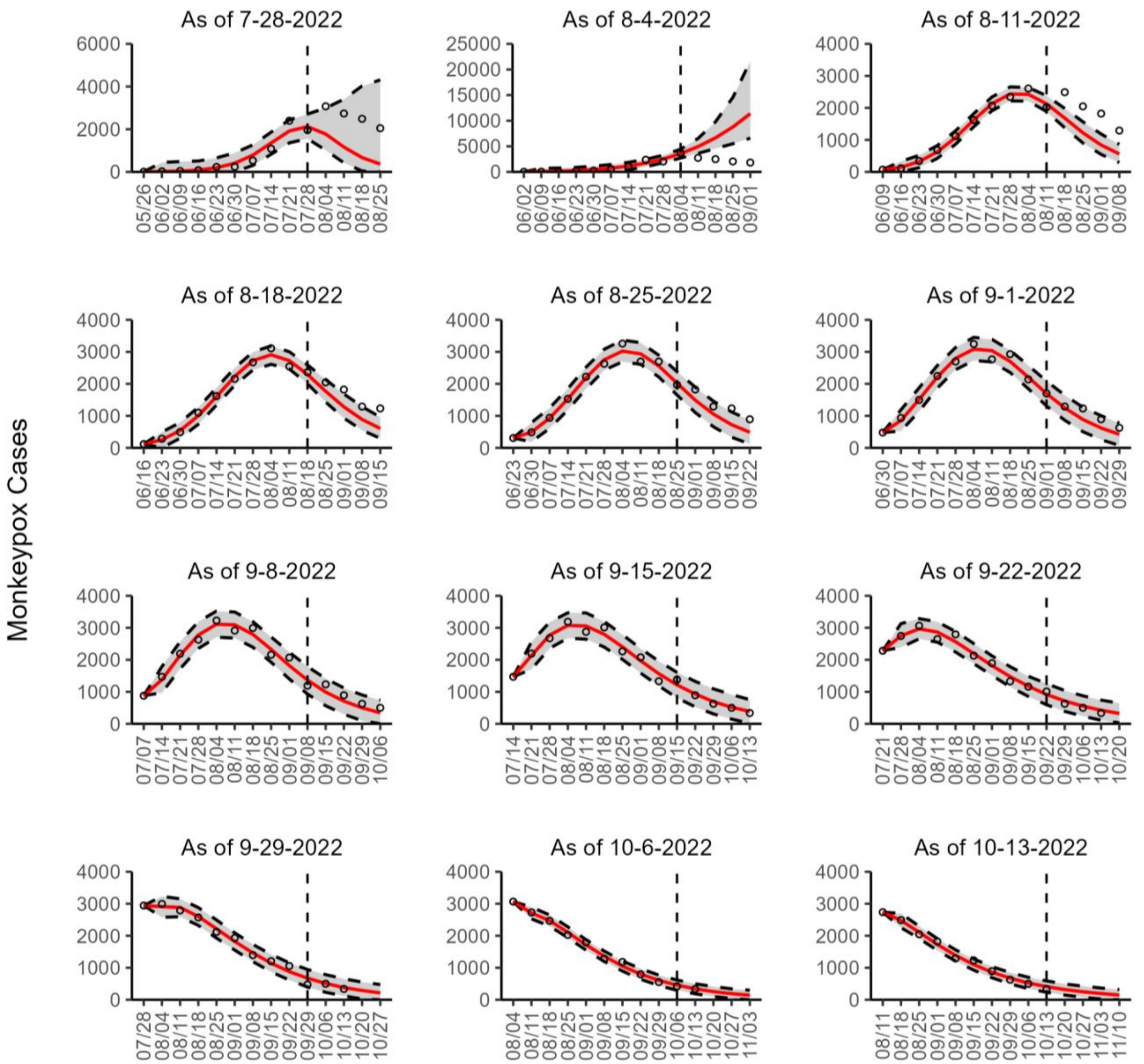


**Figure 17s.** The overlayed forecasted and reported monkeypox cases for the weeks of 7/28/2022 through the week of 10/13/2022 for the United States. The forecasts are derived from the top-ranked sub-epidemic model using 10-week calibration data, and the reported cases are obtained from the CDC [24]. The black circles to the left of the vertical line represent the reported cases as of the Wednesday of the forecast period; the solid red line corresponds to the best fit model; the dashed black lines correspond to the 95% prediction intervals. The black circles to the right of the vertical line represent the reported case counts (as of 10/19/2022) for the corresponding date.

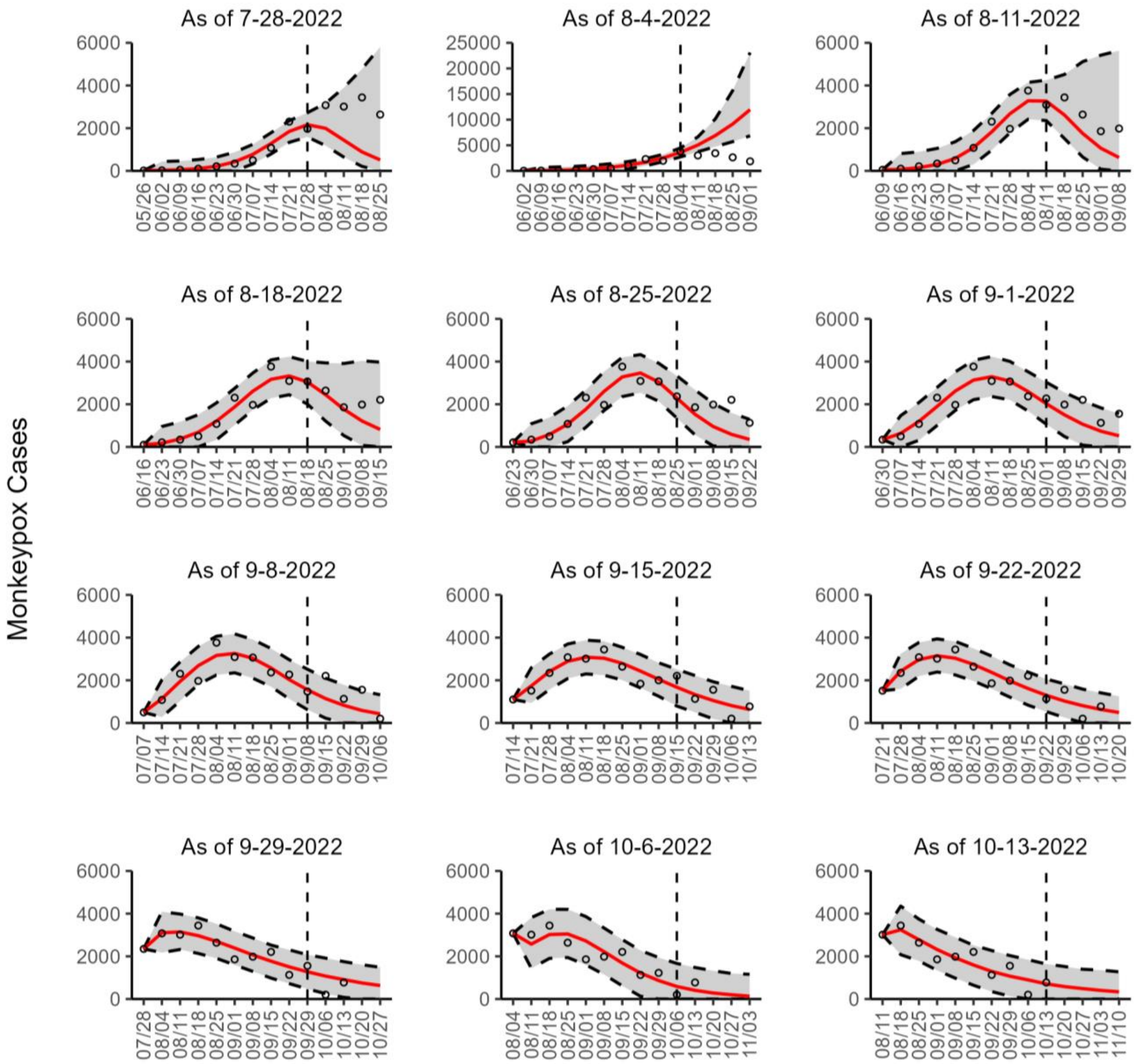


**Figure 18s.** The overlayed forecasted and reported monkeypox cases for the weeks of 7/28/2022 through the week of 10/13/2022 for the United States. The forecasts are derived from the top-ranked sub-epidemic model using 10-week calibration data, and the reported cases are obtained from the OWID GitHub [25]. The black circles to the left of the vertical line represent the reported cases as of the Friday of the forecast period; the solid red line corresponds to the best fit model; the dashed black lines correspond to the 95% prediction intervals. The black circles to the right of the vertical line represent the reported case counts (as of 10/21/2022) for the corresponding date. The vertical dashed black line indicates the start of the forecast period. For the week of 7/28/2022, data posted by the OWID team on 8/9/2022 was used to produce the forecast as it was the earliest version of data available.

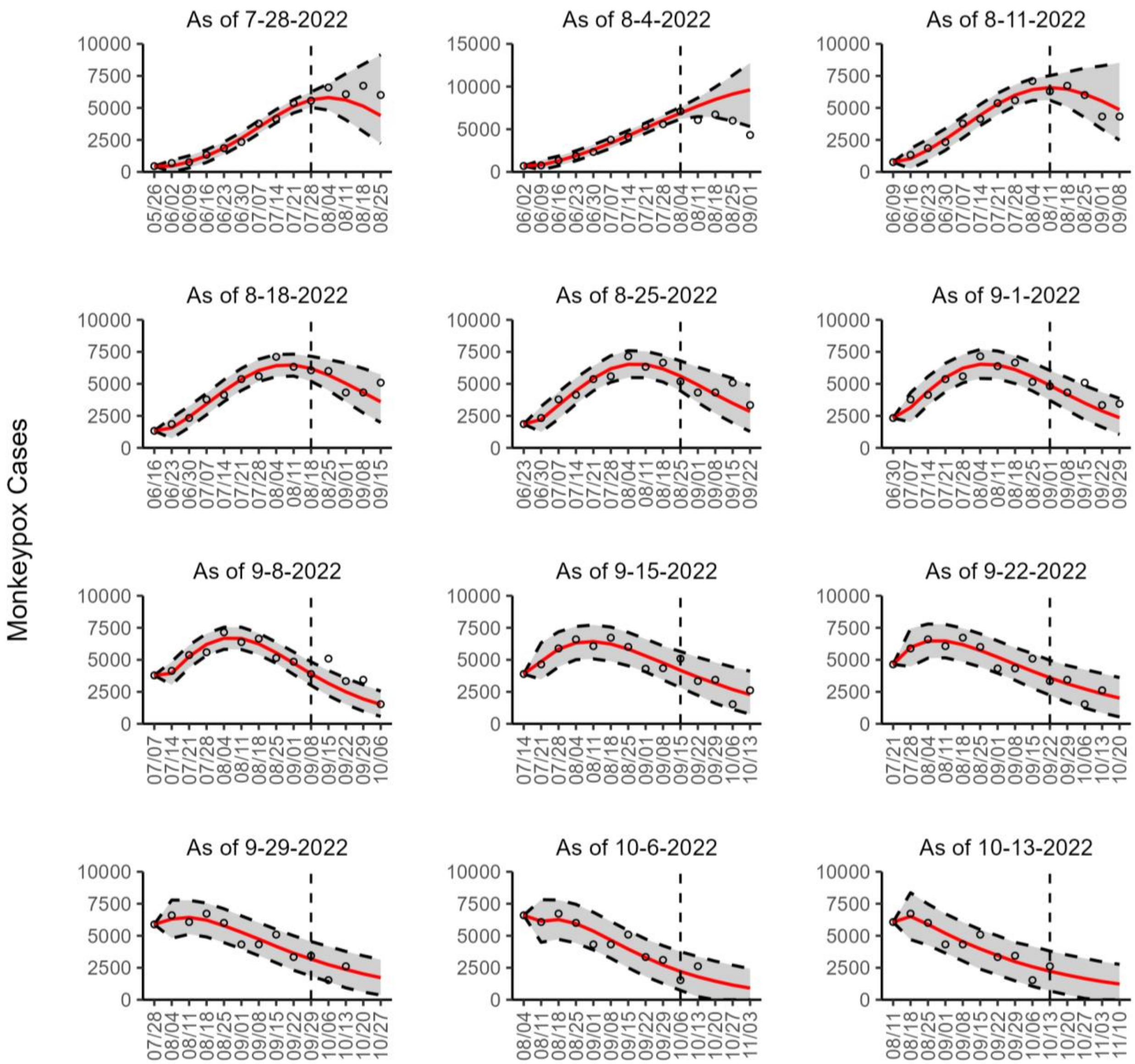


**Figure 19s.** The overlayed forecasted and reported monkeypox cases for the weeks of 7/28/2022 through the week of 10/13/2022 for the World. The forecasts are derived from the top-ranked sub-epidemic model using 10-week calibration data, and the reported cases are obtained from the OWID GitHub [25]. The black circles to the left of the vertical line represent the reported cases as of the Friday of the forecast period; the solid red line corresponds to the best fit model; the dashed black lines correspond to the 95% prediction intervals. The black circles to the right of the vertical line represent the reported case counts (as of 10/21/2022) for the corresponding date. The vertical dashed black line indicates the start of the forecast period. For the week of 7/28/2022, data posted by the OWID team on 8/9/2022 was used to produce the forecast as it was the earliest version of data available.

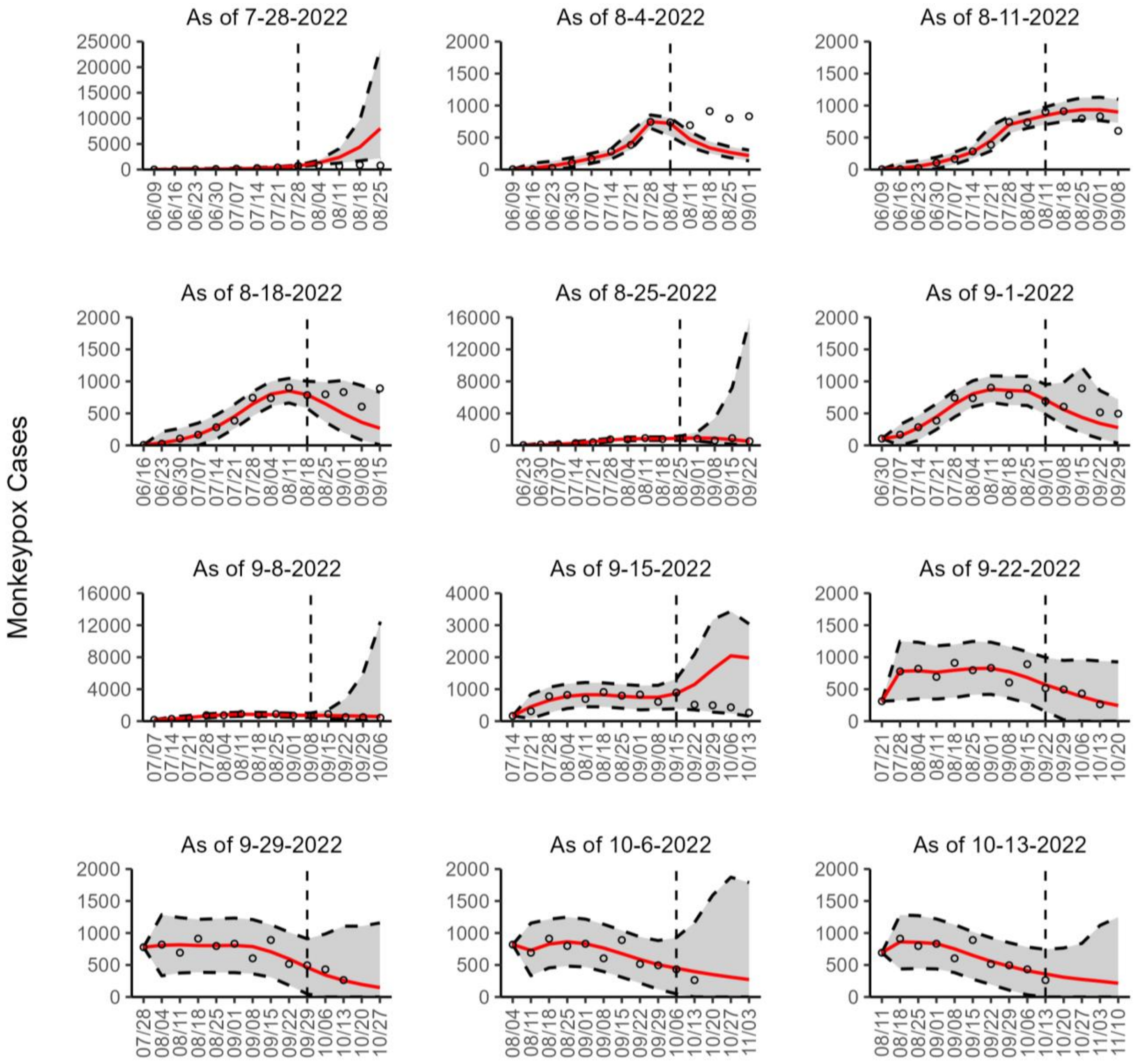


**Figure 20s.** The overlayed forecasted and reported monkeypox cases for the weeks of 7/28/2022 through the week of 10/13/2022 for Brazil. The forecasts are derived from the second-ranked sub-epidemic model using 10-week calibration data, and the reported cases are obtained from the OWID GitHub [25]. The black circles to the left of the vertical line represent the reported cases as of the Friday of the forecast period; the solid red line corresponds to the best fit model; the dashed black lines correspond to the 95% prediction intervals. The black circles to the right of the vertical line represent the reported case counts (as of 10/21/2022) for the corresponding date. The vertical dashed black line indicates the start of the forecast period. For the week of 7/28/2022, data posted by the OWID team on 8/9/2022 was used to produce the forecast as it was the earliest version of data available.

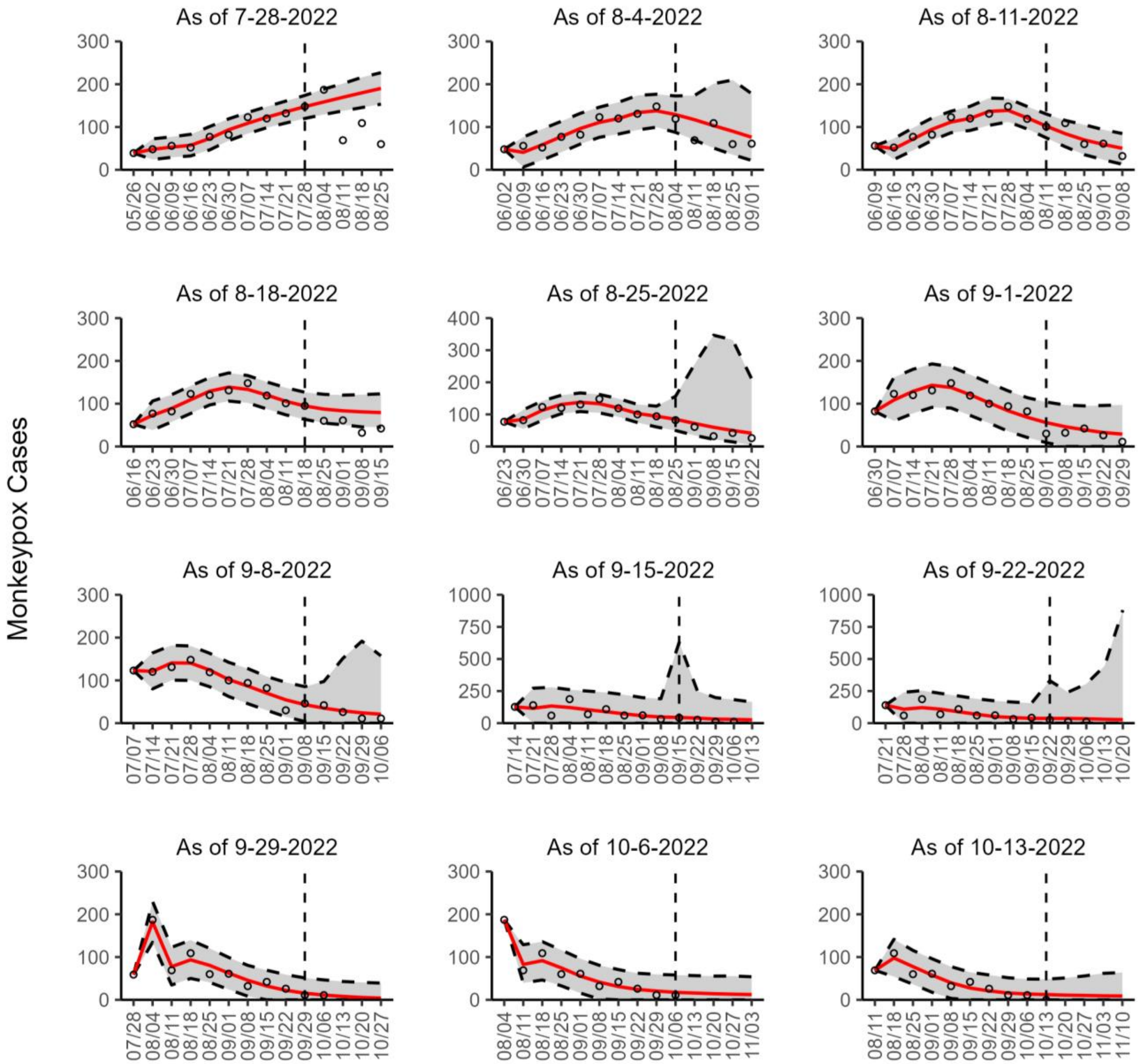


**Figure 21s.** The overlayed forecasted and reported monkeypox cases for the weeks of 7/28/2022 through the week of 10/13/2022 for Canada. The forecasts are derived from the second-ranked sub-epidemic model using 10-week calibration data, and the reported cases are obtained from the OWID GitHub [25]. The black circles to the left of the vertical line represent the reported cases as of the Friday of the forecast period; the solid red line corresponds to the best fit model; the dashed black lines correspond to the 95% prediction intervals. The black circles to the right of the vertical line represent the reported case counts (as of 10/21/2022) for the corresponding date. The vertical dashed black line indicates the start of the forecast period. For the week of 7/28/2022, data posted by the OWID team on 8/9/2022 was used to produce the forecast as it was the earliest version of data available.

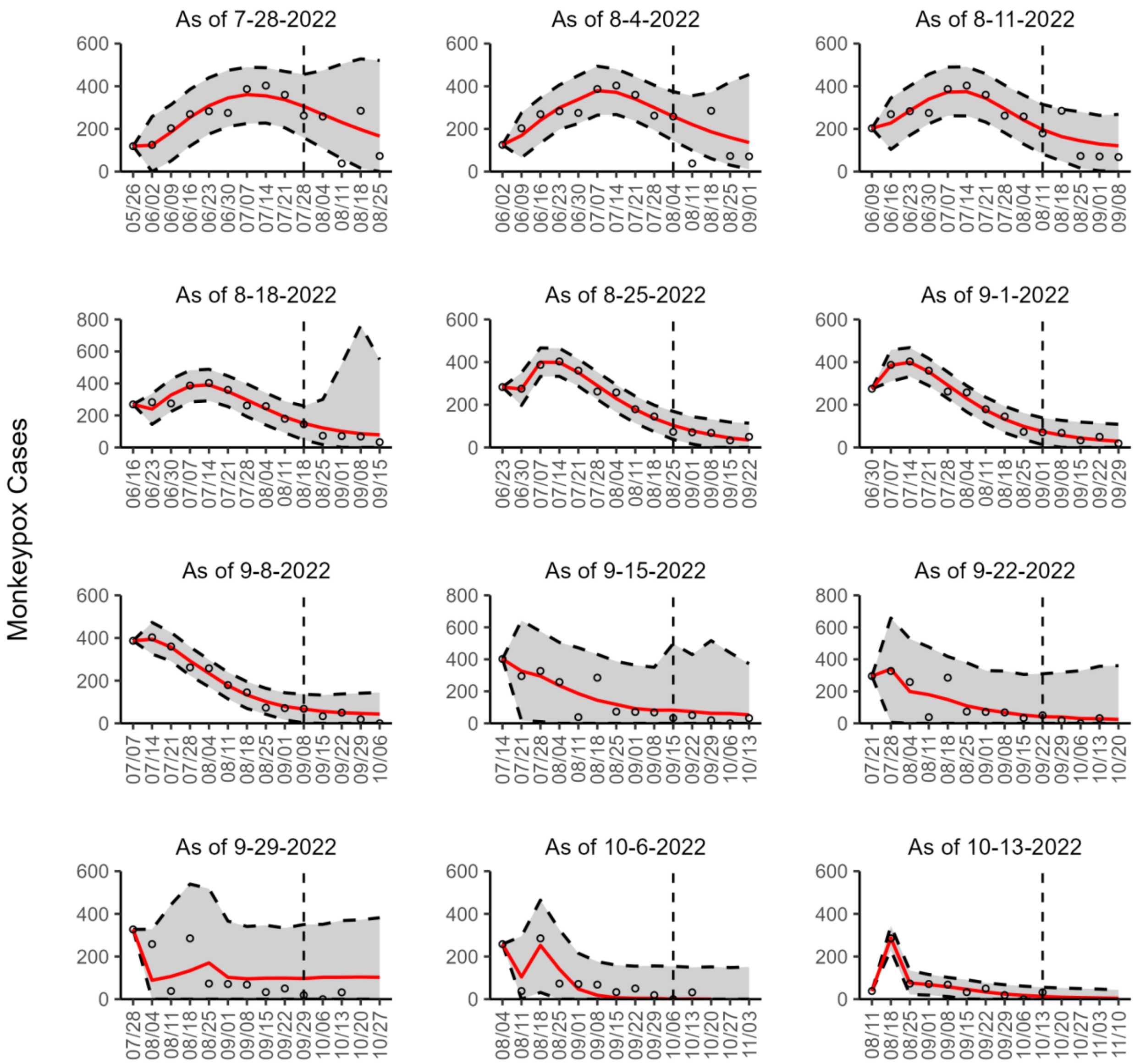


**Figure 22s.** The overlayed forecasted and reported monkeypox cases for the weeks of 7/28/2022 through the week of 10/13/2022 for England. The forecasts are derived from the second-ranked sub-epidemic model using 10-week calibration data, and the reported cases are obtained from the OWID GitHub [25]. The black circles to the left of the vertical line represent the reported cases as of the Friday of the forecast period; the solid red line corresponds to the best fit model; the dashed black lines correspond to the 95% prediction intervals. The black circles to the right of the vertical line represent the reported case counts (as of 10/21/2022) for the corresponding date. The vertical dashed black line indicates the start of the forecast period. For the week of 7/28/2022, data posted by the OWID team on 8/9/2022 was used to produce the forecast as it was the earliest version of data available.

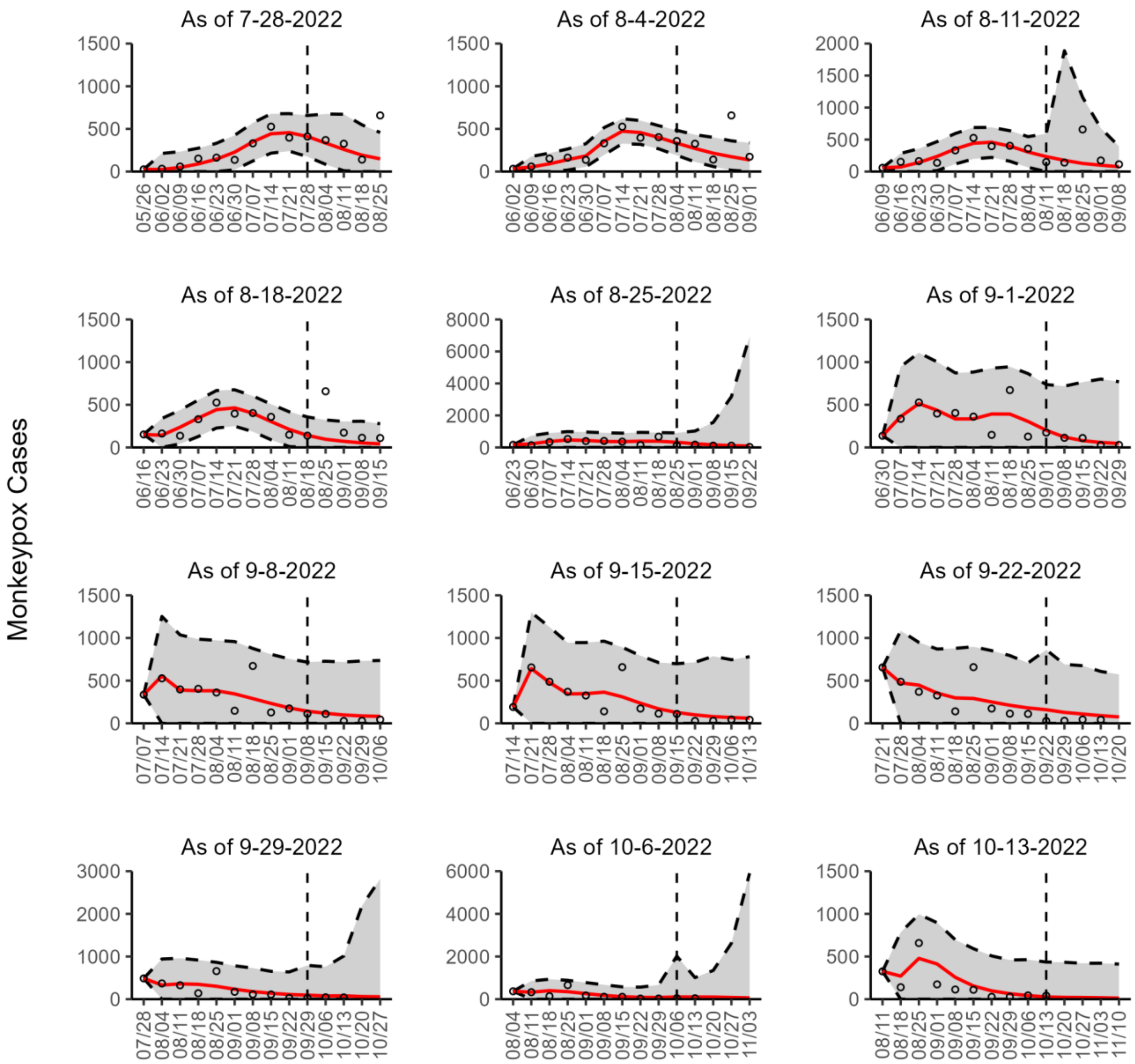


**Figure 23s.** The overlayed forecasted and reported monkeypox cases for the weeks of 7/28/2022 through the week of 10/13/2022 for France. The forecasts are derived from the second-ranked sub-epidemic model using 10-week calibration data, and the reported cases are obtained from the OWID GitHub [25]. The black circles to the left of the vertical line represent the reported cases as of the Friday of the forecast period; the solid red line corresponds to the best fit model; the dashed black lines correspond to the 95% prediction intervals. The black circles to the right of the vertical line represent the reported case counts (as of 10/21/2022) for the corresponding date. The vertical dashed black line indicates the start of the forecast period. For the week of 7/28/2022, data posted by the OWID team on 8/9/2022 was used to produce the forecast as it was the earliest version of data available.

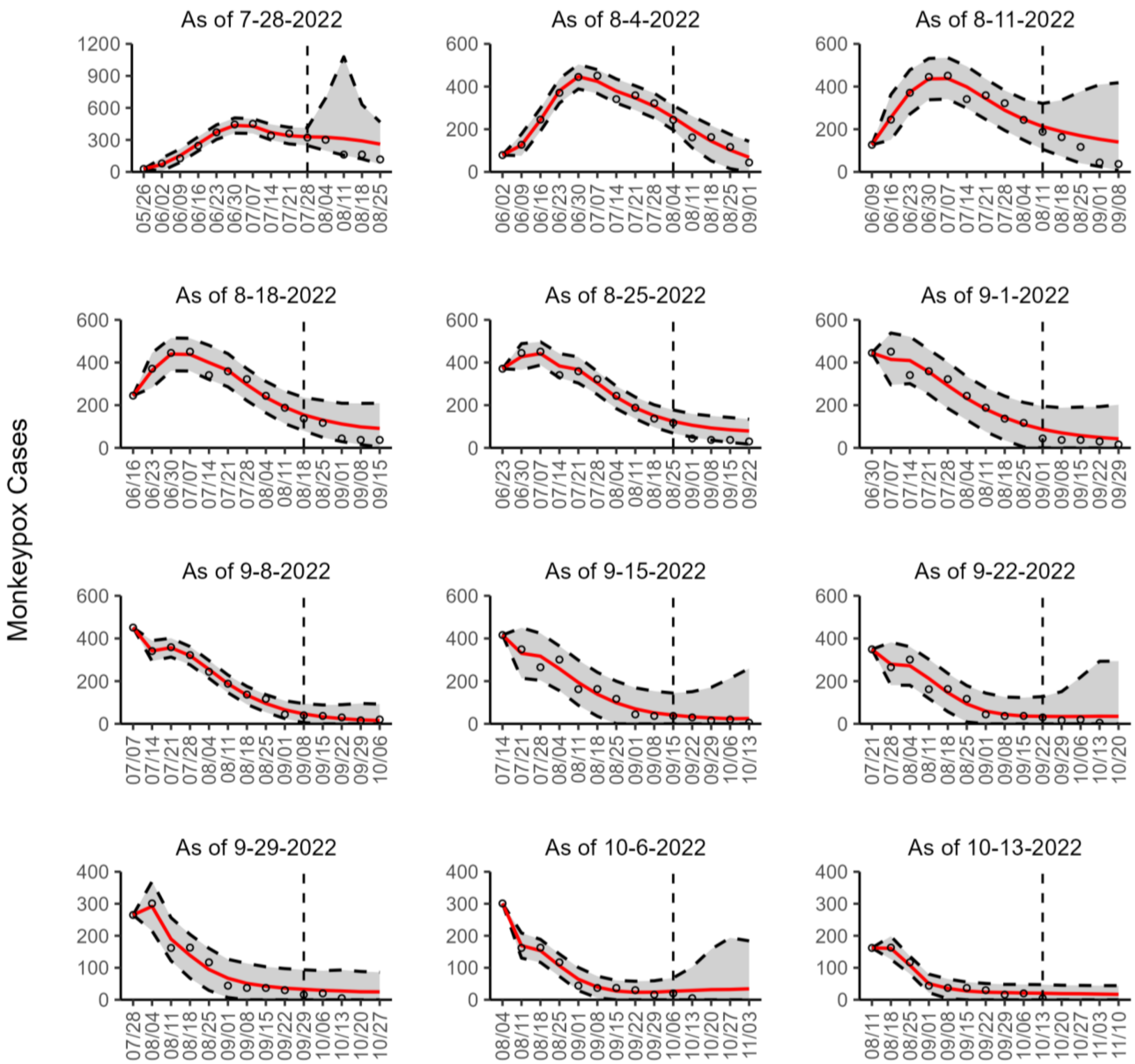


**Figure 24s.** The overlayed forecasted and reported monkeypox cases for the weeks of 7/28/2022 through the week of 10/13/2022 for Germany. The forecasts are derived from the second-ranked sub-epidemic model using 10-week calibration data, and the reported cases are obtained from the OWID GitHub [25]. The black circles to the left of the vertical line represent the reported cases as of the Friday of the forecast period; the solid red line corresponds to the best fit model; the dashed black lines correspond to the 95% prediction intervals. The black circles to the right of the vertical line represent the reported case counts (as of 10/21/2022) for the corresponding date. The vertical dashed black line indicates the start of the forecast period. For the week of 7/28/2022, data posted by the OWID team on 8/9/2022 was used to produce the forecast as it was the earliest version of data available.

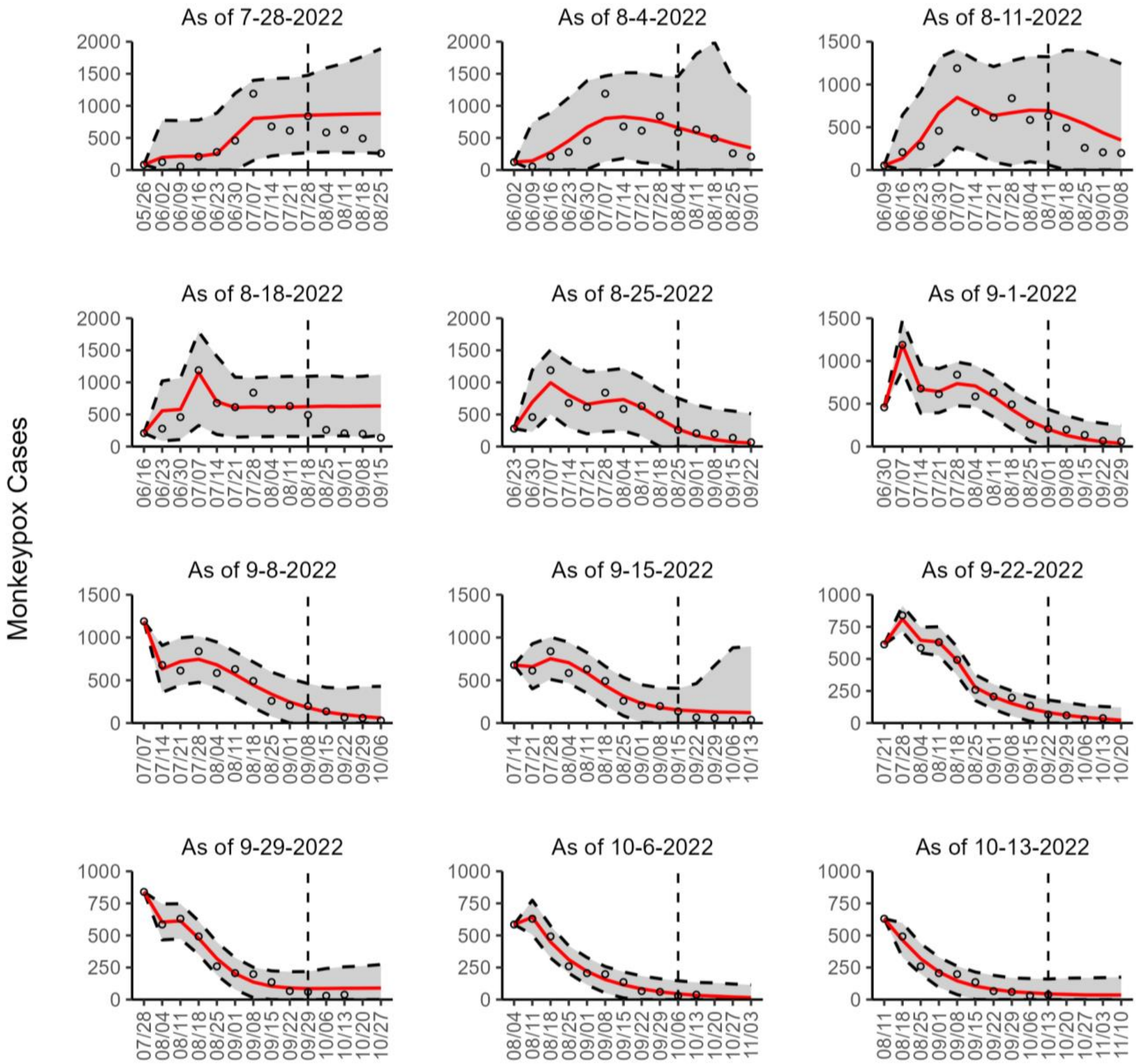


**Figure 25s.** The overlayed forecasted and reported monkeypox cases for the weeks of 7/28/2022 through the week of 10/13/2022 for Spain. The forecasts are derived from the second-ranked sub-epidemic model using 10-week calibration data, and the reported cases are obtained from the OWID GitHub [25]. The black circles to the left of the vertical line represent the reported cases as of the Friday of the forecast period; the solid red line corresponds to the best fit model; the dashed black lines correspond to the 95% prediction intervals. The black circles to the right of the vertical line represent the reported case counts (as of 10/21/2022) for the corresponding date. The vertical dashed black line indicates the start of the forecast period. For the week of 7/28/2022, data posted by the OWID team on 8/9/2022 was used to produce the forecast as it was the earliest version of data available.

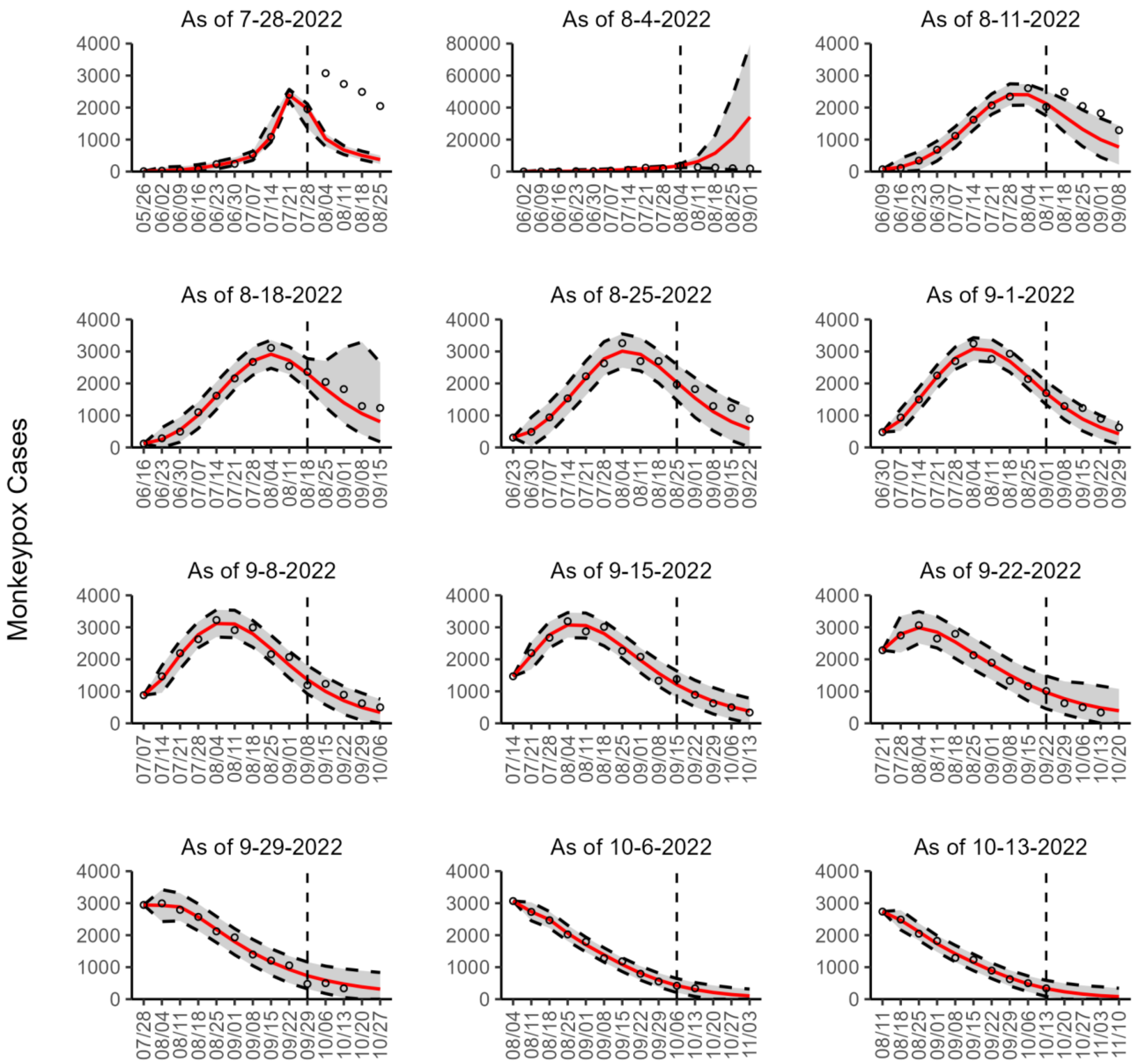


**Figure 26s.** The overlayed forecasted and reported monkeypox cases for the weeks of 7/28/2022 through the week of 10/13/2022 for the United States. The forecasts are derived from the second-ranked sub-epidemic model using 10-week calibration data, and the reported cases are obtained from the CDC [24]. The black circles to the left of the vertical line represent the reported cases as of the Wednesday of the forecast period; the solid red line corresponds to the best fit model; the dashed black lines correspond to the 95% prediction intervals. The black circles to the right of the vertical line represent the reported case counts (as of 10/19/2022) for the corresponding date.

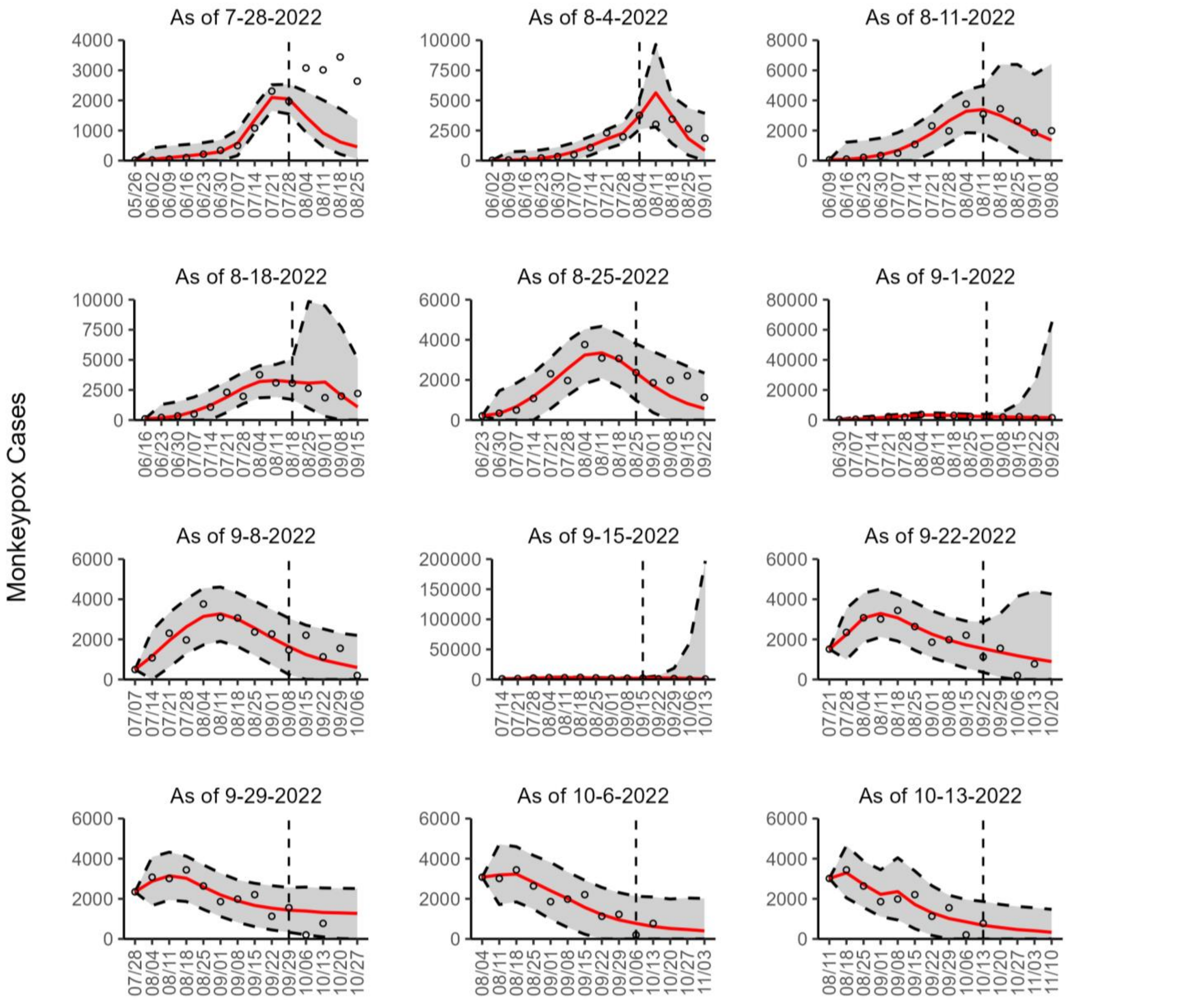


**Figure 27s.** The overlayed forecasted and reported monkeypox cases for the weeks of 7/28/2022 through the week of 10/13/2022 for the United States. The forecasts are derived from the second-ranked sub-epidemic model using 10-week calibration data, and the reported cases are obtained from the OWID GitHub [25]. The black circles to the left of the vertical line represent the reported cases as of the Friday of the forecast period; the solid red line corresponds to the best fit model; the dashed black lines correspond to the 95% prediction intervals. The black circles to the right of the vertical line represent the reported case counts (as of 10/21/2022) for the corresponding date. The vertical dashed black line indicates the start of the forecast period. For the week of 7/28/2022, data posted by the OWID team on 8/9/2022 was used to produce the forecast as it was the earliest version of data available.

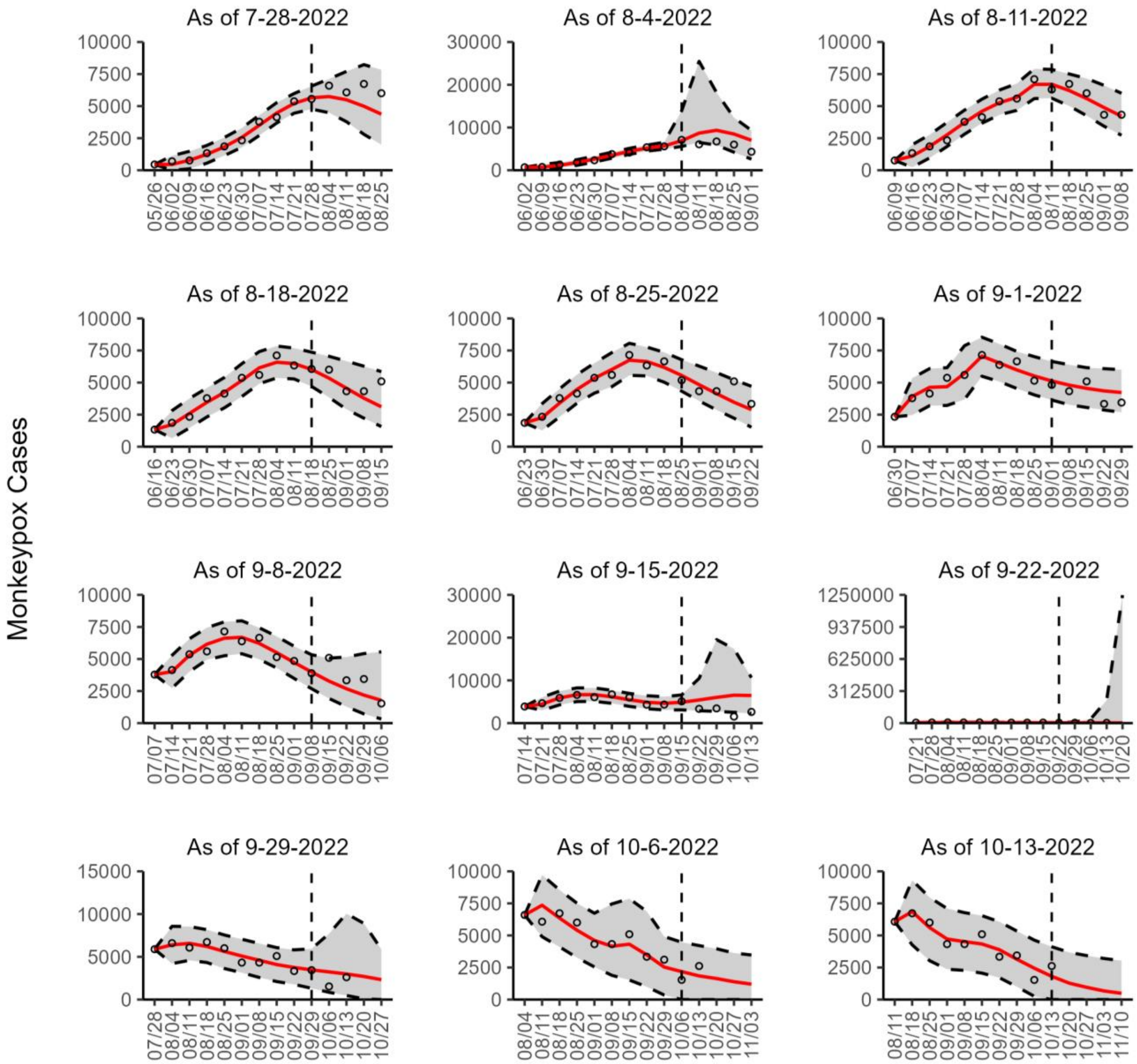


**Figure 28s.** The overlayed forecasted and reported monkeypox cases for the weeks of 7/28/2022 through the week of 10/13/2022 for the World. The forecasts are derived from the second-ranked sub-epidemic model using 10-week calibration data, and the reported cases are obtained from the OWID GitHub [25]. The black circles to the left of the vertical line represent the reported cases as of the Friday of the forecast period; the solid red line corresponds to the best fit model; the dashed black lines correspond to the 95% prediction intervals. The black circles to the right of the vertical line represent the reported case counts (as of 10/21/2022) for the corresponding date. The vertical dashed black line indicates the start of the forecast period. For the week of 7/28/2022, data posted by the OWID team on 8/9/2022 was used to produce the forecast as it was the earliest version of data available.

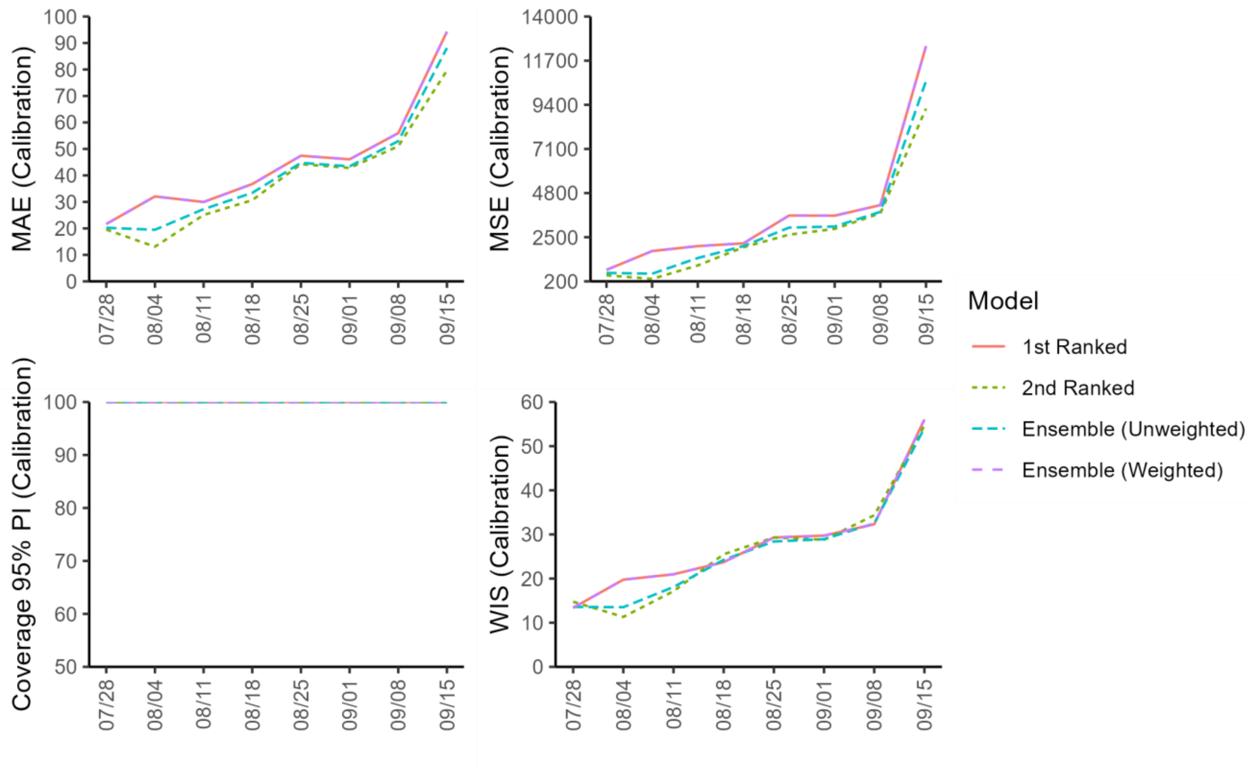


**Figure 29s.** Mean performance metrics for Brazil quantifying model fit across 20 sequential 10-week calibration periods (Week of May 26th – October 13th, 2022) using weekly timeseries data from the OWID team over each 8 forecasting periods**.** Only forecast periods in which observed data was available are included.

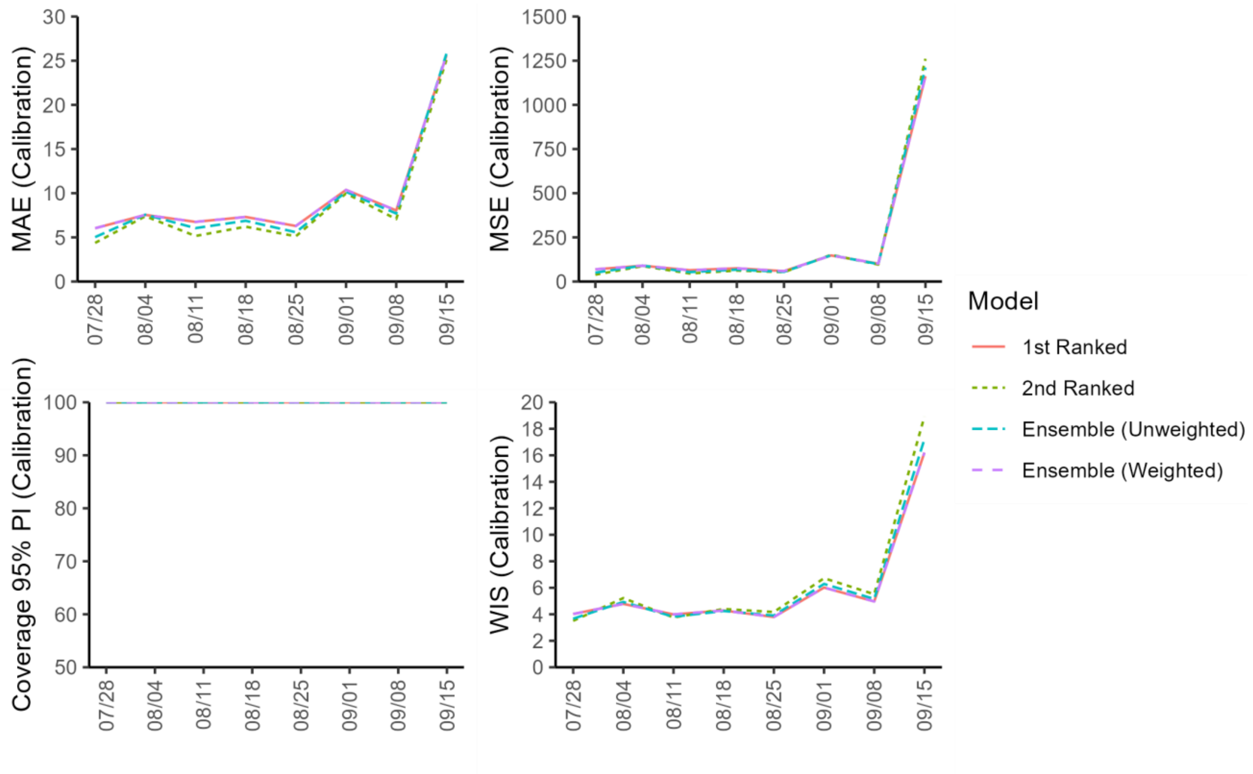


**Figure 30s.** Mean performance metrics for Canada quantifying model fit across 20 sequential 10-week calibration periods (Week of May 26th – October 13th, 2022) using weekly timeseries data from the OWID team over each 8 forecasting periods**.** Only forecast periods in which observed data was available are included.

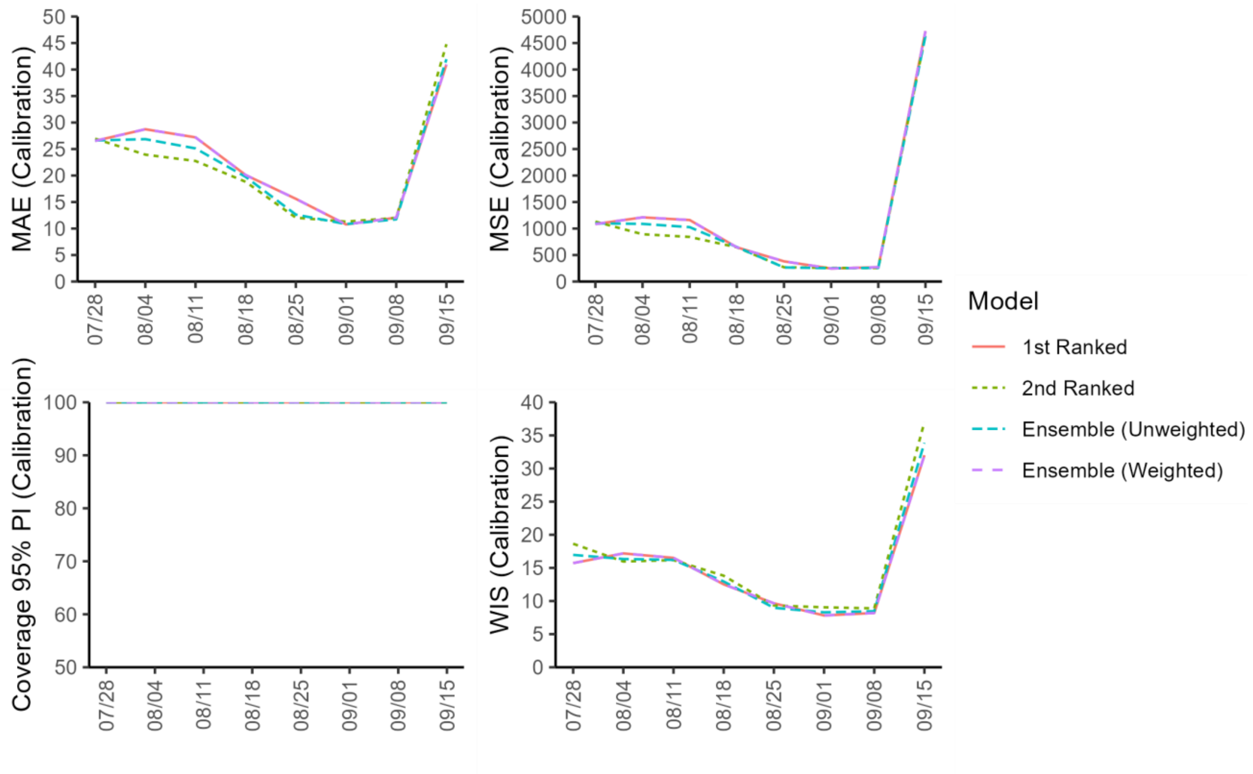


**Figure 31s.** Mean performance metrics for England quantifying model fit across 20 sequential 10-week calibration periods (Week of May 26th – October 13th, 2022) using weekly timeseries data from the OWID team over each 8 forecasting periods**.** Only forecast periods in which observed data was available are included.

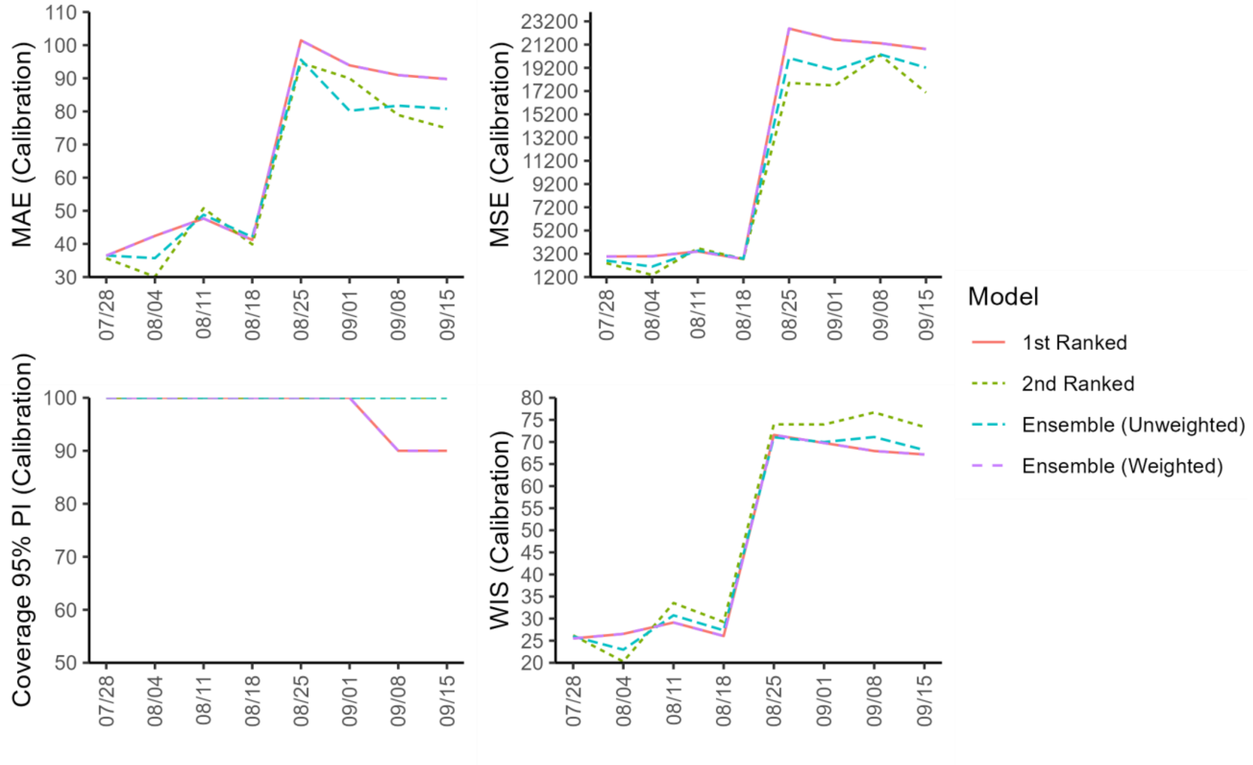


**Figure 32s.** Mean performance metrics for France quantifying model fit across 20 sequential 10-week calibration periods (Week of May 26th – October 13th, 2022) using weekly timeseries data from the OWID team over each 8 forecasting periods**.** Only forecast periods in which observed data was available are included.

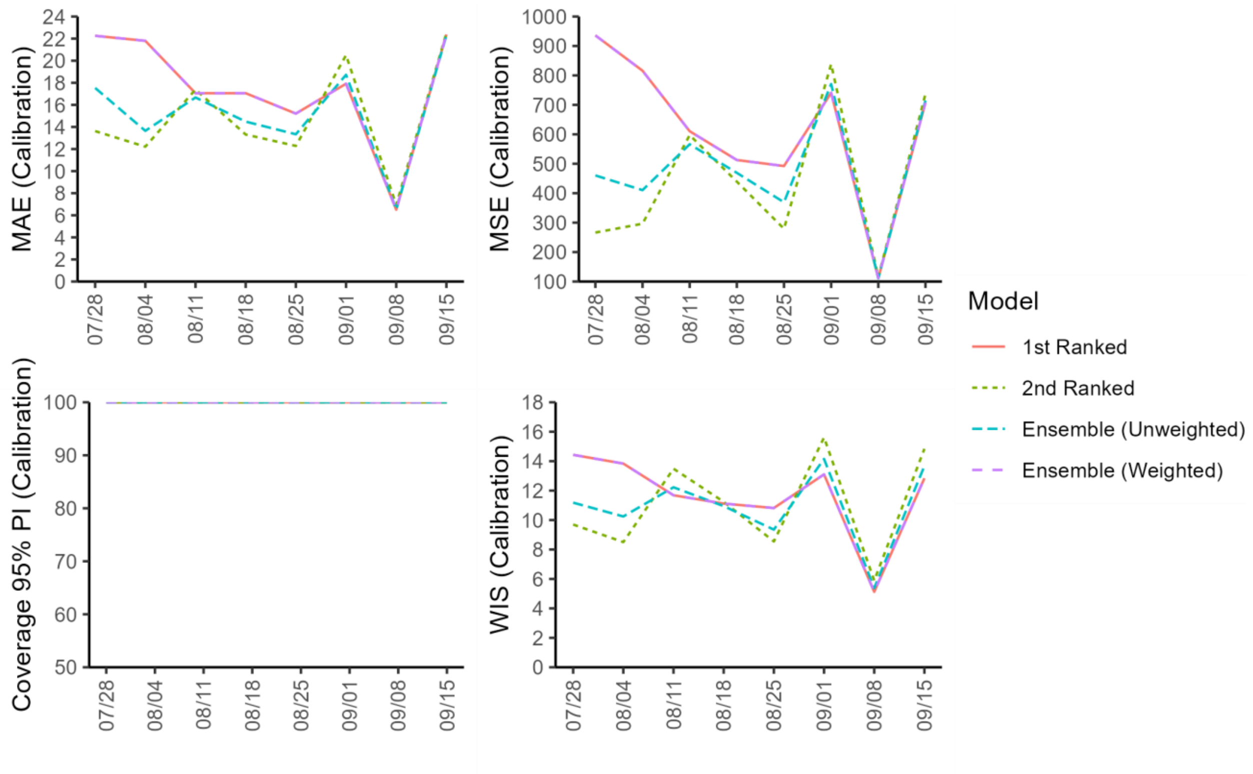


**Figure 33s.** Mean performance metrics for Germany quantifying model fit across 20 sequential 10-week calibration periods (Week of May 26th – October 13th, 2022) using weekly timeseries data from the OWID team over each 8 forecasting periods**.** Only forecast periods in which observed data was available are included.

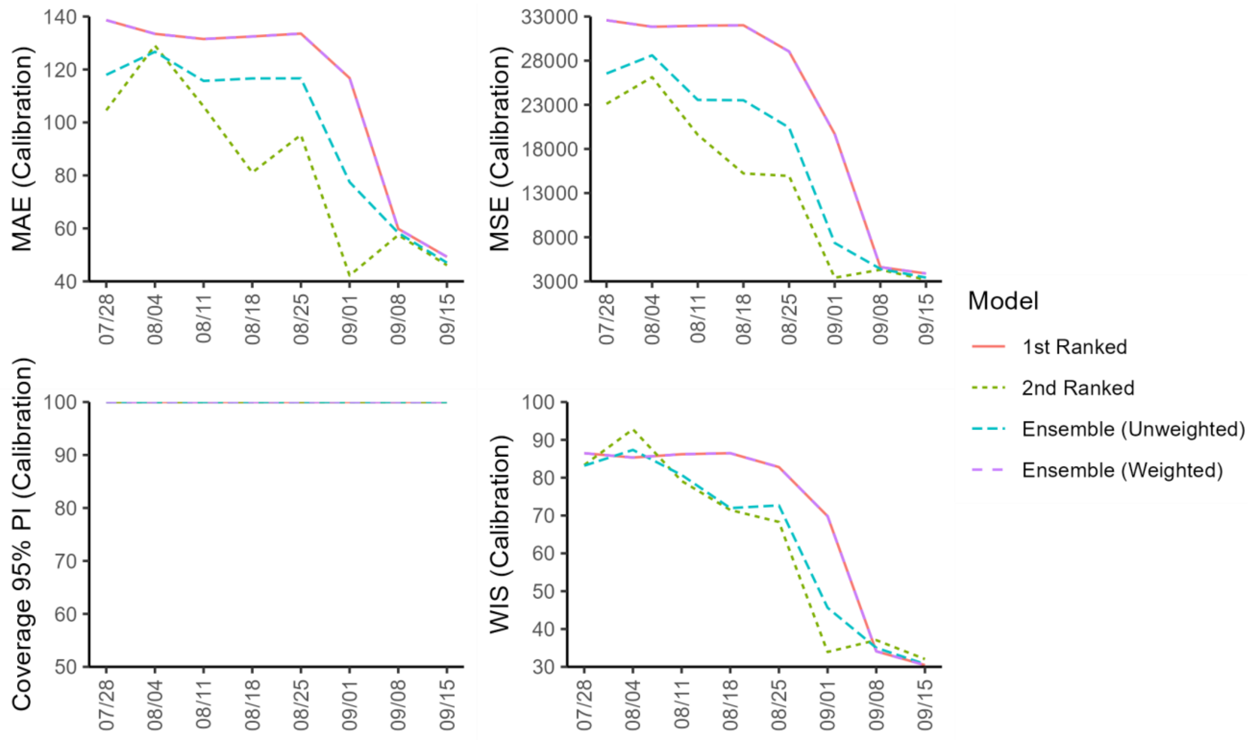


**Figure 34s.** Mean performance metrics for Spain quantifying model fit across 20 sequential 10-week calibration periods (Week of May 26th – October 13th, 2022) using weekly timeseries data from the OWID team over each 8 forecasting periods**.** Only forecast periods in which observed data was available are included.

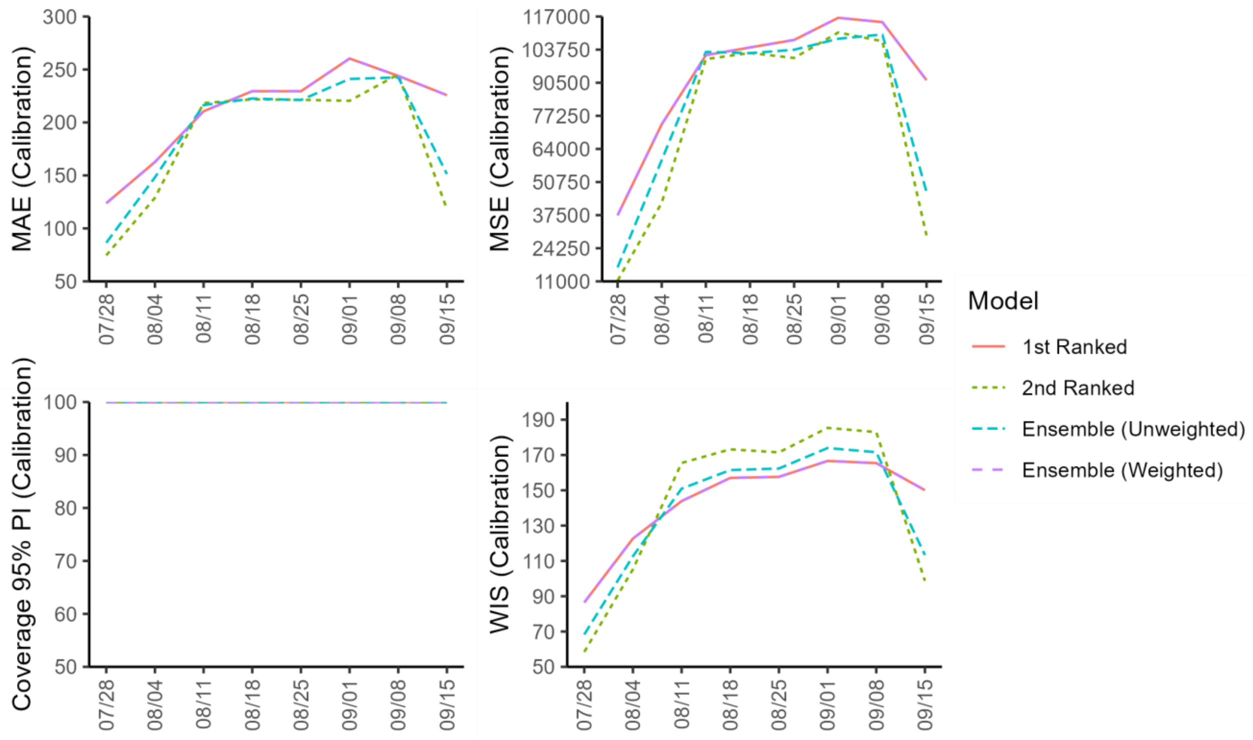


**Figure 35s.** Mean performance metrics for the United States quantifying model fit across 20 sequential 10-week calibration periods (Week of May 26th – October 13th, 2022) using weekly timeseries data from the OWID team over each 8 forecasting periods**.** Only forecast periods in which observed data was available are included.

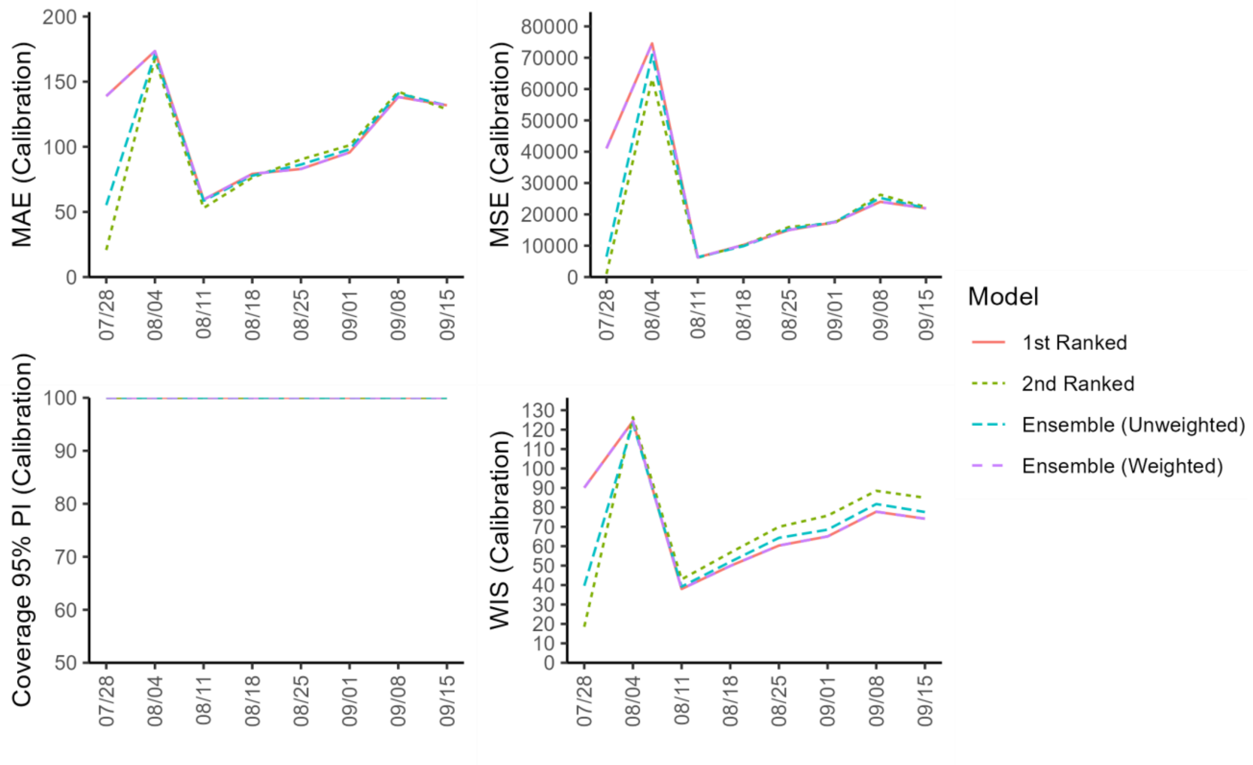


**Figure 36s.** Mean performance metrics for the United States quantifying model fit across 20 sequential 10-week calibration periods (Week of May 26th – October 13th, 2022) using weekly timeseries data from the CDC team over each 8 forecasting periods**.** Only forecast periods in which observed data was available are included.

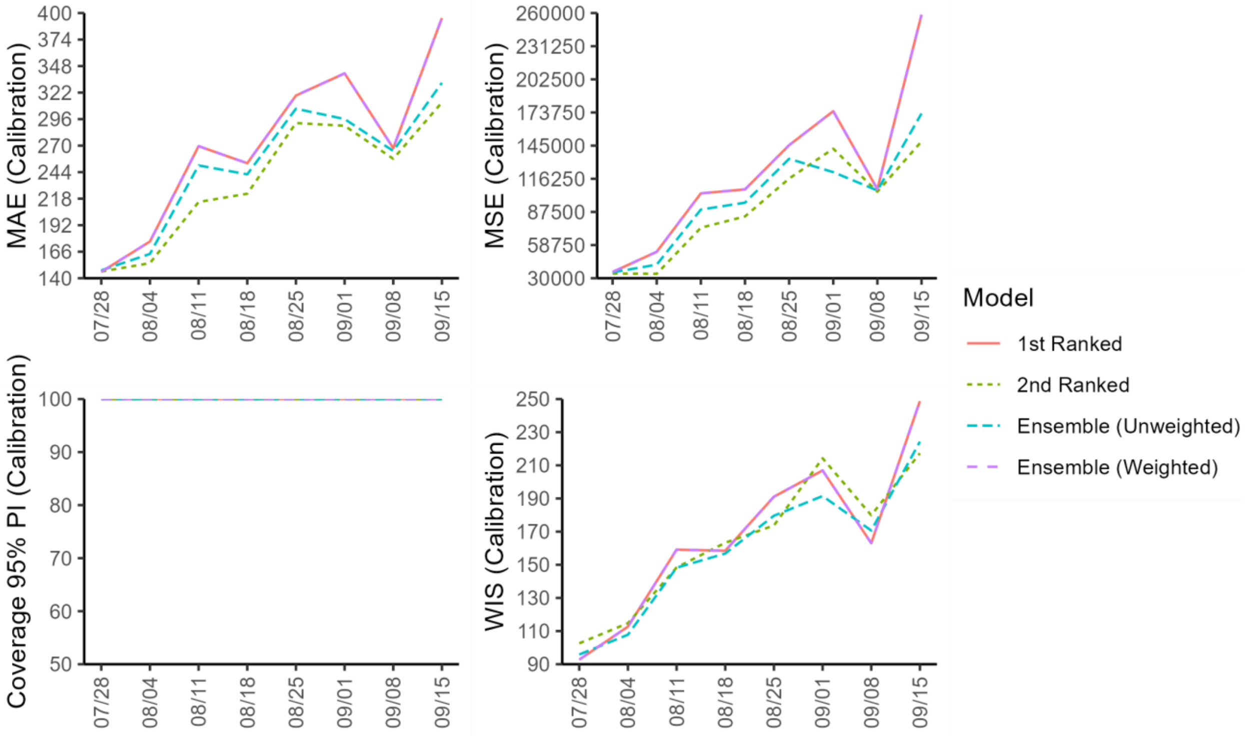


**Figure 37s.** Mean performance metrics for the World quantifying model fit across 20 sequential 10-week calibration periods (Week of May 26th – October 13th, 2022) using weekly timeseries data from the OWID team over each 8 forecasting periods**.** Only forecast periods in which observed data was available are included.

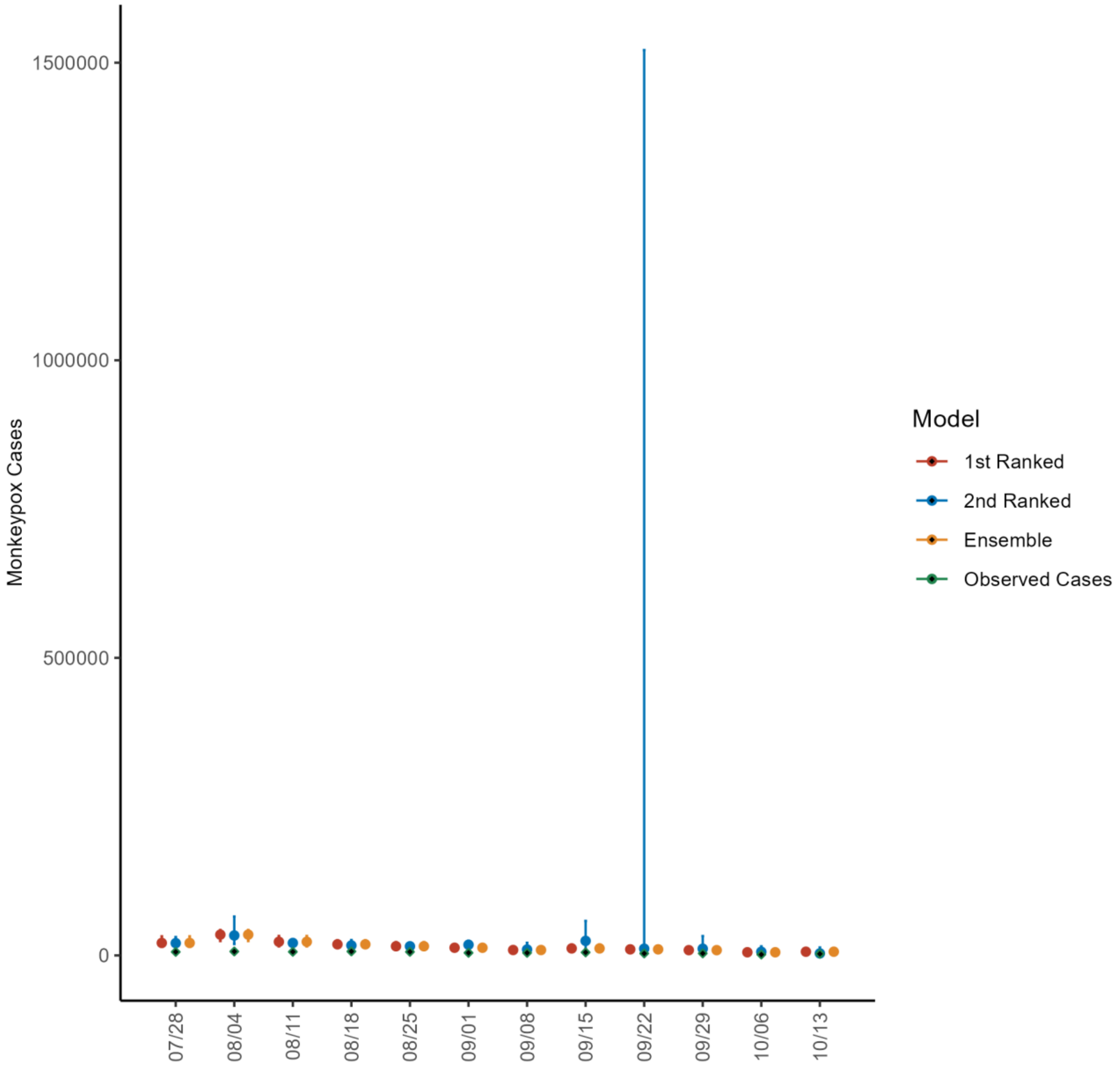


**Figure 38s.** Predicted cumulative number of new monkeypox cases for each of the 4-week ahead forecasts from the weeks of 7/28/2022 through 10/13/2022 based on a 10-week calibration period for the World. The solid circles indicate the estimated cumulative median value, and the bars indicate the corresponding 95% prediction interval. The observed cases are as reported to the OWID team as of 10/21/2022 [25]. For the week of 7/28/2022, data posted by the OWID team on 8/9/2022 was used to produce the forecast as it was the earliest version of data available.

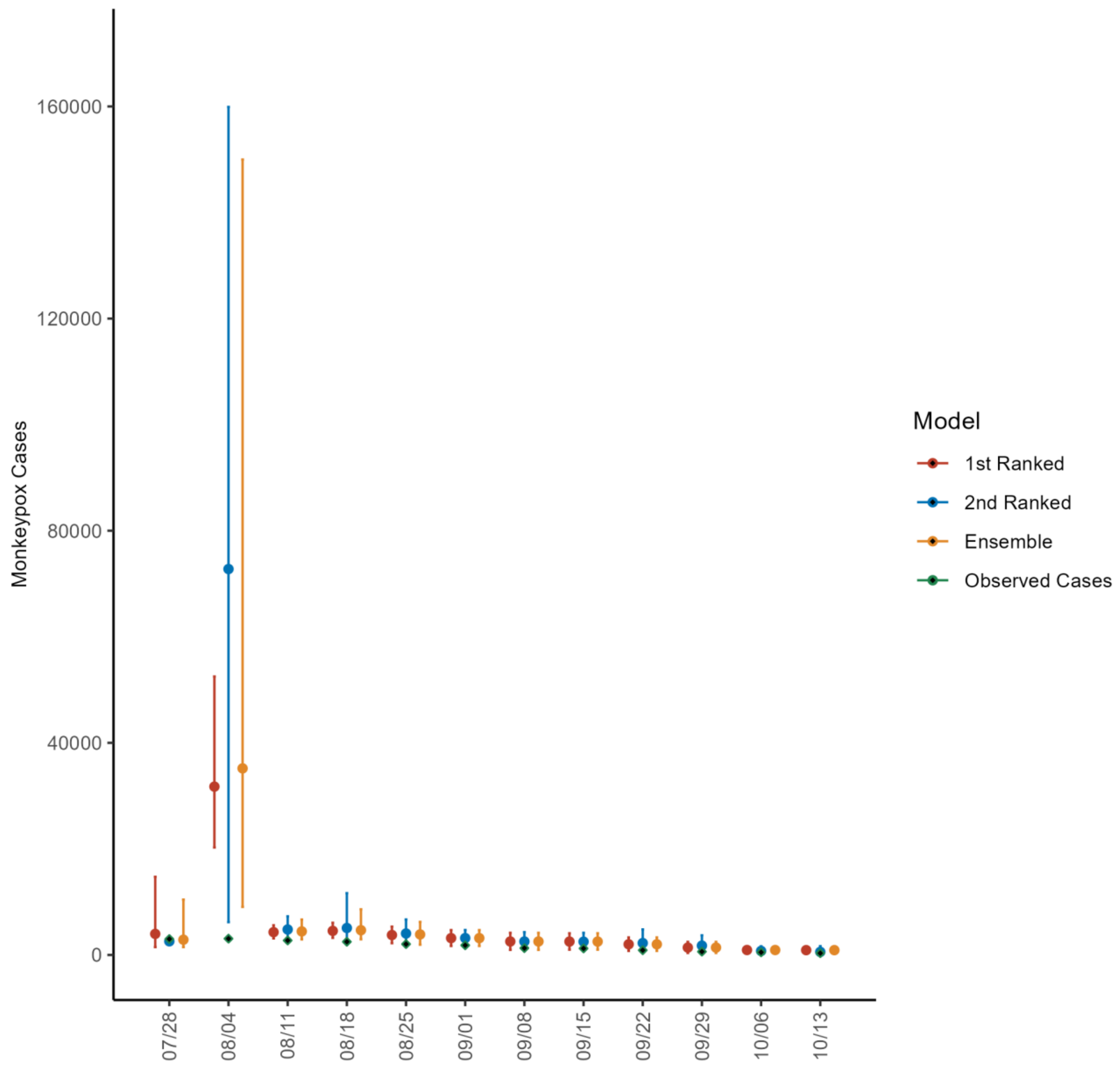


**Figure 39s.** Predicted cumulative number of new monkeypox cases for each of the 4-week ahead forecasts from the weeks of 7/28/2022 through 10/13/2022 based on a 10-week calibration period for the United States. The solid circles indicate the estimated cumulative median value, and the bars indicate the corresponding 95% prediction interval. The observed cases are as reported to the CDC as of 10/19/2022 [24].

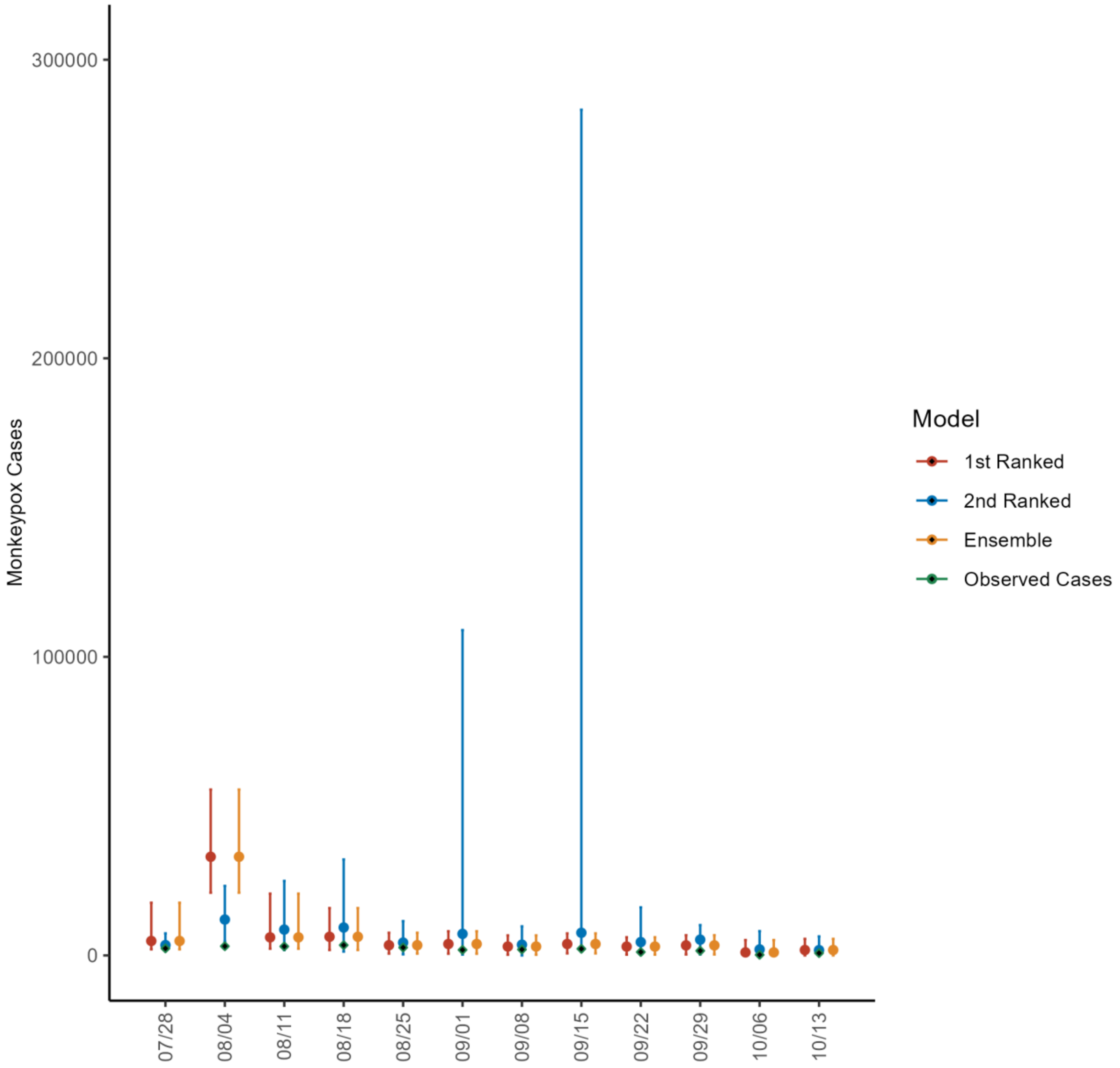


**Figure 40s.** Predicted cumulative number of new monkeypox cases for each of the 4-week ahead forecasts from the weeks of 7/28/2022 through 10/13/2022 based on a 10-week calibration period for the United States. The solid circles indicate the estimated cumulative median value, and the bars indicate the corresponding 95% prediction interval. The observed cases are as reported to the OWID team as of 10/21/2022 [25]. For the week of 7/28/2022, data posted by the OWID team on 8/9/2022 was used to produce the forecast as it was the earliest version of data available.

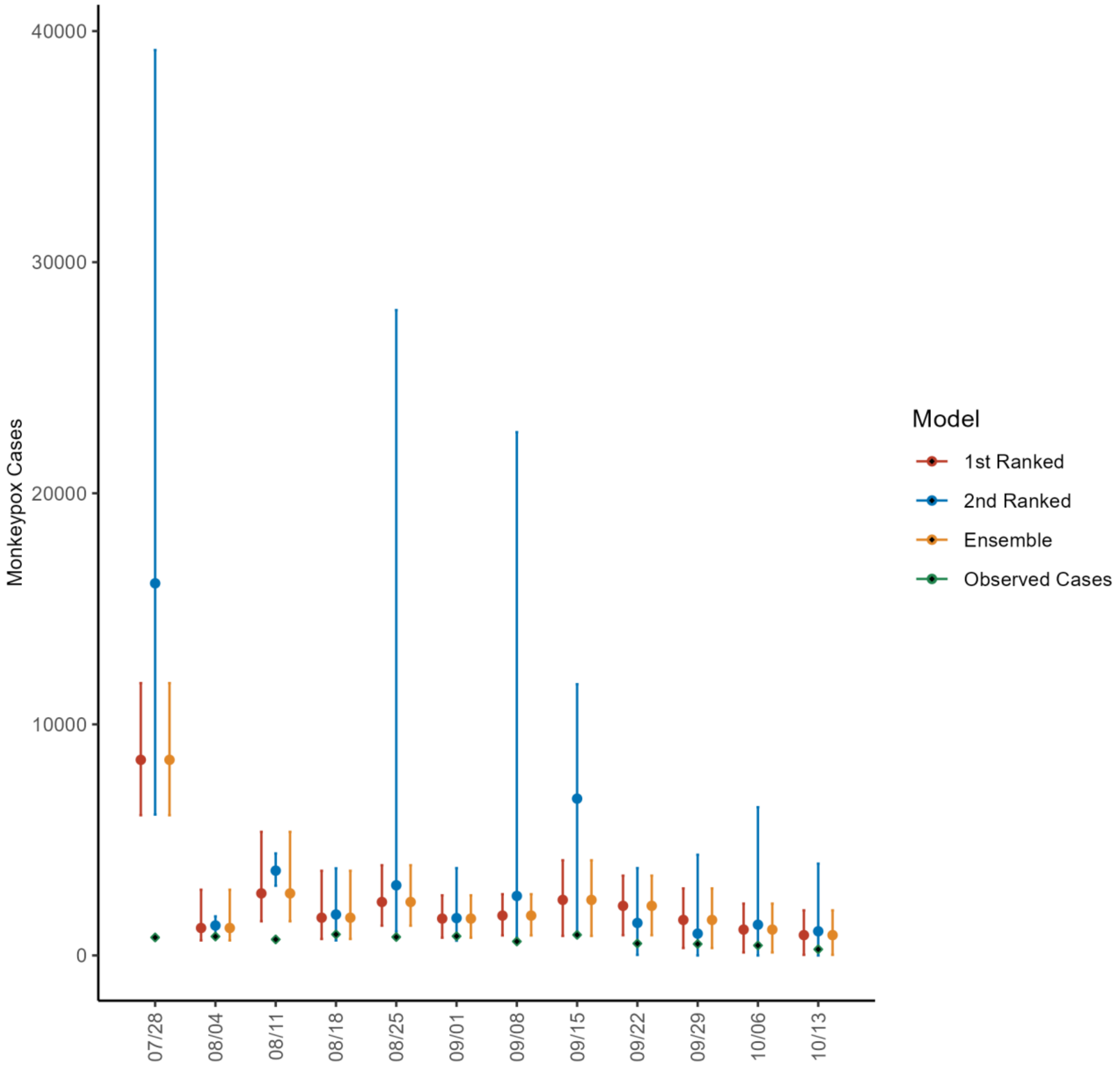


**Figure 41s.** Predicted cumulative number of new monkeypox cases for each of the 4-week ahead forecasts from the weeks of 7/28/2022 through 10/13/2022 based on a 10-week calibration period for Brazil. The solid circles indicate the estimated cumulative median value, and the bars indicate the corresponding 95% prediction interval. The observed cases are as reported to the OWID team as of 10/21/2022 [25]. For the week of 7/28/2022, data posted by the OWID team on 8/9/2022 was used to produce the forecast as it was the earliest version of data available.

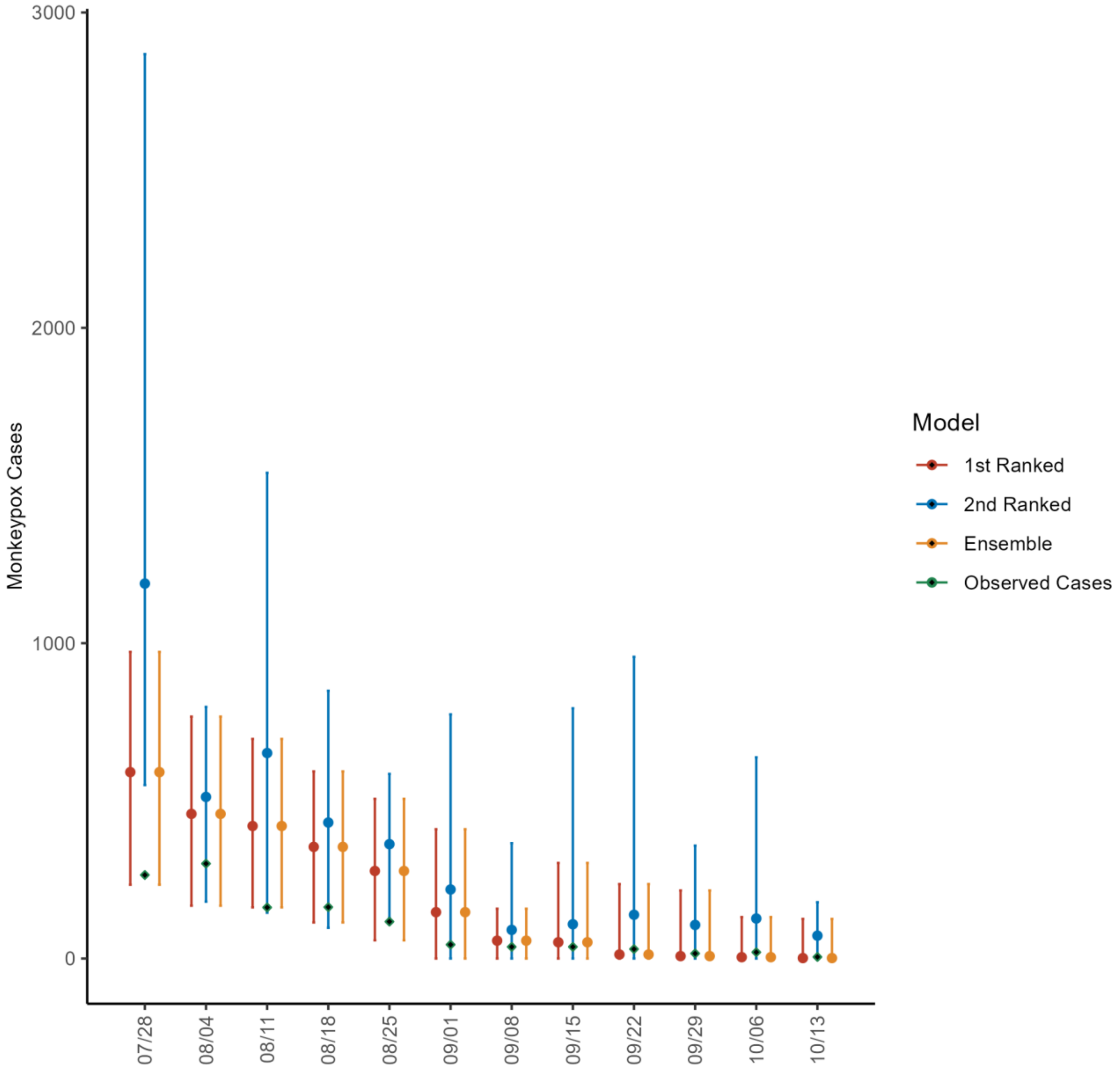


**Figure 42s.** Predicted cumulative number of new monkeypox cases for each of the 4-week ahead forecasts from the weeks of 7/28/2022 through 10/13/2022 based on a 10-week calibration period for Germany. The solid circles indicate the estimated cumulative median value, and the bars indicate the corresponding 95% prediction interval. The observed cases are as reported to the OWID team as of 10/21/2022 [25]. For the week of 7/28/2022, data posted by the OWID team on 8/9/2022 was used to produce the forecast as it was the earliest version of data available.

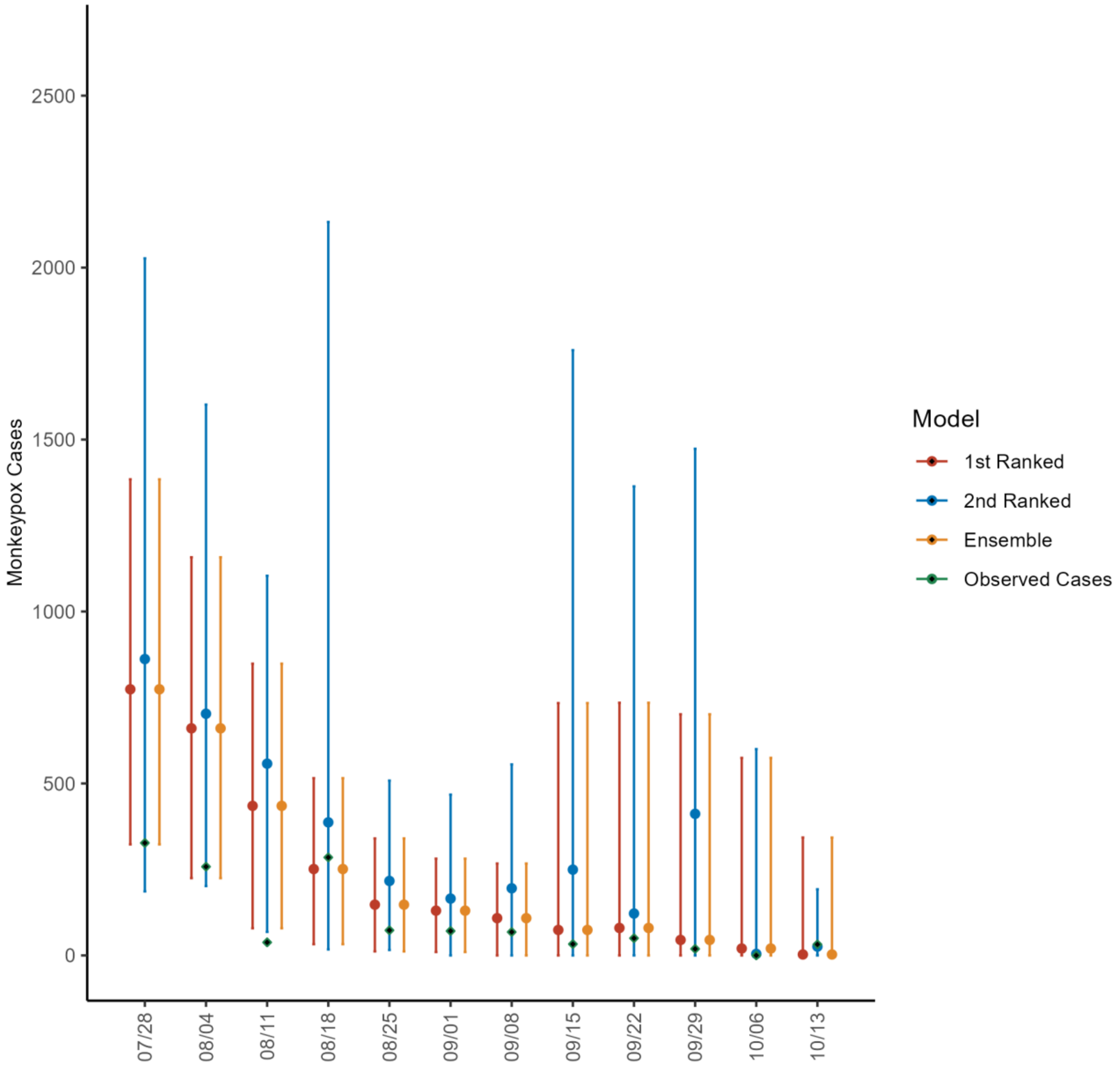


**Figure 43s.** Predicted cumulative number of new monkeypox cases for each of the 4-week ahead forecasts from the weeks of 7/28/2022 through 10/13/2022 based on a 10-week calibration period for England. The solid circles indicate the estimated cumulative median value, and the bars indicate the corresponding 95% prediction interval. The observed cases are as reported to the OWID team as of 10/21/2022 [25]. For the week of 7/28/2022, data posted by the OWID team on 8/9/2022 was used to produce the forecast as it was the earliest version of data available.

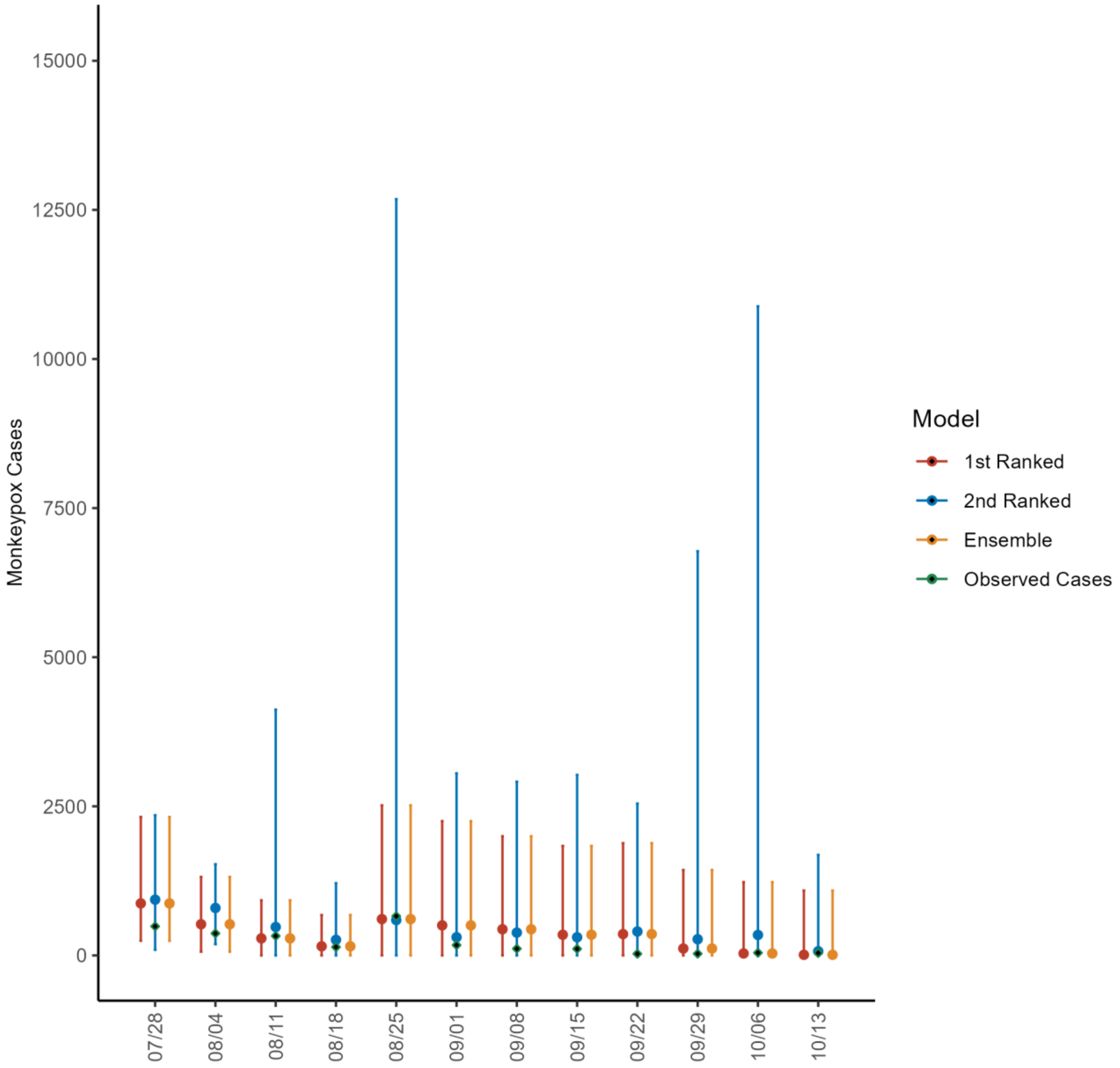


**Figure 44s.** Predicted cumulative number of new monkeypox cases for each of the 4-week ahead forecasts from the weeks of 7/28/2022 through 10/13/2022 based on a 10-week calibration period for France. The solid circles indicate the estimated cumulative median value, and the bars indicate the corresponding 95% prediction interval. The observed cases are as reported to the OWID team as of 10/21/2022 [25]. For the week of 7/28/2022, data posted by the OWID team on 8/9/2022 was used to produce the forecast as it was the earliest version of data available.

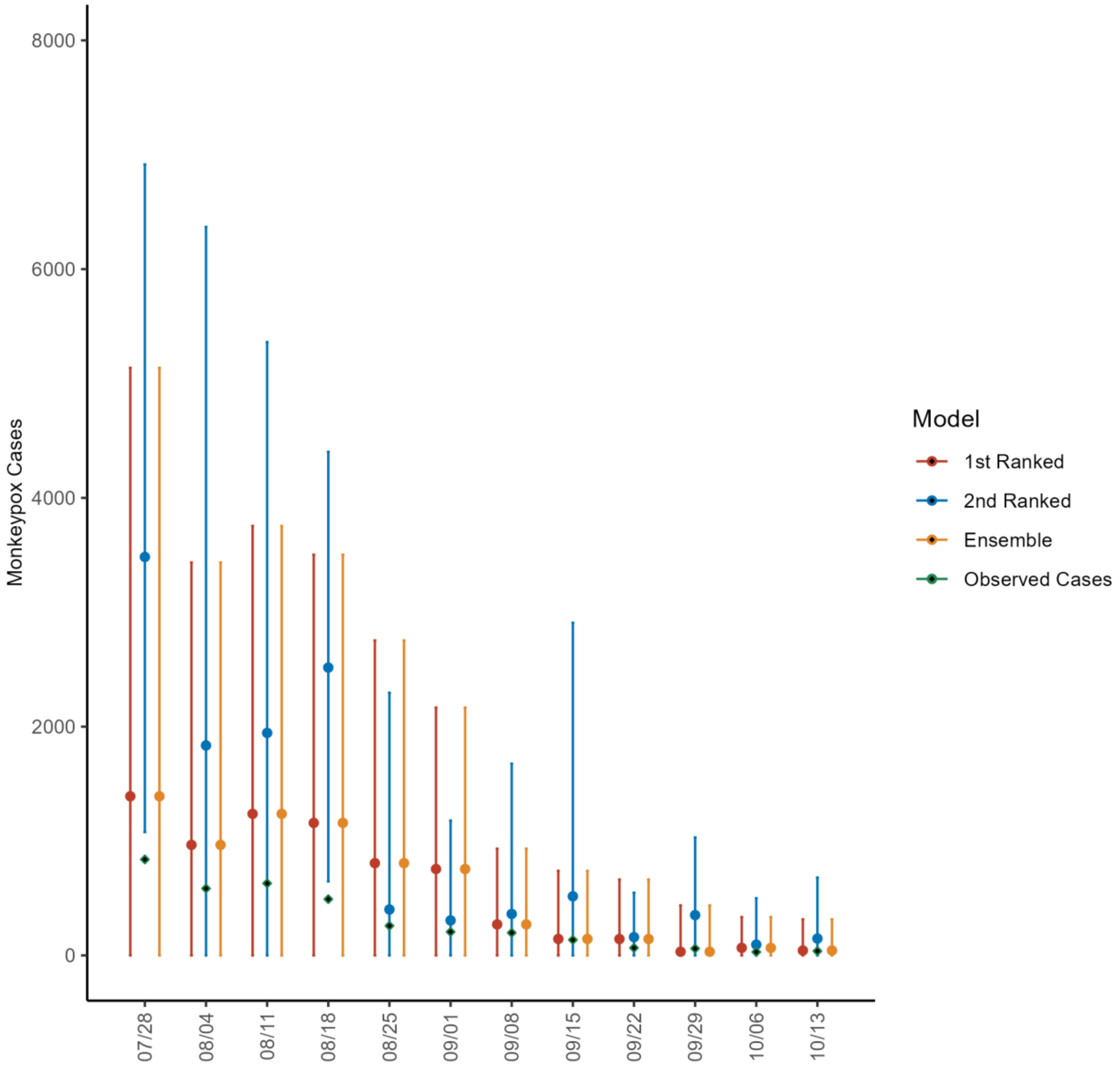


**Figure 45s.** Predicted cumulative number of new monkeypox cases for each of the 4-week ahead forecasts from the weeks of 7/28/2022 through 10/13/2022 based on a 10-week calibration period for Spain. The solid circles indicate the estimated cumulative median value, and the bars indicate the corresponding 95% prediction interval. The observed cases are as reported to the OWID team as of 10/21/2022 [25]. For the week of 7/28/2022, data posted by the OWID team on 8/9/2022 was used to produce the forecast as it was the earliest version of data available.

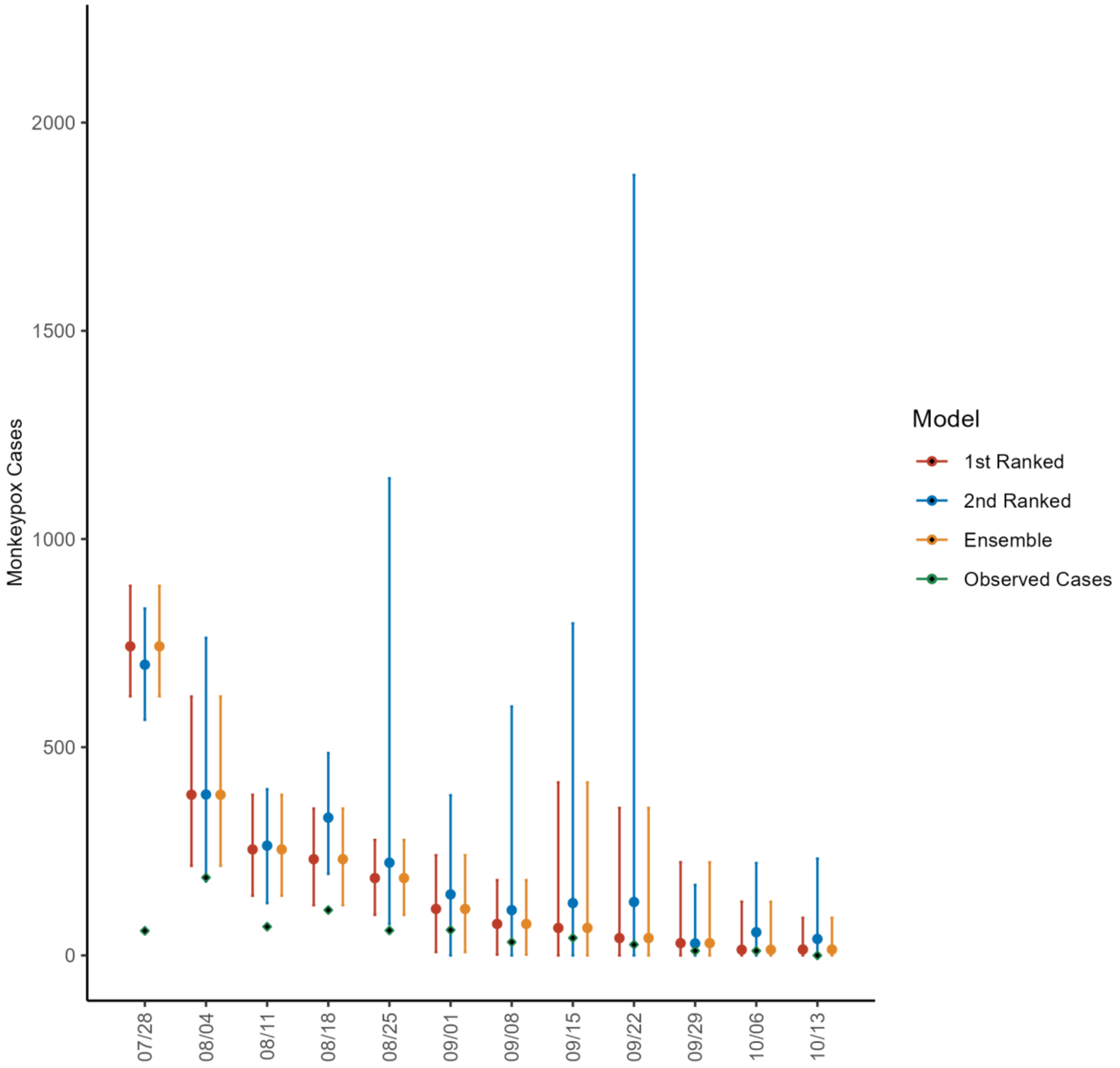


**Figure 46s.** Predicted cumulative number of new monkeypox cases for each of the 4-week ahead forecasts from the weeks of 7/28/2022 through 10/13/2022 based on a 10-week calibration period for Canada. The solid circles indicate the estimated cumulative median value, and the bars indicate the corresponding 95% prediction interval. The observed cases are as reported to the OWID team as of 10/21/2022 [25]. For the week of 7/28/2022, data posted by the OWID team on 8/9/2022 was used to produce the forecast as it was the earliest version of data available.
