## Additional file 2 for "Real-time forecasting the trajectory of monkeypox outbreaks at the national and global levels, July – October 2022"

### Additional file 2: Supplementary tables

| Week | Model | Brazil | Canada | England | France | Germany | Spain | US (OWID) | US (CDC) | World |
|---|---|---|---|---|---|---|---|---|---|---|
| 07-28-22 | 1st ranked | **1307.40** | 90.99 | **87.36** | 174.71 | 40.90 | **176.55** | **1876.66** | **1653.73** | **1152.49** |
| | 2nd ranked | 3228.21 | **82.70** | 91.80 | **172.82** | 114.07 | 341.63 | 2195.80 | 1941.08 | 1241.66 |
| | Ensemble (Weighted) | **1307.40** | 90.99 | **87.36** | 174.71 | 40.90 | **176.55** | **1876.66** | **1653.73** | **1152.49** |
| | Ensemble (Unweighted) | 1501.62 | 86.74 | 88.42 | **172.53** | **29.31** | 236.46 | 2119.44 | 1905.29 | 1170.79 |
| 08-04-22 | 1st ranked | 513.10 | **24.25** | **99.60** | 204.65 | 23.44 | 195.65 | 5524.23 | **5600.15** | 2961.41 |
| | 2nd ranked | **483.25** | 24.47 | 107.57 | **162.16** | **22.90** | **106.63** | **1220.59** | 17132.86 | 2577.23 |
| | Ensemble (Weighted) | 513.10 | **24.25** | **99.60** | 204.65 | 23.44 | 195.65 | 5524.23 | **5600.15** | 2961.41 |
| | Ensemble (Unweighted) | 485.83 | 24.76 | 102.62 | 184.40 | 24.12 | 108.30 | 2724.37 | 6737.27 | **2565.84** |
| 08-11-22 | 1st ranked | 116.98 | **11.86** | **48.94** | 206.31 | **22.16** | **48.18** | 1045.84 | 838.32 | 511.33 |
| | 2nd ranked | 135.49 | 13.76 | 76.46 | **180.87** | 73.47 | 185.03 | **407.53** | **700.86** | 423.42 |
| | Ensemble (Weighted) | 116.98 | **11.86** | **48.94** | 206.31 | **22.16** | **48.18** | 1045.84 | 838.32 | 511.33 |
| | Ensemble (Unweighted) | **102.19** | 12.95 | 60.27 | 196.99 | 30.02 | 102.43 | 742.33 | 775.69 | **384.81** |
| 08-18-22 | 1st ranked | 376.04 | **9.7** | **11.77** | 228.39 | 28.67 | **60.4** | 676.85 | 477.58 | **645.08** |
| | 2nd ranked | **341.39** | 33.19 | 30.48 | **215.18** | 43.62 | 399.73 | 712.06 | **203.62** | 885.96 |
| | Ensemble (Weighted) | 376.04 | **9.7** | **11.77** | 228.39 | **28.67** | **60.4** | 676.85 | 477.58 | **645.08** |
| | Ensemble (Unweighted) | 362.89 | 22.95 | 14.42 | 225.5 | 36.77 | 364.3 | **556.96** | 422.07 | 746.89 |
| 08-25-22 | 1st ranked | 154.1 | **7.09** | 19.13 | 22.54 | **31.17** | 31.84 | 972.14 | 377.49 | 716.45 |
| | 2nd ranked | 133.98 | 15.04 | **10.83** | 31.33 | 54.38 | 74.27 | **821.92** | **292.29** | **696.84** |
| | Ensemble (Weighted) | 154.1 | **7.09** | 19.13 | 22.54 | **31.17** | 31.84 | 972.14 | 377.49 | 716.45 |
| | Ensemble (Unweighted) | **128.79** | 9.5 | 12.95 | **19.16** | 39.88 | **24.53** | 892.96 | 331.98 | 729.9 |
| 09-01-22 | 1st ranked | **226.82** | **7.22** | 11.79 | 37.14 | **6.22** | 73.35 | 776.82 | 208.93 | 823.08 |
| | 2nd ranked | 244.09 | 10.55 | 11.35 | 34.63 | 18.85 | 39.74 | **256.01** | **126.26** | 677.37 |
| | Ensemble (Weighted) | **226.82** | **7.22** | 11.79 | 37.14 | **6.22** | 73.35 | 776.82 | 208.93 | 823.08 |
| | Ensemble (Unweighted) | 237.09 | 7.99 | **11.17** | **16.47** | 9.70 | **13.50** | 621.87 | 184.41 | **418.90** |
| 09-08-22 | 1st ranked | 157.48 | 5.85 | **11.49** | 43.49 | 11.67 | 12.21 | 642.87 | 164.77 | 1120.58 |
| | 2nd ranked | 156.00 | 7.82 | 22.72 | **20.63** | **6.42** | **10.95** | **588.87** | **149.85** | **994.90** |
| | Ensemble (Weighted) | 157.48 | 5.85 | **11.49** | 43.49 | 11.67 | 12.21 | 642.87 | 164.77 | 1120.58 |
| | Ensemble (Unweighted) | **110.68** | **5.83** | 13.52 | 22.76 | 10.37 | 11.30 | 618.69 | 156.78 | 1054.48 |
| 09-15-22 | 1st ranked | **171.27** | **4.09** | 16.82 | 35.83 | 6.28 | 14.54 | **368.93** | **36.40** | **523.75** |
| | 2nd ranked | 1272.59 | 15.74 | 20.83 | **23.62** | **4.55** | 75.21 | 938.93 | 236.66 | 3730.62 |
| | Ensemble (Weighted) | **171.27** | **4.09** | 16.82 | 35.83 | 6.28 | 14.54 | **368.93** | **36.40** | **523.75** |
| | Ensemble (Unweighted) | 337.08 | 5.59 | **15.03** | 25.13 | 5.58 | **8.27** | 404.45 | 91.64 | 1050.04 |

**Table 1s. Mean absolute errors (MAE) metric of the forecasts generated by the sub-epidemic models.** The mean absolute errors (MAE) metric of the forecasts generated by the sub-epidemic models over 8 sequential forecasting periods and for each geographical area.  Values highlighted in bold correspond to best performing model during the forecasting period for a given area. There may be more than one value bolded for a given forecast period and country. This indicates that the bolded models performed equally well. Only forecast periods in which observed data was available are included.

| Week | Model | Brazil | Canada | England | France | Germany | Spain | US (OWID) | US (CDC) | World |
|---|---|---|---|---|---|---|---|---|---|---|
| 7-28-22 | 1st ranked | **2364049.19** | 10320.24 | **11615.61** | 80048.99 | 2506.33 | **39129.18** | **3841147.07** | **2778792.34** | **1597604.21** |
| | 2nd ranked | 16925765.93 | **8276.33** | 12500.64 | **70993.28** | 15632.66 | 137353.23 | 5009878.03 | 3792893.47 | 1804185.50 |
| | Ensemble (Weighted) | **2364049.19** | 10320.24 | **11615.61** | 80048.99 | 2506.33 | **39129.18** | **3841147.07** | **2778792.34** | **1597604.21** |
| | Ensemble (Unweighted) | 3184897.05 | 9230.12 | 11903.15 | 75643.17 | **1151.32** | 71912.88 | 4739459.68 | 3648248.53 | 1617873.05 |
| 8-04-22 | 1st ranked | 322108.04 | 859.00 | **12512.59** | 90471.12 | 617.31 | 43823.72 | 40217736.19 | **38797623.51** | 10893647.67 |
| | 2nd ranked | **258364.19** | **850.35** | 13646.66 | **60875.53** | 678.16 | **16511.82** | **2313218.19** | 457651393.86 | **6650709.10** |
| | Ensemble (Weighted) | 322108.04 | 859.00 | **12512.59** | 90471.12 | 617.31 | 43823.72 | 40217736.19 | **38797623.51** | 10893647.67 |
| | Ensemble (Unweighted) | 265627.4 | 878.62 | 12801.13 | 74574.77 | **593.18** | 16328.29 | 8500892.42 | 57281239.98 | 7048019.09 |
| 8-11-22 | 1st ranked | 20771.72 | **176.02** | **4983.70** | 92537.33 | **742.54** | **2872.34** | 1152686.27 | 711359.42 | 435608.17 |
| | 2nd ranked | 28835.70 | 262.01 | 6517.98 | **77711.78** | 6589.97 | 37471.21 | **203317.81** | **504943.73** | **206499.89** |
| | Ensemble (Weighted) | 20771.72 | **176.02** | **4983.70** | 92537.33 | **742.54** | **2872.34** | 1152686.27 | 711359.42 | 435608.17 |
| | Ensemble (Unweighted) | **18051.77** | 218.02 | 5119.08 | 88428.49 | 1427.14 | 14396.77 | 614876.18 | 609758.03 | 231478.04 |
| 8-18-22 | 1st ranked | 179673.13 | **153.26** | **261.77** | 95708.70 | **1128.73** | **4949.63** | 736308.70 | 245904.12 | **727711.73** |
| | 2nd ranked | **148790.82** | 1195.22 | 1084.46 | **88929.44** | 2369.84 | 161896.62 | 770797.59 | **79446.26** | 1292512.11 |
| | Ensemble (Weighted) | 179673.13 | **153.26** | **261.77** | 95708.70 | **1128.73** | **4949.63** | 736308.70 | 245904.12 | **727711.73** |
| | Ensemble (Unweighted) | 165001.10 | 594.81 | 314.37 | 94223.70 | 1754.56 | 134229.87 | **598252.85** | 200615.58 | 898035.03 |
| 8-25-22 | 1st ranked | 41954.48 | **98.71** | 483.35 | 805.69 | **1168.14** | 1574.15 | 1173259.62 | 152750.92 | 813000.99 |
| | 2nd ranked | 28943.29 | 284.95 | **143.33** | 1538.58 | 2987.99 | 6369.42 | **888425.7711** | **94560.05** | **790633.89** |
| | Ensemble (Weighted) | 41954.48 | **98.71** | 483.35 | 805.69 | **1168.14** | 1574.15 | 1173259.62 | 152750.92 | 813000.99 |
| | Ensemble (Unweighted) | **27364.56** | 151.04 | 287.80 | **403.64** | 1719.40 | **1125.78** | 1015576.84 | 120862.39 | 834190.66 |
| 9-01-22 | 1st ranked | **73078.49** | **61.98** | 250.16 | 1792.28 | **78.89** | 5876.02 | 714440.86 | 56435.89 | 992196.47 |
| | 2nd ranked | 81670.49 | 144.59 | **170.67** | 1772.04 | 399.19 | 2015.50 | **100213.29** | **21634.49** | 497612.85 |
| | Ensemble (Weighted) | **73078.49** | **61.98** | 250.16 | 1792.28 | **78.89** | 5876.02 | 714440.86 | 56435.89 | 992196.47 |
| | Ensemble (Unweighted) | 78784.07 | 80.44 | 220.81 | **510.22** | 143.79 | **264.46** | 493508.74 | 46412.09 | **326677.47** |
| 9-08-22 | 1st ranked | 35058.06 | 58.73 | **182.38** | 2494.38 | 146.86 | 272.99 | 565025.46 | 28528.03 | 1777396.54 |
| | 2nd ranked | 25019.09 | 76.08 | 725.15 | **507.58** | **51.37** | **144.12** | **457688.84** | **23812.47** | **1383510.77** |
| | Ensemble (Weighted) | 35058.06 | 58.73 | **182.38** | 2494.38 | 146.86 | 272.99 | 565025.46 | 28528.03 | 1777396.54 |
| | Ensemble (Unweighted) | **23324.14** | **46.95** | 204.79 | 998.69 | 117.86 | 166.66 | 519936.15 | 25883.95 | 1632965.52 |
| 9-15-22 | 1st ranked | **31512.15** | **23.23** | 380.81 | 1941.18 | 60.25 | 278.07 | **172498.66** | **1708.39** | **395043.07** |
| | 2nd ranked | 1813357.91 | 276.42 | 699.37 | **664.36** | **23.13** | 5720.43 | 1184513.80 | 57656.67 | 15477666.88 |
| | Ensemble (Weighted) | **31512.15** | **23.23** | 380.81 | 1941.18 | 60.25 | 278.07 | **172498.66** | 1708.39 | **395043.07** |
| | Ensemble (Unweighted) | 121858.32 | 44.39 | **260.88** | 1065.67 | 44.77 | **76.74** | 216304.99 | 8734.33 | 1467433.58 |

**Table 2s. Mean square errors (MSE) metric of the forecasts generated by the sub-epidemic models.** The mean square errors (MSE) metric of the forecasts generated by the sub-epidemic models over 8 sequential forecasting periods and for each geographical area. Values highlighted in bold correspond to best performing model during the forecasting period for a given area. There may be more than one value bolded for a given forecast period and country. This indicates that the bolded models performed equally well. Only forecast periods in which observed data was available are included.

| **Week** | **Model** | **Brazil** | **Canada** | **England** | **France** | **Germany** | **Spain** | **US (OWID)** | **US (CDC)** | **World** |
|---|---|---|---|---|---|---|---|---|---|---|
| 07-28-22 | 1st ranked | **0.00** | **25.00** | **75.00** | **75.00** | **100.00** | **100.00** | **100.00** | **75.00** | **100.00** |
| | 2nd ranked | **0.00** | **25.00** | **75.00** | **75.00** | **100.00** | 75.00 | 0.00 | 0.00 | **100.00** |
| | Ensemble (Weighted) | **0.00** | **25.00** | **75.00** | **75.00** | **100.00** | **100.00** | **100.00** | **75.00** | **100.00** |
| | Ensemble (Unweighted) | **0.00** | **25.00** | **75.00** | **75.00** | **100.00** | **100.00** | 75.00 | 0.00 | **100.00** |
| 08-04-22 | 1st ranked | **25.00** | **75.00** | **75.00** | **75.00** | **100.00** | **100.00** | 0.00 | 0.00 | 50.00 |
| | 2nd ranked | 0.00 | **75.00** | **75.00** | **75.00** | **100.00** | **100.00** | **100.00** | **75.00** | **75.00** |
| | Ensemble (Weighted) | **25.00** | **75.00** | **75.00** | **75.00** | **100.00** | **100.00** | 0.00 | 0.00 | 50.00 |
| | Ensemble (Unweighted) | **25.00** | **75.00** | **75.00** | **75.00** | **100.00** | **100.00** | **75.00** | **75.00** | **75.00** |
| 08-11-22 | 1st ranked | **100.00** | **100.00** | 75.00 | 75.00 | **100.00** | **100.00** | **100.00** | 0.00 | **100.00** |
| | 2nd ranked | 75.00 | **100.00** | **100.00** | **100.00** | **100.00** | **100.00** | **100.00** | **25.00** | **100.00** |
| | Ensemble (Weighted) | **100.00** | **100.00** | 75.00 | 75.00 | **100.00** | **100.00** | **100.00** | 0.00 | **100.00** |
| | Ensemble (Unweighted) | **100.00** | **100.00** | 75.00 | **100.00** | **100.00** | **100.00** | **100.00** | **25.00** | **100.00** |
| 08-18-22 | 1st ranked | 75.00 | **100.00** | **100.00** | **75.00** | **100.00** | **100.00** | **100.00** | 25.00 | **100.00** |
| | 2nd ranked | 75.00 | 50.00 | **100.00** | **75.00** | **100.00** | 75.00 | **100.00** | **100.00** | **100.00** |
| | Ensemble (Weighted) | 75.00 | **100.00** | **100.00** | **75.00** | **100.00** | **100.00** | **100.00** | 25.00 | **100.00** |
| | Ensemble (Unweighted) | **100.00** | **100.00** | **100.00** | **75.00** | **100.00** | **100.00** | **100.00** | **100.00** | **100.00** |
| 08-25-22 | 1st ranked | **100.00** | **100.00** | **100.00** | **100.00** | **100.00** | **100.00** | 75.00 | 50.00 | **100.00** |
| | 2nd ranked | **100.00** | **100.00** | **100.00** | **100.00** | 50.00 | **100.00** | **100.00** | **100.00** | **100.00** |
| | Ensemble (Weighted) | **100.00** | **100.00** | **100.00** | **100.00** | **100.00** | **100.00** | 75.00 | 50.00 | **100.00** |
| | Ensemble (Unweighted) | **100.00** | **100.00** | **100.00** | **100.00** | 75.00 | **100.00** | **100.00** | **100.00** | **100.00** |
| 09-01-22 | 1st ranked | 75.00 | **100.00** | **100.00** | **100.00** | **100.00** | **100.00** | 50.00 | **100.00** | 75.00 |
| | 2nd ranked | **100.00** | **100.00** | **100.00** | **100.00** | **100.00** | **100.00** | **100.00** | **100.00** | **100.00** |
| | Ensemble (Weighted) | 75.00 | **100.00** | **100.00** | **100.00** | **100.00** | **100.00** | 50.00 | **100.00** | 75.00 |
| | Ensemble (Unweighted) | 75.00 | **100.00** | **100.00** | **100.00** | **100.00** | **100.00** | **100.00** | **100.00** | **100.00** |
| 09-08-22 | 1st ranked | 75.00 | **100.00** | **100.00** | **100.00** | **100.00** | **100.00** | 50.00 | **100.00** | 50.00 |
| | 2nd ranked | **100.00** | **100.00** | **100.00** | **100.00** | **100.00** | **100.00** | **100.00** | **100.00** | **75.00** |
| | Ensemble (Weighted) | 75.00 | **100.00** | **100.00** | **100.00** | **100.00** | **100.00** | 50.00 | **100.00** | 50.00 |
| | Ensemble (Unweighted) | **100.00** | **100.00** | **100.00** | **100.00** | **100.00** | **100.00** | **100.00** | **100.00** | **75.00** |
| 09-15-22 | 1st ranked | **100.00** | **100.00** | **100.00** | **100.00** | **100.00** | **100.00** | **100.00** | **100.00** | **100.00** |
| | 2nd ranked | **100.00** | **100.00** | **100.00** | **100.00** | **100.00** | **100.00** | **100.00** | **100.00** | 75.00 |
| | Ensemble (Weighted) | **100.00** | **100.00** | **100.00** | **100.00** | **100.00** | **100.00** | **100.00** | **100.00** | **100.00** |
| | Ensemble (Unweighted) | **100.00** | **100.00** | **100.00** | **100.00** | **100.00** | **100.00** | **100.00** | **100.00** | **100.00** |

**Table 3s. Percent coverage of the 95% PI metric of the forecasts generated by the sub-epidemic models.** The percent coverage of the 95% prediction interval (PI) metric of the forecasts generated by the sub-epidemic models over 8 sequential forecasting periods and for each geographical area. Values highlighted in bold correspond to best performing model during the forecasting period for a given area. There may be more than one value bolded for a given forecast period and country. This indicates that the bolded models performed equally well. Only forecast periods in which observed data was available are included.

| Week | Model | Brazil | Canada | England | France | Germany | Spain | US (OWID) | US (CDC) | World |
|---|---|---|---|---|---|---|---|---|---|---|
| 07-28-22 | 1st ranked | **1040.18** | 79.75 | 60.75 | **120.88** | **24.55** | **93.64** | **1107.61** | **1013.06** | **628.79** |
| | 2nd ranked | 2128.96 | **69.82** | 62.24 | 126.69 | 70.97 | 225.15 | 1835.73 | 1891.16 | 720.81 |
| | Ensemble (Weighted) | **1040.18** | 79.75 | 60.75 | **120.88** | **24.55** | **93.64** | **1107.61** | **1013.06** | **628.79** |
| | Ensemble (Unweighted) | 1384.18 | 74.18 | **60.48** | 122.32 | 29.70 | 113.98 | 1376.73 | 1311.82 | 676.83 |
| 08-04-22 | 1st ranked | **394.05** | 16.48 | **67.01** | 155.85 | 13.51 | 91.93 | 4414.97 | **4580.72** | 1915.62 |
| | 2nd ranked | 447.96 | **15.76** | 69.17 | **126.96** | 13.51 | 92.72 | **697.57** | 9284.33 | **1610.64** |
| | Ensemble (Weighted) | **394.05** | 16.48 | **67.01** | 155.85 | 13.51 | 91.93 | 4414.97 | **4580.72** | 1915.62 |
| | Ensemble (Unweighted) | 412.27 | 16.04 | 68.13 | 138.94 | **13.13** | **81.48** | 1498.48 | 5567.13 | 1637.65 |
| 08-11-22 | 1st ranked | 71.68 | **6.91** | **35.46** | 161.83 | **14.44** | **62.34** | 550.30 | 712.04 | 358.12 |
| | 2nd ranked | 96.94 | 8.33 | 42.90 | **116.03** | 43.00 | 115.02 | **287.13** | **479.62** | **244.81** |
| | Ensemble (Weighted) | 71.68 | **6.91** | **35.46** | 161.83 | **14.44** | **62.34** | 550.30 | 712.04 | 358.12 |
| | Ensemble (Unweighted) | **57.72** | 7.68 | 37.68 | 131.32 | 24.28 | 83.68 | 388.21 | 580.48 | 279.50 |
| 08-18-22 | 1st ranked | 255.27 | **6.69** | **9.42** | 177.93 | **18.41** | **66.74** | **386.12** | 339.92 | **405.01** |
| | 2nd ranked | **217.68** | 22.41 | 23.92 | **147.73** | 29.29 | 261.16 | 621.02 | 337.29 | 510.20 |
| | Ensemble (Weighted) | 255.27 | **6.69** | **9.42** | 177.93 | **18.41** | **66.74** | **386.12** | 339.92 | **405.01** |
| | Ensemble (Unweighted) | 234.53 | 11.95 | 14.37 | 161.09 | 22.95 | 141.33 | 430.33 | **270.76** | 446.24 |
| 08-25-22 | 1st ranked | **91.88** | **4.96** | 10.85 | **47.33** | **18.90** | 51.96 | 628.12 | 234.08 | 441.09 |
| | 2nd ranked | 132.93 | 10.40 | **8.37** | 71.68 | 33.98 | **44.06** | **424.17** | **155.30** | **433.50** |
| | Ensemble (Weighted) | **91.88** | **4.96** | 10.85 | **47.33** | **18.90** | 51.96 | 628.12 | 234.08 | 441.09 |
| | Ensemble (Unweighted) | 95.80 | 7.21 | 8.95 | 55.93 | 25.19 | 45.58 | 511.04 | 193.22 | 435.97 |
| 09-01-22 | 1st ranked | 160.56 | **4.69** | **7.77** | 46.40 | **7.88** | 48.60 | 465.91 | 127.59 | 530.15 |
| | 2nd ranked | **144.22** | 7.42 | 8.54 | **43.69** | 15.75 | **26.65** | 466.12 | 127.35 | 392.40 |
| | Ensemble (Weighted) | 160.56 | **4.69** | **7.77** | 46.40 | **7.88** | 48.60 | 465.91 | 127.59 | 530.15 |
| | Ensemble (Unweighted) | 150.66 | 5.74 | **7.77** | 45.07 | 10.83 | 31.94 | **380.37** | **115.15** | **333.79** |
| 09-08-22 | 1st ranked | 100.51 | **4.12** | **7.13** | **41.14** | 6.31 | **17.10** | 399.84 | **91.87** | 814.40 |
| | 2nd ranked | 122.09 | 6.25 | 14.66 | 49.34 | **5.13** | 28.37 | **329.40** | 113.29 | **626.04** |
| | Ensemble (Weighted) | 100.51 | **4.12** | **7.13** | **41.14** | 6.31 | **17.10** | 399.84 | **91.87** | 814.40 |
| | Ensemble (Unweighted) | **89.02** | 4.77 | 9.71 | 45.47 | 5.15 | 21.55 | 363.42 | 94.89 | 698.72 |
| 09-15-22 | 1st ranked | **95.61** | **7.80** | **13.45** | **39.02** | **5.92** | **14.13** | **216.45** | **46.88** | **329.59** |
| | 2nd ranked | 678.89 | 14.18 | 29.16 | 47.79 | 11.11 | 47.46 | 1733.14 | 162.57 | 2137.17 |
| | Ensemble (Weighted) | **95.61** | **7.80** | **13.45** | **39.02** | **5.92** | **14.13** | **216.45** | **46.88** | **329.59** |
| | Ensemble (Unweighted) | 272.38 | 10.66 | 19.78 | 42.34 | 7.96 | 24.93 | 722.95 | 87.13 | 844.38 |

**Table 4s. Weighted Interval Score (WIS) metric of the forecasts generated by the sub-epidemic models.** The weighted Interval Score (WIS) metric of the forecasts generated by the sub-epidemic models over 8 sequential forecasting periods and for each geographical area. Values highlighted in bold correspond to best performing model during the forecasting period for a given area. There may be more than one value bolded for a given forecast period and country. This indicates that the bolded models performed equally well. Only forecast periods in which observed data was available are included.

| | **Model** | **Brazil** | **Canada** | **England** | **France** | **Germany** | **Spain** | **US (OWID)** | **US (CDC)** | **World** |
|---|---|---|---|---|---|---|---|---|---|---|
| Mean Absolute Error (MAE) | 1st ranked* | **37.50** | **75.00** | **62.50** | 0.00 | **50.00** | **37.50** | 25.00 | 37.50 | **37.50** |
| | 2nd ranked | 25.00 | 12.50 | 12.50 | **62.50** | 37.50 | 25.00 | **62.50** | **62.50** | 25.00 |
| | Ensemble* (Weighted) | **37.50** | **75.00** | **62.50** | 0.00 | **50.00** | **37.50** | 25.00 | 37.50 | **37.50** |
| | Ensemble (Unweighted) | **37.50** | 12.50 | 25.00 | 37.50 | 12.50 | **37.50** | 12.50 | 0.00 | **37.50** |
| Mean Square Error (MSE) | 1st ranked* | **37.50** | **62.50** | **62.50** | 0.00 | **50.00** | 37.50 | 25.00 | 37.50 | 37.50 |
| | 2nd ranked | 25.00 | 25.00 | 25.00 | **75.00** | 25.00 | 12.50 | **62.50** | **62.50** | **50.00** |
| | Ensemble* (Weighted) | **37.50** | **62.50** | **62.50** | 0.00 | **50.00** | 37.50 | 25.00 | 37.50 | 37.50 |
| | Ensemble (Unweighted) | **37.50** | 12.50 | 12.50 | 25.00 | 25.00 | **50.00** | 12.50 | 0.00 | 12.50 |
| Percent coverage of the 95% prediction interval (PI) | 1st ranked* | 62.50 | **100.00** | 87.50 | 87.50 | **100.00** | **100.00** | 50.00 | 50.00 | 62.50 |
| | 2nd ranked | 62.50 | 87.50 | **100.00** | **100.00** | 87.50 | 75.00 | **87.50** | **87.50** | 87.50 |
| | Ensemble* (Weighted) | 62.50 | **100.00** | 87.50 | 87.50 | **100.00** | **100.00** | 50.00 | 50.00 | 62.50 |
| | Ensemble (Unweighted) | **87.50** | **100.00** | 87.50 | **100.00** | 87.50 | **100.00** | 75.00 | **87.50** | **100.00** |
| Weighted Interval Score (WIS) | 1st ranked* | **50.00** | **75.00** | **75.00** | **50.00** | **75.00** | **62.50** | 37.50 | **50.00** | 37.50 |
| | 2nd ranked | 25.00 | 25.00 | 12.50 | **50.00** | 12.50 | 25.00 | **50.00** | 25.00 | **50.00** |
| | Ensemble* (Weighted) | **50.00** | **75.00** | **75.00** | **50.00** | **75.00** | **62.50** | 37.50 | **50.00** | 37.50 |
| | Ensemble (Unweighted) | 25.00 | 0.00 | 25.00 | 0.00 | 12.50 | 12.50 | 12.50 | 25.00 | 12.50 |

**Table 5s. Frequency a given model ranked the best for a given metric.** The frequency a given model ranked the best for a given metric generate for forecasts over the course of 8 sequential forecasting periods (July 28th – September 15th, 2022) for geographic regions included in the study. Frequencies are listed as the percent out of 100. *The 1st ranked, and weighted ensemble forecasts were the exact same for every given metric and geographical region.

| | Model | Brazil* | Canada* | England* | France* | Germany* | Spain* | US* (OWID) | US$^{\dagger}$ (CDC) | World* |
|---|---|---|---|---|---|---|---|---|---|---|
| Mean Absolute Error (MAE) | Top-ranked | 45.52 | 9.74 | 22.74 | 67.98 | 17.52 | 111.94 | 210.74 | 112.45 | 270.87 |
| | 2nd ranked | **38.25** | **8.80** | **21.57** | **61.85** | **14.85** | **82.73** | **181.08** | **97.51** | **236.24** |
| | Ensemble (Weighted) | 45.52 | 9.74 | 22.74 | 67.98 | 17.52 | 111.94 | 210.74 | 112.45 | 270.87 |
| | Ensemble (Unweighted) | 41.21 | 9.34 | 21.94 | 62.67 | 15.40 | 97.06 | 191.19 | 102.44 | 250.32 |
| Mean Square Error (MSE) | Top-ranked | 3833.94 | **220.99** | 1215.30 | 12303.24 | 615.95 | 23202.07 | 93478.23 | 26323.77 | 123124.47 |
| | 2nd ranked | **2795.90** | 224.38 | **1118.59** | **10393.44** | **446.15** | **13740.62** | **75456.03** | **20300.88** | **92214.48** |
| | Ensemble (Weighted) | 3833.94 | **220.99** | 1215.30 | 12303.24 | 615.95 | 23202.07 | 93478.23 | 26323.77 | 123124.47 |
| | Ensemble (Unweighted) | 3150.86 | 221.88 | 1159.47 | 11197.14 | 483.93 | 17227.40 | 81207.31 | 21724.52 | 99542.69 |
| Percent coverage of the 95% prediction interval (PI) | Top-ranked | **100.00** | **100.00** | **100.00** | 97.50 | **100.00** | **100.00** | **100.00** | **100.00** | **100.00** |
| | 2nd ranked | **100.00** | **100.00** | **100.00** | **100.00** | **100.00** | **100.00** | **100.00** | **100.00** | **100.00** |
| | Ensemble (Weighted) | **100.00** | **100.00** | **100.00** | 97.50 | **100.00** | **100.00** | **100.00** | **100.00** | **100.00** |
| | Ensemble (Unweighted) | **100.00** | **100.00** | **100.00** | **100.00** | **100.00** | **100.00** | **100.00** | **100.00** | **100.00** |
| Weighted Interval Score (WIS) | Top-ranked | 28.15 | **6.01** | **14.95** | **47.96** | 11.62 | 70.21 | 143.71 | 72.45 | 166.61 |
| | 2nd ranked | 26.98 | 6.53 | 16.11 | 50.91 | 10.98 | **62.26** | 142.61 | 70.46 | 164.27 |
| | Ensemble (Weighted) | 28.15 | **6.01** | **14.95** | **47.96** | 11.62 | 70.21 | 143.71 | 72.45 | 166.61 |
| | Ensemble (Unweighted) | **26.67** | 6.15 | 15.26 | 48.42 | **10.89** | 63.41 | **139.32** | **68.27** | **159.34** |

**Table 6s. Mean performance metrics quantifying model fit.** The mean performance metrics quantifying model fit across 20 sequential 10-week calibration periods (Week of May 26$^{th}$ – October 13$^{th}$, 2022) using weekly timeseries data from the CDC[†] and OWID[*] teams for each geographical region. Values highlighted in bold correspond to best performing model during the forecasting period for a given area. There may be more than one value bolded for a given forecast period and country. This indicates that the bolded models performed equally well. Only forecast periods in which observed data was available are included.

| Week | Model | Brazil | Canada | England | France | Germany | Spain | US (OWID) | US (CDC) | World |
|---|---|---|---|---|---|---|---|---|---|---|
| 07-28-22 | 1st ranked | 21.64 | 6.03 | **26.54** | 36.44 | 22.26 | 138.62 | 123.71 | 138.91 | **146.21** |
| | 2nd ranked | **19.56** | **4.37** | 26.95 | **35.65** | **13.63** | **104.59** | **74.53** | **20.75** | 146.53 |
| | Ensemble (Weighted) | 21.64 | 6.03 | **26.54** | 36.44 | 22.26 | 138.62 | 123.71 | 138.91 | **146.21** |
| | Ensemble (Unweighted) | 20.23 | 5.03 | 26.62 | 36.51 | 17.53 | 117.97 | 86.41 | 55.22 | 147.68 |
| 08-04-22 | 1st ranked | 32.06 | 7.54 | 28.73 | 42.42 | 21.80 | 133.46 | 162.67 | 173.45 | 175.95 |
| | 2nd ranked | **13.08** | **7.39** | **23.94** | **30.07** | **12.21** | 129.05 | **128.86** | **167.53** | **154.76** |
| | Ensemble (Weighted) | 32.06 | 7.54 | 28.73 | 42.42 | 21.80 | 133.46 | 162.67 | 173.45 | 175.95 |
| | Ensemble (Unweighted) | 19.48 | 7.57 | 26.86 | 35.70 | 13.65 | **126.69** | 148.16 | 170.89 | 163.80 |
| 08-11-22 | 1st ranked | 29.97 | 6.75 | 27.21 | **47.67** | 17.05 | 131.51 | **210.55** | 59.41 | 269.48 |
| | 2nd ranked | **25.05** | **5.16** | **22.75** | 50.76 | 17.39 | **106.04** | 218.29 | **52.98** | **214.71** |
| | Ensemble (Weighted) | 29.97 | 6.75 | 27.21 | **47.67** | 17.05 | 131.51 | **210.55** | 59.41 | 269.48 |
| | Ensemble (Unweighted) | 27.27 | 6.05 | 25.12 | 48.81 | **16.67** | 115.69 | 216.37 | 58.57 | 250.58 |
| 08-18-22 | 1st ranked | 36.75 | 7.31 | 20.09 | 41.20 | 17.06 | 132.46 | 229.51 | 79.16 | 252.74 |
| | 2nd ranked | **30.66** | **6.21** | **18.82** | **39.85** | **13.31** | **81.08** | **221.78** | **76.35** | **222.85** |
| | Ensemble (Weighted) | 36.75 | 7.31 | 20.09 | 41.20 | 17.06 | 132.46 | 229.51 | 79.16 | 252.74 |
| | Ensemble (Unweighted) | 33.41 | 6.88 | 19.74 | 42.07 | 14.48 | 116.62 | 222.58 | 77.81 | 241.85 |
| 08-25-22 | 1st ranked | 47.44 | 6.29 | 15.60 | 101.45 | 15.21 | 133.56 | 229.45 | **82.96** | 319.08 |
| | 2nd ranked | **44.26** | **5.12** | **12.04** | **94.66** | **12.28** | **95.30** | 221.37 | 90.28 | **292.14** |
| | Ensemble (Weighted) | 47.44 | 6.29 | 15.60 | 101.45 | 15.21 | 133.56 | 229.45 | **82.96** | 319.08 |
| | Ensemble (Unweighted) | 44.77 | 5.58 | 12.63 | 95.60 | 13.34 | 116.65 | **221.15** | 86.33 | 306.17 |
| 09-01-22 | 1st ranked | 46.08 | 10.37 | **10.77** | 93.91 | **17.92** | 116.77 | 260.35 | **95.67** | 340.84 |
| | 2nd ranked | **42.81** | **9.97** | 11.30 | 89.97 | 20.50 | **42.21** | **220.45** | 101.14 | **289.47** |
| | Ensemble (Weighted) | 46.08 | 10.37 | **10.77** | 93.91 | **17.92** | 116.77 | 260.35 | **95.67** | 340.84 |
| | Ensemble (Unweighted) | 43.42 | 10.15 | 10.84 | **80.17** | 18.73 | 77.48 | 241.06 | 98.09 | 296.05 |
| 09-08-22 | 1st ranked | 55.98 | 8.05 | 12.05 | 90.95 | **6.51** | 59.83 | 243.95 | **138.23** | 267.55 |
| | 2nd ranked | **51.01** | **7.07** | 12.00 | **78.91** | 6.94 | **57.48** | 245.35 | 142.29 | **257.11** |
| | Ensemble (Weighted) | 55.98 | 8.05 | 12.05 | 90.95 | **6.51** | 59.83 | 243.95 | **138.23** | 267.55 |
| | Ensemble (Unweighted) | 52.96 | 7.70 | **11.73** | 81.74 | 6.53 | 58.32 | **242.55** | 140.89 | 264.78 |
| 09-15-22 | 1st ranked | 94.28 | 25.60 | **40.95** | 89.78 | 22.38 | 49.27 | 225.75 | 131.85 | 395.12 |
| | 2nd ranked | **79.60** | **25.08** | 44.78 | **74.89** | 22.51 | **46.09** | **118.04** | **128.79** | **312.35** |
| | Ensemble (Weighted) | 94.28 | 25.60 | **40.95** | 89.78 | 22.38 | 49.27 | 225.75 | 131.85 | 395.12 |
| | Ensemble (Unweighted) | 88.13 | 25.78 | 41.95 | 80.77 | **22.30** | 47.05 | 151.23 | 131.69 | 331.61 |

**Table 7s. Mean absolute error (MAE) metrics quantifying model fit.** The mean absolute error (MAE) metrics quantifying model fit across 20 sequential 10-week calibration periods (Week of May 26$^{th}$ – October 13$^{th}$, 2022) used during forecasting periods for the given geographical regions. Values highlighted in bold correspond to best performing model during the forecasting period for a given area. There may be more than one value bolded for a given forecast period and country. This indicates that the bolded models performed equally well. Only forecast periods in which observed data was available are included.

| Week | Model | Brazil | Canada | England | France | Germany | Spain | US (OWID) | US (CDC) | World |
|---|---|---|---|---|---|---|---|---|---|---|
| 07-28-22 | 1st ranked | 797.18 | 69.02 | **1084.19** | 2964.07 | 936.01 | 32577.00 | 37426.62 | 41061.16 | 35401.61 |
| | 2nd ranked | **517.01** | **38.92** | 1130.97 | **2396.74** | **266.33** | **23107.15** | **11310.19** | **993.61** | **34107.69** |
| | Ensemble (Weighted) | 797.18 | 69.02 | **1084.19** | 2964.07 | 936.01 | 32577.00 | 37426.62 | 41061.16 | 35401.61 |
| | Ensemble (Unweighted) | 638.21 | 50.24 | 1097.57 | 2609.83 | 460.37 | 26537.40 | 16583.00 | 6469.11 | 35066.30 |
| 08-04-22 | 1st ranked | 1778.58 | 89.70 | 1210.71 | 2986.61 | 815.99 | 31839.75 | 73858.07 | 74580.14 | 52914.01 |
| | 2nd ranked | **328.41** | **87.18** | **892.64** | **1377.14** | **296.36** | **26130.86** | **42452.64** | **63253.40** | **33827.46** |
| | Ensemble (Weighted) | 1778.58 | 89.70 | 1210.71 | 2986.61 | 815.99 | 31839.75 | 73858.07 | 74580.14 | 52914.01 |
| | Ensemble (Unweighted) | 598.69 | 90.25 | 1087.66 | 2097.18 | 410.45 | 28603.31 | 59515.87 | 70969.79 | 41772.18 |
| 08-11-22 | 1st ranked | 2037.12 | 64.00 | 1160.27 | **3400.90** | 610.48 | 31954.23 | 101526.93 | 6266.19 | 103584.14 |
| | 2nd ranked | **1029.36** | **45.50** | **841.49** | 3673.42 | 597.38 | **19598.69** | **99956.47** | **6178.18** | **73762.56** |
| | Ensemble (Weighted) | 2037.12 | 64.00 | 1160.27 | **3400.90** | 610.48 | 31954.23 | 101526.93 | 6266.19 | 103584.14 |
| | Ensemble (Unweighted) | 1415.44 | 53.21 | 1026.27 | 3475.13 | **566.17** | 23555.35 | 102836.07 | 6335.19 | 89546.09 |
| 08-18-22 | 1st ranked | 2180.36 | 74.93 | **645.32** | **2746.61** | 512.74 | 32008.76 | 104611.47 | 10241.78 | 107034.19 |
| | 2nd ranked | **1976.88** | **62.10** | 645.74 | 2753.31 | **439.24** | **15225.73** | 102405.02 | 10154.03 | **83580.23** |
| | Ensemble (Weighted) | 2180.36 | 74.93 | **645.32** | **2746.61** | 512.74 | 32008.76 | 104611.47 | 10241.78 | 107034.19 |
| | Ensemble (Unweighted) | 2035.91 | 67.27 | 651.19 | 2789.54 | 469.14 | 23514.26 | **102339.35** | **9852.72** | 95498.10 |
| 08-25-22 | 1st ranked | 3629.89 | 58.78 | 380.95 | 22578.97 | 492.25 | 29041.11 | 107625.64 | **14990**.15 | 145361.68 |
| | 2nd ranked | **2632.41** | **53.06** | 268.86 | **17902.65** | **279.30** | **14942.30** | **100406.00** | 15976.68 | **116437.94** |
| | Ensemble (Weighted) | 3629.89 | 58.78 | 380.95 | 22578.97 | 492.25 | 29041.11 | 107625.64 | **14990.15** | 145361.68 |
| | Ensemble (Unweighted) | 3001.05 | 54.66 | **265.87** | 20042.38 | 369.12 | 20419.05 | 103700.44 | 15331.37 | 133545.45 |
| 09-01-22 | 1st ranked | 3622.06 | **147.75** | **245.07** | 21616.42 | **740.96** | 19695.80 | 116488.17 | 17498.43 | 174865.61 |
| | 2nd ranked | **2933.55** | 151.40 | 257.32 | **17678.92** | 838.36 | **3412.73** | 110619.95 | **17271.81** | 142329.80 |
| | Ensemble (Weighted) | 3622.06 | **147.75** | **245.07** | 21616.42 | **740.96** | 19695.80 | 116488.17 | 17498.43 | 174865.61 |
| | Ensemble (Unweighted) | 3056.94 | 149.02 | 251.09 | 18995.35 | 770.86 | 7351.48 | **108100.31** | 17604.84 | **121988.23** |
| 09-08-22 | 1st ranked | 4172.76 | 101.17 | 270.48 | 21313.98 | 111.68 | 4612.78 | 114765.99 | **24014.42** | 107287.31 |
| | 2nd ranked | **3745.92** | **96.00** | **254.58** | **20296.42** | 114.99 | **4316.43** | **107104.49** | 26269.55 | **105033.78** |
| | Ensemble (Weighted) | 4172.76 | 101.17 | 270.48 | 21313.98 | 111.68 | 4612.78 | 114765.99 | **24014.42** | 107287.31 |
| | Ensemble (Unweighted) | 3822.69 | 98.69 | 257.00 | 20353.34 | **110.65** | 4413.96 | 109812.89 | 25256.66 | 106099.55 |
| 09-15-22 | 1st ranked | 12453.56 | **1162.51** | 4725.45 | 20818.35 | **707.49** | 3887.13 | 91522.97 | **21937.91** | 258547.18 |
| | 2nd ranked | **9203.64** | 1260.88 | 4657.13 | **17068.89** | 737.19 | **3191.04** | **29393.45** | 22309.78 | **148636.39** |
| | Ensemble (Weighted) | 12453.56 | **1162.51** | 4725.45 | 20818.35 | **707.49** | 3887.13 | 91522.97 | **21937.91** | 258547.18 |
| | Ensemble (Unweighted) | 10637.98 | 1211.67 | **4639.07** | 19214.38 | 714.72 | 3424.40 | 46770.58 | 21976.45 | 172825.59 |

**Table 8s. Mean square error (MSE) metrics quantifying model fit.** The mean square error (MSE) metrics quantifying model fit across 20 sequential 10-week calibration periods (Week of May 26th – October 13th, 2022) used during forecasting periods for the given geographical regions. Values highlighted in bold correspond to best performing model during the forecasting period for a given area. There may be more than one value bolded for a given forecast period and country. This indicates that the bolded models performed equally well. Only forecast periods in which observed data was available are included.

| Week | Model | Brazil | Canada | England | France | Germany | Spain | US (OWID) | US (CDC) | World |
|---|---|---|---|---|---|---|---|---|---|---|
| 07-28-22 | 1st ranked | **100.00** | **100.00** | **100.00** | **100.00** | **100.00** | **100.00** | **100.00** | **100.00** | **100.00** |
| | 2nd ranked | **100.00** | **100.00** | **100.00** | **100.00** | **100.00** | **100.00** | **100.00** | **100.00** | **100.00** |
| | Ensemble (Weighted) | **100.00** | **100.00** | **100.00** | **100.00** | **100.00** | **100.00** | **100.00** | **100.00** | **100.00** |
| | Ensemble (Unweighted) | **100.00** | **100.00** | **100.00** | **100.00** | **100.00** | **100.00** | **100.00** | **100.00** | **100.00** |
| 08-04-22 | 1st ranked | **100.00** | **100.00** | **100.00** | **100.00** | **100.00** | **100.00** | **100.00** | **100.00** | **100.00** |
| | 2nd ranked | **100.00** | **100.00** | **100.00** | **100.00** | **100.00** | **100.00** | **100.00** | **100.00** | **100.00** |
| | Ensemble (Weighted) | **100.00** | **100.00** | **100.00** | **100.00** | **100.00** | **100.00** | **100.00** | **100.00** | **100.00** |
| | Ensemble (Unweighted) | **100.00** | **100.00** | **100.00** | **100.00** | **100.00** | **100.00** | **100.00** | **100.00** | **100.00** |
| 08-11-22 | 1st ranked | **100.00** | **100.00** | **100.00** | **100.00** | **100.00** | **100.00** | **100.00** | **100.00** | **100.00** |
| | 2nd ranked | **100.00** | **100.00** | **100.00** | **100.00** | **100.00** | **100.00** | **100.00** | **100.00** | **100.00** |
| | Ensemble (Weighted) | **100.00** | **100.00** | **100.00** | **100.00** | **100.00** | **100.00** | **100.00** | **100.00** | **100.00** |
| | Ensemble (Unweighted) | **100.00** | **100.00** | **100.00** | **100.00** | **100.00** | **100.00** | **100.00** | **100.00** | **100.00** |
| 08-18-22 | 1st ranked | **100.00** | **100.00** | **100.00** | **100.00** | **100.00** | **100.00** | **100.00** | **100.00** | **100.00** |
| | 2nd ranked | **100.00** | **100.00** | **100.00** | **100.00** | **100.00** | **100.00** | **100.00** | **100.00** | **100.00** |
| | Ensemble (Weighted) | **100.00** | **100.00** | **100.00** | **100.00** | **100.00** | **100.00** | **100.00** | **100.00** | **100.00** |
| | Ensemble (Unweighted) | **100.00** | **100.00** | **100.00** | **100.00** | **100.00** | **100.00** | **100.00** | **100.00** | **100.00** |
| 08-25-22 | 1st ranked | **100.00** | **100.00** | **100.00** | **100.00** | **100.00** | **100.00** | **100.00** | **100.00** | **100.00** |
| | 2nd ranked | **100.00** | **100.00** | **100.00** | **100.00** | **100.00** | **100.00** | **100.00** | **100.00** | **100.00** |
| | Ensemble (Weighted) | **100.00** | **100.00** | **100.00** | **100.00** | **100.00** | **100.00** | **100.00** | **100.00** | **100.00** |
| | Ensemble (Unweighted) | **100.00** | **100.00** | **100.00** | **100.00** | **100.00** | **100.00** | **100.00** | **100.00** | **100.00** |
| 09-01-22 | 1st ranked | **100.00** | **100.00** | **100.00** | **100.00** | **100.00** | **100.00** | **100.00** | **100.00** | **100.00** |
| | 2nd ranked | **100.00** | **100.00** | **100.00** | **100.00** | **100.00** | **100.00** | **100.00** | **100.00** | **100.00** |
| | Ensemble (Weighted) | **100.00** | **100.00** | **100.00** | **100.00** | **100.00** | **100.00** | **100.00** | **100.00** | **100.00** |
| | Ensemble (Unweighted) | **100.00** | **100.00** | **100.00** | **100.00** | **100.00** | **100.00** | **100.00** | **100.00** | **100.00** |
| 09-08-22 | 1st ranked | **100.00** | **100.00** | **100.00** | 90.00 | **100.00** | **100.00** | **100.00** | **100.00** | **100.00** |
| | 2nd ranked | **100.00** | **100.00** | **100.00** | **100.00** | **100.00** | **100.00** | **100.00** | **100.00** | **100.00** |
| | Ensemble (Weighted) | **100.00** | **100.00** | **100.00** | 90.00 | **100.00** | **100.00** | **100.00** | **100.00** | **100.00** |
| | Ensemble (Unweighted) | **100.00** | **100.00** | **100.00** | **100.00** | **100.00** | **100.00** | **100.00** | **100.00** | **100.00** |
| 09-15-22 | 1st ranked | **100.00** | **100.00** | **100.00** | 90.00 | **100.00** | **100.00** | **100.00** | **100.00** | **100.00** |
| | 2nd ranked | **100.00** | **100.00** | **100.00** | **100.00** | **100.00** | **100.00** | **100.00** | **100.00** | **100.00** |
| | Ensemble (Weighted) | **100.00** | **100.00** | **100.00** | 90.00 | **100.00** | **100.00** | **100.00** | **100.00** | **100.00** |
| | Ensemble (Unweighted) | **100.00** | **100.00** | **100.00** | **100.00** | **100.00** | **100.00** | **100.00** | **100.00** | **100.00** |

**Table 9s. Percent coverage of the 95% (PI) metrics quantifying model fit.** The percent coverage of the 95% prediction interval (PI) metrics quantifying model fit across 20 sequential 10-week calibration periods (Week of May 26th – October 13th, 2022) used during forecasting periods for the given geographical regions. Values highlighted in bold correspond to best performing model during the forecasting period for a given area. There may be more than one value bolded for a given forecast period and country. This indicates that the bolded models performed equally well. Only forecast periods in which observed data was available are included.

| Week | Model | Brazil | Canada | England | France | Germany | Spain | US (OWID) | US (CDC) | World |
|---|---|---|---|---|---|---|---|---|---|---|
| 07-28-22 | 1st ranked | **13.38** | 4.02 | **15.72** | **25.51** | 14.43 | 86.47 | 86.45 | 90.11 | **92.74** |
| | 2nd ranked | 14.76 | **3.51** | 18.65 | 26.21 | **9.70** | 83.34 | **58.45** | **18.52** | 102.60 |
| | Ensemble (Weighted) | **13.38** | 4.02 | **15.72** | **25.51** | 14.43 | 86.47 | 86.45 | 90.11 | **92.74** |
| | Ensemble (Unweighted) | 13.57 | 3.66 | 16.96 | 25.98 | 11.18 | **83.17** | 68.32 | 39.64 | 95.85 |
| 08-04-22 | 1st ranked | 19.76 | **4.78** | 17.19 | 26.54 | 13.84 | **85.33** | 122.64 | 124.34 | 112.68 |
| | 2nd ranked | **11.32** | 5.22 | **15.96** | **20.27** | **8.50** | 92.79 | **105.21** | 126.40 | 114.76 |
| | Ensemble (Weighted) | 19.76 | **4.78** | 17.19 | 26.54 | 13.84 | **85.33** | 122.64 | 124.34 | 112.68 |
| | Ensemble (Unweighted) | 13.52 | 4.93 | 16.34 | 22.97 | 10.24 | 87.34 | 112.64 | **123.26** | **107.80** |
| 08-11-22 | 1st ranked | 20.97 | 3.99 | 16.51 | **29.12** | **11.69** | 86.20 | **143.87** | **37.99** | 159.13 |
| | 2nd ranked | **17.14** | **3.76** | **16.17** | 33.54 | 13.49 | **79.14** | 165.48 | 42.95 | 148.37 |
| | Ensemble (Weighted) | 20.97 | 3.99 | 16.51 | **29.12** | **11.69** | 86.20 | **143.87** | **37.99** | 159.13 |
| | Ensemble (Unweighted) | 18.08 | 3.81 | 16.21 | 30.74 | 12.23 | 80.72 | 150.92 | 39.19 | **148.23** |
| 08-18-22 | 1st ranked | **23.72** | 4.29 | **12.52** | **26.06** | 11.12 | 86.47 | **157.02** | **49.85** | 158.50 |
| | 2nd ranked | 25.52 | 4.40 | 13.82 | 29.24 | 11.24 | **71.47** | 173.19 | 56.66 | 163.29 |
| | Ensemble (Weighted) | **23.72** | 4.29 | **12.52** | **26.06** | 11.12 | 86.47 | **157.02** | **49.85** | 158.50 |
| | Ensemble (Unweighted) | 24.25 | **4.25** | 12.95 | 27.34 | **10.95** | 71.97 | 161.47 | 51.94 | **156.79** |
| 08-25-22 | 1st ranked | 29.30 | **3.80** | 9.66 | 71.59 | 10.82 | 82.80 | **157.60** | **60.34** | 191.08 |
| | 2nd ranked | 29.30 | 4.17 | 9.33 | 73.98 | **8.55** | **68.30** | 171.48 | 69.97 | **173.72** |
| | Ensemble (Weighted) | 29.30 | **3.80** | 9.66 | 71.59 | 10.82 | 82.80 | **157.60** | **60.34** | 191.08 |
| | Ensemble (Unweighted) | **28.40** | 3.91 | **8.98** | **71.07** | 9.35 | 72.68 | 162.33 | 64.31 | 179.59 |
| 09-01-22 | 1st ranked | 29.73 | **6.02** | **7.82** | **69.77** | **13.10** | 69.82 | **166.63** | **65.03** | 206.93 |
| | 2nd ranked | **28.80** | 6.72 | 9.05 | 73.95 | 15.61 | **33.94** | 185.35 | 75.72 | 214.27 |
| | Ensemble (Weighted) | 29.73 | **6.02** | **7.82** | **69.77** | **13.10** | 69.82 | **166.63** | **65.03** | 206.93 |
| | Ensemble (Unweighted) | 28.92 | 6.29 | 8.29 | 69.98 | 14.15 | 45.63 | 173.94 | 68.50 | **191.47** |
| 09-08-22 | 1st ranked | **32.31** | **4.97** | **8.19** | **67.95** | **5.13** | **34.11** | **165.41** | **77.75** | **163.05** |
| | 2nd ranked | 34.34 | 5.52 | 8.89 | 76.69 | 5.89 | 37.05 | 182.97 | 88.51 | 179.88 |
| | Ensemble (Weighted) | **32.31** | **4.97** | **8.19** | **67.95** | **5.13** | **34.11** | **165.41** | **77.75** | **163.05** |
| | Ensemble (Unweighted) | 32.49 | 5.16 | 8.46 | 71.12 | 5.38 | 35.04 | 171.57 | 81.72 | 170.66 |
| 09-15-22 | 1st ranked | 56.03 | **16.20** | **32.01** | **67.17** | **12.84** | **30.46** | 150.05 | **74.15** | 248.77 |
| | 2nd ranked | 54.65 | 18.92 | 37.02 | 73.39 | 14.84 | 32.05 | **98.72** | 84.95 | **217.31** |
| | Ensemble (Weighted) | 56.03 | **16.20** | **32.01** | **67.17** | **12.84** | **30.46** | 150.05 | **74.15** | 248.77 |
| | Ensemble (Unweighted) | **54.12** | 17.21 | 33.88 | 68.16 | 13.67 | 30.73 | 113.33 | 77.60 | 224.32 |

**Table 10s. Weighted interval score (WIS) metrics quantifying model fit.** The weighted interval score (WIS) metrics quantifying model fit across 20 sequential 10-week calibration periods (Week of May 26th – October 13th, 2022) used during forecasting periods for the given geographical regions. Values highlighted in bold correspond to best performing model during the forecasting period for a given area. There may be more than one value bolded for a given forecast period and country. This indicates that the bolded models performed equally well. Only forecast periods in which observed data was available are included.

| Second-Ranked Model | | | | | | | | | | |
|---|---|---|---|---|---|---|---|---|---|---|
| **Week** | | **Brazil** | **Canada** | **England** | **France** | **Germany** | **Spain** | **US (OWID)** | **US (CDC)** | **World** |
| 07-28-22 | Median | 16102.9 | 698.1 | 861.7 | 935.2 | 1189.0 | 3483.8 | 3448.7 | 2573.8 | 20604.8 |
| | LB | 6096.9 | 565.7 | 186.1 | 93.8 | 550.1 | 1078.3 | 1679.7 | 1921.9 | 13073.0 |
| | UB | 39180.3 | 833.1 | 2026.9 | 2350.8 | 2868.0 | 6915.3 | 7389.5 | 3134.8 | 30896.7 |
| 08-04-22 | Median | 1295.2 | 386.4 | 702.7 | 795.3 | 512.5 | 1835.7 | 12011.9 | 72771.3 | 33511.9 |
| | LB | 925.5 | 177.4 | 201.7 | 187.8 | 180.4 | 0.0 | 4640.2 | 6186.8 | 19185.0 |
| | UB | 1689.7 | 762.7 | 1601.6 | 1529.4 | 797.9 | 6369.0 | 23275.5 | 159904.1 | 65275.2 |
| 08-11-22 | Median | 3668.9 | 263.6 | 557.5 | 476.7 | 651.7 | 1945.2 | 8652.7 | 4803.5 | 20910.2 |
| | LB | 3017.8 | 125.6 | 68.4 | 0.0 | 145.3 | 0.0 | 1790.0 | 2688.0 | 15426.7 |
| | UB | 4412.5 | 399.1 | 1103.9 | 4121.0 | 1540.3 | 5362.3 | 24943.3 | 7276.3 | 27280.5 |
| 08-18-22 | Median | 1770.8 | 330.8 | 386.9 | 261.3 | 431.2 | 2516.3 | 9344.5 | 5083.5 | 16790.7 |
| | LB | 650.9 | 196.0 | 17.4 | 0.0 | 97.5 | 647.2 | 1288.9 | 2617.6 | 10550.5 |
| | UB | 3767.3 | 485.9 | 2132.6 | 1210.2 | 849.1 | 4402.7 | 32128.1 | 11648.3 | 25842.3 |
| 08-25-22 | Median | 3034.9 | 222.7 | 216.5 | 592.4 | 362.7 | 401.5 | 4296.7 | 4047.7 | 15331.8 |
| | LB | 1037.6 | 75.4 | 15.8 | 0.0 | 128.4 | 0.0 | 369.0 | 1682.3 | 10176.1 |
| | UB | 27926.0 | 1145.7 | 508.3 | 12679.9 | 585.5 | 2296.1 | 11480.4 | 6690.7 | 21986.0 |
| 09-01-22 | Median | 1615.7 | 146.5 | 165.2 | 305.3 | 219.0 | 305.9 | 7213.8 | 3184.7 | 17921.9 |
| | LB | 638.5 | 0.0 | 0.0 | 0.0 | 0.0 | 0.0 | 279.2 | 1740.5 | 11869.3 |
| | UB | 3777.4 | 384.7 | 467.5 | 3053.3 | 774.3 | 1179.2 | 108959.8 | 4702.4 | 24638.6 |
| 09-08-22 | Median | 2572.8 | 108.8 | 195.2 | 382.4 | 90.8 | 362.6 | 3592.6 | 2540.1 | 9916.2 |
| | LB | 765.2 | 0.0 | 0.0 | 0.0 | 0.0 | 0.0 | 0.0 | 943.9 | 4243.7 |
| | UB | 22643.0 | 597.8 | 555.4 | 2912.5 | 365.8 | 1676.5 | 9732.7 | 4298.7 | 21199.2 |
| 09-15-22 | Median | 6784.5 | 125.7 | 249.4 | 304.4 | 108.8 | 516.6 | 7584.4 | 2502.8 | 24432.4 |
| | LB | 1031.7 | 0.0 | 0.0 | 0.0 | 0.0 | 0.0 | 1426.7 | 940.1 | 10430.3 |
| | UB | 11736.4 | 797.4 | 1759.9 | 3026.6 | 793.5 | 2908.5 | 283241.3 | 4173.2 | 58026.3 |
| 09-22-22 | Median | 1404.9 | 128.3 | 121.8 | 400.8 | 138.8 | 159.9 | 4464.6 | 2242.4 | 11317.7 |
| | LB | 23.0 | 0.0 | 0.0 | 0.0 | 0.0 | 0.0 | 102.9 | 411.5 | 1876.8 |
| | UB | 3777.2 | 1874.2 | 1363.8 | 2546.3 | 956.8 | 548.2 | 16090.2 | 4805.1 | 1521252.0 |
| 09-29-22 | Median | 942.9 | 28.7 | 411.7 | 271.7 | 106.6 | 352.6 | 5275.8 | 1759.7 | 11260.9 |
| | LB | 0.0 | 0.0 | 0.0 | 0.0 | 0.0 | 0.0 | 317.9 | 227.2 | 1405.9 |
| | UB | 4356.1 | 169.3 | 1473.2 | 6776.6 | 358.0 | 1031.4 | 10158.9 | 3703.1 | 32533.4 |
| 10-06-22 | Median | 1328.6 | 55.7 | 4.4 | 343.6 | 126.9 | 94.6 | 2005.9 | 756.3 | 6075.6 |
| | LB | 0.0 | 0.0 | 0.0 | 0.0 | 0.0 | 0.0 | 0.0 | 84.3 | 0.0 |
| | UB | 6414.1 | 221.7 | 599.9 | 10883.7 | 638.0 | 500.0 | 8133.4 | 1627.7 | 15271.4 |
| 10-13-22 | Median | 1044.6 | 39.5 | 25.7 | 70.0 | 72.5 | 147.5 | 1768.7 | 579.2 | 3393.7 |
| | LB | 0.0 | 0.0 | 0.0 | 0.0 | 0.0 | 0.0 | 0.0 | 0.0 | 0.0 |
| | UB | 3970.7 | 232.6 | 192.3 | 1686.1 | 178.7 | 681.0 | 6361.3 | 1652.4 | 13340.1 |

**Table 11s. Predicted cumulative number of newly forecasted monkeypox cases and 95% PIs for the second-ranked model.** Predicted cumulative number of newly forecasted monkeypox cases and 95% PIs for each of the 4-week ahead forecasts during the weeks of 7-28-2022 through 10-13-2022 based on a 10-week calibration period for the second-ranked sub-epidemic model.

| Weighted - Ensemble Model | | | | | | | | | | |
|---|---|---|---|---|---|---|---|---|---|---|
| Week | | **Brazil** | **Canada** | **England** | **France** | **Germany** | **Spain** | **US (OWID)** | **US (CDC)** | **World** |
| 07-28-22 | Median | 8463.7 | 742.3 | 773.9 | 872.4 | 591.2 | 1391.2 | 4838.8 | 2874.5 | 20957.3 |
| | LB | 6067.5 | 622.3 | 322.9 | 245.6 | 234.1 | 0.0 | 2038.2 | 1484.1 | 14263.4 |
| | UB | 11783.6 | 887.6 | 1384.3 | 2322.0 | 972.4 | 5137.4 | 17645.5 | 10437.6 | 32121.5 |
| 08-04-22 | Median | 1187.2 | 385.9 | 660.3 | 522.6 | 458.7 | 966.3 | 33042.3 | 35181.3 | 34936.8 |
| | LB | 651.6 | 215.0 | 224.7 | 62.0 | 167.6 | 0.0 | 21005.3 | 9062.0 | 24148.0 |
| | UB | 2841.9 | 621.8 | 1157.7 | 1316.6 | 767.2 | 3435.6 | 55553.7 | 149984.9 | 42706.4 |
| 08-11-22 | Median | 2685.3 | 254.5 | 434.9 | 286.1 | 420.3 | 1237.7 | 6058.4 | 4442.5 | 22930.0 |
| | LB | 1477.3 | 143.3 | 79.0 | 0.0 | 162.3 | 0.0 | 2242.7 | 2943.4 | 15153.2 |
| | UB | 5348.1 | 385.7 | 848.3 | 924.7 | 696.3 | 3754.3 | 20689.6 | 6676.0 | 32717.9 |
| 08-18-22 | Median | 1631.6 | 231.2 | 251.2 | 152.8 | 354.1 | 1158.6 | 6247.6 | 4667.2 | 18643.0 |
| | LB | 711.3 | 120.6 | 32.6 | 0.0 | 114.4 | 0.0 | 1816.2 | 2955.4 | 12818.4 |
| | UB | 3662.6 | 352.7 | 515.4 | 677.8 | 593.4 | 3503.7 | 15853.4 | 8601.9 | 25447.7 |
| 08-25-22 | Median | 2312.1 | 185.7 | 147.6 | 609.1 | 277.7 | 806.8 | 3420.0 | 3871.9 | 15412.5 |
| | LB | 1288.4 | 97.3 | 11.7 | 0.0 | 57.8 | 0.0 | 610.6 | 1938.8 | 9456.3 |
| | UB | 3903.2 | 277.4 | 340.1 | 2517.0 | 506.4 | 2753.8 | 7594.1 | 6232.0 | 22458.5 |
| 09-01-22 | Median | 1593.3 | 111.7 | 130.1 | 503.9 | 147.1 | 755.3 | 3824.4 | 3175.5 | 12899.5 |
| | LB | 768.7 | 8.0 | 9.9 | 0.0 | 0.0 | 0.0 | 571.0 | 1698.0 | 7794.2 |
| | UB | 2599.4 | 240.7 | 281.3 | 2252.0 | 410.0 | 2166.7 | 8101.4 | 4687.9 | 18540.1 |
| 09-08-22 | Median | 1723.7 | 75.6 | 108.4 | 438.6 | 56.7 | 271.0 | 2978.7 | 2542.6 | 9097.0 |
| | LB | 866.6 | 1.9 | 0.0 | 0.0 | 0.0 | 0.0 | 223.6 | 959.7 | 5279.6 |
| | UB | 2648.9 | 180.9 | 267.2 | 1998.6 | 158.0 | 933.0 | 6706.9 | 4174.2 | 13323.9 |
| 09-15-22 | Median | 2401.9 | 66.0 | 74.0 | 346.6 | 51.6 | 143.9 | 3826.9 | 2526.5 | 11764.0 |
| | LB | 844.3 | 0.0 | 0.0 | 0.0 | 0.0 | 0.0 | 690.5 | 999.4 | 5704.9 |
| | UB | 4116.9 | 415.6 | 733.7 | 1836.3 | 303.1 | 739.6 | 7379.2 | 4087.7 | 18545.3 |
| 09-22-22 | Median | 2144.9 | 41.7 | 79.9 | 357.9 | 12.8 | 143.3 | 2958.6 | 2011.8 | 10205.4 |
| | LB | 875.0 | 0.0 | 0.0 | 0.0 | 0.0 | 0.0 | 265.3 | 768.4 | 4305.2 |
| | UB | 3452.4 | 354.2 | 734.6 | 1882.9 | 236.1 | 663.5 | 6078.7 | 3299.8 | 16332.0 |
| 09-29-22 | Median | 1534.7 | 29.5 | 45.0 | 115.6 | 7.7 | 32.3 | 3365.1 | 1395.1 | 8853.4 |
| | LB | 320.1 | 0.0 | 0.0 | 0.0 | 0.0 | 0.0 | 326.2 | 377.7 | 3312.8 |
| | UB | 2898.0 | 223.8 | 701.1 | 1432.3 | 215.9 | 437.5 | 6758.5 | 2469.3 | 14514.1 |
| 10-06-22 | Median | 1117.8 | 13.6 | 20.0 | 32.7 | 4.3 | 66.1 | 1017.4 | 931.4 | 5297.9 |
| | LB | 129.8 | 0.0 | 0.0 | 0.0 | 0.0 | 0.0 | 0.0 | 330.9 | 244.2 |
| | UB | 2240.8 | 129.0 | 574.3 | 1229.4 | 131.4 | 335.6 | 5154.0 | 1551.8 | 11508.3 |
| 10-13-22 | Median | 878.9 | 14.2 | 2.7 | 10.3 | 1.5 | 43.0 | 1806.3 | 910.1 | 6231.5 |
| | LB | 25.2 | 0.0 | 0.0 | 0.0 | 0.0 | 0.0 | 0.0 | 245.3 | 492.8 |
| | UB | 1950.5 | 90.3 | 342.6 | 1087.6 | 125.6 | 315.5 | 5542.4 | 1603.3 | 12463.1 |

**Table 12s. Predicted cumulative number of newly forecasted monkeypox cases and 95% PIs for the weighted ensemble model.** Predicted cumulative number of newly forecasted monkeypox cases and 95% PIs for each 4-week ahead forecasts during the weeks of 7-28-2022 through 10-13-2022 based on a 10-week calibration period for the weighted sub-epidemic ensemble model.

| Unweighted - Ensemble Model | | | | | | | | | | |
|---|---|---|---|---|---|---|---|---|---|---|
| Week | | Brazil | Canada | England | France | Germany | Spain | US (OWID) | US (CDC) | World |
| 07-28-22 | Median | 9453.81 | 720.35 | 812.55 | 892.66 | 811.35 | 2501.29 | 3947.64 | 2787.79 | 20769.27 |
| | LB | 6136.76 | 583.03 | 251.00 | 154.97 | 286.08 | 83.57 | 1722.17 | 1467.06 | 13597.43 |
| | UB | 33425.20 | 866.54 | 1750.31 | 2298.95 | 2179.16 | 6289.50 | 14718.29 | 9995.46 | 31957.42 |
| 08-04-22 | Median | 1255.12 | 387.40 | 675.83 | 668.80 | 484.80 | 1301.58 | 21959.92 | 37056.90 | 33481.53 |
| | LB | 644.02 | 196.25 | 211.91 | 94.33 | 169.67 | 0.00 | 5763.36 | 9169.81 | 20344.07 |
| | UB | 2154.17 | 655.15 | 1314.96 | 1430.76 | 782.50 | 5118.04 | 53110.37 | 151064.53 | 61582.74 |
| 08-11-22 | Median | 3465.88 | 257.71 | 489.50 | 360.88 | 494.27 | 1556.86 | 7090.51 | 4526.81 | 21736.19 |
| | LB | 1630.85 | 134.47 | 73.84 | 0.00 | 153.42 | 0.00 | 2045.94 | 3008.05 | 15234.04 |
| | UB | 5086.97 | 390.58 | 1016.95 | 2905.87 | 1317.54 | 4640.71 | 23052.82 | 6832.69 | 31359.19 |
| 08-18-22 | Median | 1691.65 | 278.17 | 302.44 | 200.16 | 388.30 | 1898.78 | 7120.39 | 4791.23 | 17854.07 |
| | LB | 691.80 | 136.15 | 22.71 | 0.00 | 111.59 | 0.00 | 1552.00 | 2937.10 | 11341.66 |
| | UB | 3661.71 | 459.49 | 1339.64 | 1033.44 | 758.20 | 4211.14 | 29838.26 | 29020.30 | 25529.89 |
| 08-25-22 | Median | 2576.71 | 200.15 | 176.60 | 609.45 | 319.26 | 587.55 | 3807.32 | 3919.09 | 15401.01 |
| | LB | 1150.41 | 85.09 | 13.61 | 0.00 | 77.69 | 0.00 | 453.12 | 1989.57 | 9839.95 |
| | UB | 20773.99 | 808.69 | 451.66 | 4883.46 | 558.30 | 2537.73 | 9944.00 | 6237.37 | 22282.28 |
| 09-01-22 | Median | 1602.62 | 125.83 | 141.45 | 432.51 | 173.88 | 481.38 | 4827.79 | 3399.82 | 15298.59 |
| | LB | 706.05 | 2.07 | 1.95 | 0.00 | 0.00 | 0.00 | 428.49 | 1447.05 | 8529.28 |
| | UB | 2828.54 | 331.73 | 410.76 | 2643.14 | 630.89 | 1931.92 | 84582.47 | 16358.72 | 23416.86 |
| 09-08-22 | Median | 1966.47 | 89.40 | 138.00 | 406.53 | 70.80 | 307.24 | 3204.48 | 2692.46 | 9405.82 |
| | LB | 832.92 | 0.00 | 0.00 | 0.00 | 0.00 | 0.00 | 46.76 | 670.41 | 4754.03 |
| | UB | 14835.60 | 319.38 | 457.35 | 2483.85 | 277.94 | 1437.07 | 8769.74 | 9638.91 | 16766.10 |
| 09-15-22 | Median | 3133.06 | 88.63 | 136.83 | 332.19 | 74.15 | 267.11 | 5045.99 | 2832.23 | 15653.78 |
| | LB | 954.99 | 0.00 | 0.00 | 0.00 | 0.00 | 0.00 | 835.58 | 838.21 | 6437.87 |
| | UB | 11140.29 | 645.42 | 1366.06 | 2374.49 | 571.20 | 2121.99 | 230054.20 | 7442.24 | 42965.67 |

**Table 13s. Predicted cumulative number of newly forecasted monkeypox cases and 95% PIs for the unweighted ensemble model.** Predicted cumulative number of newly forecasted monkeypox cases and 95% PIs for each of the 4-week ahead forecasts during the weeks of 7-28-2022 through 9-15-2022 based on a 10-week calibration period for the unweighted sub-epidemic ensemble model. Only weeks in which the 4-week ahead forecast have past are included in this table.
